# Balanced deep learning on multi-omics networks identifies molecular subgroups of pathological brain aging

**DOI:** 10.64898/2026.02.18.26346567

**Authors:** Yacoub Abelard Njipouombe Nsangou, Maria A. Ulmer, Nicholas T. Seyfried, Jürgen Dönitz, Alzheimer’s Disease Metabolomics Consortium, The AMP-AD Consortium, Rima Kaddurah-Daouk, Gabi Kastenmüller, Matthias Arnold

**Affiliations:** Institute of Computational Biology, Helmholtz Zentrum München - German Research Center for Environmental Health, Neuherberg, Germany; Department of Medical Bioinformatics, University Medical Center Göttingen, Göttingen, Germany; Center for Neurodegenerative Disease, Emory University School of Medicine, Atlanta, Georgia, USA; Department of Neurology, Emory University School of Medicine, Atlanta, Georgia, USA; Department of Biochemistry, Emory School of Medicine, Atlanta, Georgia, USA; Department of Psychiatry and Behavioral Sciences, Duke University, Durham, NC, USA; Department of Medicine, Duke Center for the Study of Aging, and Duke Institute for Brain Sciences, Duke University School of Medicine, Durham, NC, Durham

**Author notes:** Metabolomics data used in preparation of this article were generated by the Alzheimer’s Disease Metabolomics Consortium (ADMC). As such, the investigators within the ADMC provided data but did not participate in analysis or writing of this report. A complete listing of ADMC investigators can be found at: https://sites.duke.edu/adnimetab/team/. The Accelerating Medicines Partnership - Alzheimer’s Disease (AMP-AD) Consortium: The full list of contributing scientists is available at https://adknowledgeportal.org/AMPADConsortiumMembers.

## Abstract

**Background:** Neurodegenerative diseases, including Alzheimer’s disease (AD), exhibit substantial clinical and molecular heterogeneity, complicating accurate diagnosis and development of effective therapies. Although multi-omics profiling provides unprecedented molecular resolution, systematic integration of high-dimensional, imbalanced data modalities with disease-relevant biological networks remains a methodological challenge.

**Methods:** We developed a network-informed multi-omics integration framework that combines data-driven molecular networks with brain transcriptomic, proteomic, and metabolomic data from 356 participants in the Religious Orders Study and Rush Memory and Aging Project (ROS/MAP). Utilizing 25 functional, data-driven multi-omics groups (DAD-MUGs) derived by graph embedding from the AD Atlas, co-expression-guided feature extraction and systematic two-phase feature balancing were applied to derive representative molecular features, which were subsequently learned using DAD-MUG–specific autoencoders to generate compact multi-omics expression scores. These were then used to identify molecular subgroups via hierarchical clustering. Subgroup robustness was assessed in an independent ROS/MAP cohort (n=327) using a two-round nested classification strategy.

**Results:** Subgroup identification based on DAD-MUG-derived expression scores resulted in five molecular subgroups exhibiting significant differences in cognitive performance and core neuropathological measures. Cross-validated nested classification using transcriptomic and proteomic data demonstrated reliable discrimination of subgroups. Applying these classifiers to the replication cohort, subgroup-trait association patterns showed strong agreement with discovery findings (Spearman ρ = 0.65). Differential expression analysis further revealed stage-dependent biological patterns of brain pathologies, ranging from early synaptic and immune activation to mitochondrial bioenergetic dysfunction at disease transition and proteostatic impairment in advanced stages.

**Conclusion:** Using a balanced, network-informed multi-omics integration framework, we identified five molecular subgroups of brain aging, including a reference control subgroup and a distinct mixed subgroup characterized by amyloid, vascular pathology, and early-life adversity. Three additional subgroups formed a structured spectrum comprising molecularly Alzheimer’s-like but cognitively and neuropathologically unimpaired At-risk controls, an intermediate stage, and typical Alzheimer’s disease, with tau pathology differentiating advanced disease, underscoring the value of molecular subgroup identification beyond clinical diagnosis.

## Introduction

Alzheimer’s disease (AD) is a progressive neurodegenerative disorder characterized by cognitive, functional, and behavioral impairments. It is the most common cause of dementia globally, accounting for 60–80% of the more than 50 million dementia cases worldwide ^1^. The disease manifests with heterogeneous clinical presentations and pathological patterns, posing significant challenges for accurate diagnosis, prognosis and therapeutic intervention ^2, 3^. Notably, this heterogeneity suggests that clinically defined AD may encompass multiple, partially overlapping molecular processes contributing to pathological brain aging. Traditional diagnostic approaches rely primarily on clinical assessments and neuroimaging, which often fail to capture the underlying molecular complexity contributing to this clinical heterogeneity ^4^.

Recent advances in high-throughput molecular profiling technologies have enabled comprehensive characterization of AD pathophysiology across multiple biological layers. Multi-omics approaches integrating transcriptomic, proteomic, and metabolomic data have demonstrated improved ability to identify disease-relevant molecular signatures compared to single-omics analyses ^5–9^. However, the integration of such high-dimensional, multi-scale molecular data presents substantial analytical challenges, including the curse of dimensionality, platform-specific biases and the need to preserve biologically meaningful relationships ^10, 11^.

Several computational strategies have been proposed to address these challenges. Network-based approaches have shown promise for structuring molecular features according to known biological pathways and interactions ^12, 13^, while weighted gene co-expression network analysis (WGCNA) ^14^ has been applied in AD research to uncover molecular co-expression modules associated with disease mechanisms in a data driven fashion ^15, 16^. In parallel, unsupervised deep learning methods, particularly autoencoders, have proven effective for multi-omics dimensionality reduction and pattern discovery across individual omics layers ^17–25^.

Despite these advances, existing multi-omics integration strategies face persistent limitations when attempting to link network-level biological information with patient-level stratification. For example, WGCNA has been successfully applied across multi-omics layers including transcriptomics, proteomics, and metabolomics ^26–28^, where modules are typically summarized through their module eigenfeature (termed eigengene for transcriptomic data, eigenprotein for proteomic data, and eigenmetabolite for metabolomic data) — defined as the first principal component — compressing module-wide variation into a single value and obscuring individual feature contributions. Alternative feature selection strategies based on connectivity or correlation typically also do not account for the full variance structure within modules ^14^. In addition, when organizing molecular features according to biological networks, large disparities in module size can introduce bias by overrepresenting larger pathways and underrepresenting smaller, independent of disease-relevance. In practice, unconstrained dimensionality reduction on multi-omics data can be dominated by large feature sets or technical structure, limiting interpretability and destabilizing patient stratification.

To date, no existing framework systematically combines (i) WGCNA-based identification of co-expression modules within individual omics layers, (ii) within-module selection of representative features beyond hub genes using principal component (PC) and correlation analysis, (iii) systematic feature balancing to address size disparities across variable-sized network-derived modules, and (iv) deep learning-based multi-omics integration for patient stratification. This gap is particularly relevant for AD research, where resources such as the AD Atlas—a comprehensive network-based integration resource for AD-relevant multi-omics data ^29^—remain underutilized for systematic multi-omics-based stratification.

Addressing this methodological gap is critical for advancing precision medicine applications in AD. The ability to systematically integrate data-driven disease network associations with balanced multi-omics feature representation could enable identification of molecular patient subgroups that reflect both established biological pathways and genuine inter-patient molecular variation. Such an approach could reveal clinically relevant disease subtypes that remain hidden when using either purely data-driven clustering methods or unbalanced network-informed approaches, ultimately supporting more personalized diagnostic and therapeutic strategies.

To address this gap, we developed and validated a two-part analytical framework that leverages data-driven multi-scale network structures from the AD Atlas for clinically meaningful patient stratification using integrated transcriptomic, proteomic, and metabolomic data. Part A applied deep graph representation learning and unsupervised clustering to the AD Atlas network to identify functional multi-omics clusters – hereafter referred to throughout the manuscript as data-driven multi-omics groups (DAD-MUGs) – representing data-driven functional organization. Part B identified co-expression modules using WGCNA for each omics layer separately, selected representative features through PCA-based correlation analysis, systematically balanced these features across the DAD-MUGs in a randomized way, and applied an autoencoder model to generate DAD-MUG-based expression scores (DMES) for robust patient subgroup identification and clinical association analysis.

## Methods

### Analytical Framework Overview

To address the challenge of integrating network-level biological data with patient omics data for stratification, we developed a four-stage analytical framework (**Figure 1**). First, we applied graph embedding to the AD Atlas to identify DAD-MUGs. Second, we performed systematic feature selection and balancing across these DAD-MUGs using patient multi-omics data. Third, we trained DAD-MUG-specific autoencoders to derive integrated molecular signatures. Finally, we applied hierarchical clustering to identify clinically relevant patient subgroups. The complete workflow and methodological details are presented below, with each analytical component corresponding to panels A-D in **Figure 1**.

**Figure 1.**
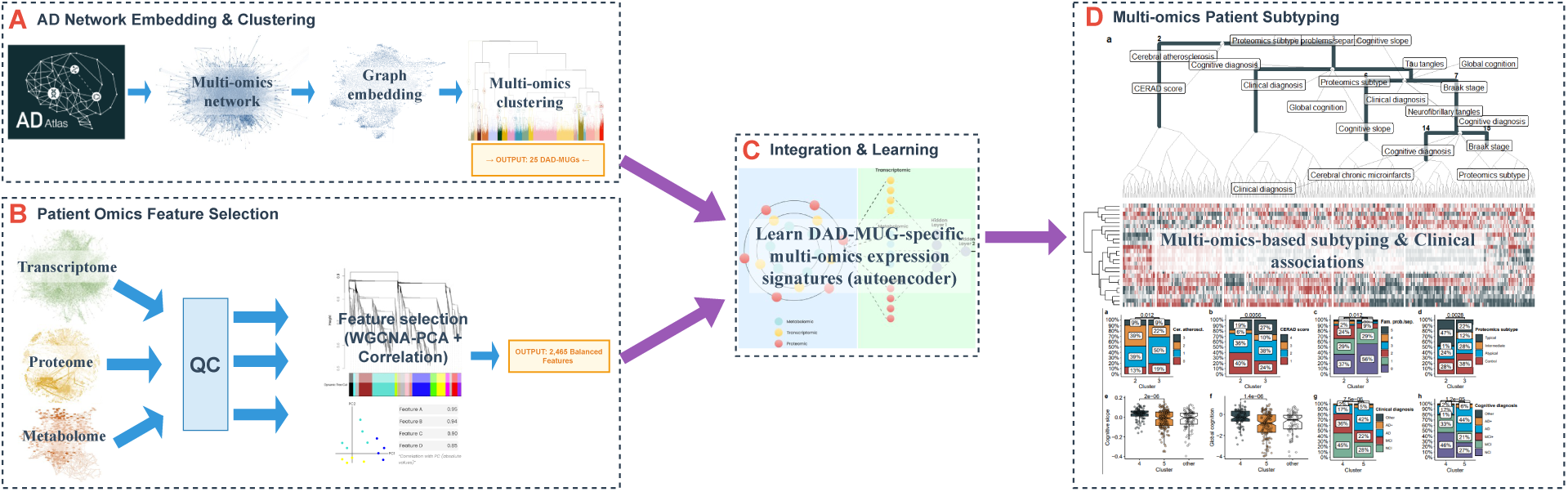
Network-informed multi-omics patient stratification framework in AD. **A)** Graph embedding and hierarchical clustering of the AD Atlas identify 25 DAD-MUGs representing functional modules spanning genes, proteins, and metabolites. **B)** Patient omics data (transcriptome, proteome, metabolome) undergo quality control and WGCNA-PCA-based feature selection, yielding 2,465 balanced features distributed across the 25 DAD-MUGs. **C)** DAD-MUG-specific autoencoders integrate balanced features to generate DMES (356 samples × 25 DAD-MUGs). **D)** Hierarchical clustering and statistical testing of DMES identify clinically relevant patient subgroups with significant clinical associations. Heatmaps display expression patterns and clinical association plots across identified subgroups.

### Cohort Characteristics and Omics Data Description

The Religious Order Study (ROS) and the complementary Rush Memory and Aging Project (MAP; together referred to as ROS/MAP) studies led by the Rush Alzheimer’s Disease Center are longitudinal cohort studies of aging and AD and designed to be used in joint analyses to maximize sample size ^30^. Both studies were approved by an Institutional Review Board at Rush University Medical Center. All participants signed an informed consent and a repository consent to allow their biospecimens and data to be used for ancillary studies. Further, all participants signed an Anatomic Gift Act for organ donation for research. More details can be found at www.radc.rush.edu.

We obtained processed multi-omics (transcriptomics, proteomics, metabolomics) data from brain tissue (dorsolateral prefrontal cortex or DLPFC) from the AD Knowledge Portal (https://adknowledgeportal.org). Data quality control and preprocessing protocols have been published before ^31–34^ and are summarized below.

For bulk-tissue RNA-seq data, we used processed data from the Accelerating Medicines Partnership - Alzheimer’s Disease (AMP-AD) RNA-seq Harmonization Study. Briefly, in the established pipeline, aligned reads in BAM files were converted to FASTQ after sorting paired-end reads. FASTQ files were aligned to the GENCODE24 (GRCh38) reference genome, gene counts were computed for each sample, and transcript abundance was estimated. Genes expressed in more than 1 CPM (read Counts Per Million total reads) in at least 50% of samples and diagnosis category were retained and normalized using conditional quantile normalization (CQN) to account for variations in gene length and GC content. The final dataset comprised 18,859 genes across 1,092 samples.

For isobaric tandem mass tag (TMT) mass spectrometry (MS)-based proteomics data, proteins whose measurements fell outside the 95% confidence interval of internal standards or showed high missingness were removed. Protein abundances were sample-scaled and log2-transformed. Sample outliers were identified using iterative principal component analysis. Subsequently, regression was used to remove the effects of proteomic sequencing batch and mass spectrometry reporter quantification mode. Missing values were imputed using k-nearest-neighbors (kNN). The final dataset comprised 10,030 proteins across 971 samples.

For MS-based untargeted metabolomics data, metabolites with high missingness were removed and concentrations normalized using cross-plate batch normalization and probabilistic quotient normalization. Multivariable outlier samples were removed, as were single concentration value outliers with <2.5% two-tailed probability to originate from the same normal distribution as the rest of the measurement values. Missing values were imputed using kNN. The final dataset comprised 667 metabolites across 509 samples.

All omics measurements were adjusted for post-mortem interval using regression analysis, and residualized values were used for all downstream analyses.

All omics features were further mapped one-to-one to DAD-MUGs (see next section) using standardized identifiers (gene/protein and metabolite IDs) consistent with AD Atlas curation. Features not represented in any DAD-MUG were excluded, reducing feature set sizes to 14,046 genes, 7,074 proteins, and 557 metabolites, respectively. The final discovery cohort with all three omics data types available consisted of 356 ROS/MAP participants, the non-overlapping replication cohort with transcriptomics and proteomics data only consisted of an additional 327 ROS/MAP participants (**Supplementary Table S6**).

On the side of demographic, molecular, and clinicopathological variables, we incorporated fifteen items: Sex, age at death, and education for demographics, as well as adverse events in early life; *APOE* ε4 genotype and brain tissue bulk RNA-seq- and proteomics-based subtype assignments published previously (both grouping individuals into controls vs. atypical, intermediate, and typical AD based on their respective omics fingerprints) ^35, 36^; clinical diagnosis and a composite for global cognition at the last antemortem visit, two neuropathological diagnostic assessments (CERAD score and Braak stage), quantitative measures for both brain amyloid load and tau tangle burden based on immunohistochemistry, and two variables for vascular pathologies (cerebral atherosclerosis and cerebral chronic microinfarcts).

### Definition of data-driven multi-omics groups (DAD-MUGs) using the AD Atlas

Data extraction, graph representation learning and unsupervised clustering for the definition of DAD-MUGs was performed as previously described ^29^. Briefly, we extracted an undirected graph from the AD Atlas that includes no self-loops and singular edges between nodes without differentiating edge or node types. Disease information was not included in this network, and only brain-specific information was included. Vector representations of nodes were learnt by sampling unbiased random walks over the network and using neighboring node pairs from these walk sequences as input-output training sets for a fully connected neural network. The resulting 130-dimensional vector representations (embeddings) of gene (collapsed transcriptomic and proteomic representation) and metabolite nodes were subjected to hierarchical clustering using Euclidean distance and Ward’s minimum variance method (ward.D2) ^37^. Clusters at height h = 30 determined 25 DAD-MUGs significantly enriched for AD-relevant biological domains.

### Single-Omics Feature Extraction

To ensure balanced representation while reducing dimensionality across our multi-omics datasets, we performed feature selection separately for each omics data using a combined weighted gene co-expression network analysis (WGCNA) ^14^ and principal component analysis (PCA) approach. The complete workflow is illustrated in **Figure 2**.

**Figure 2.**
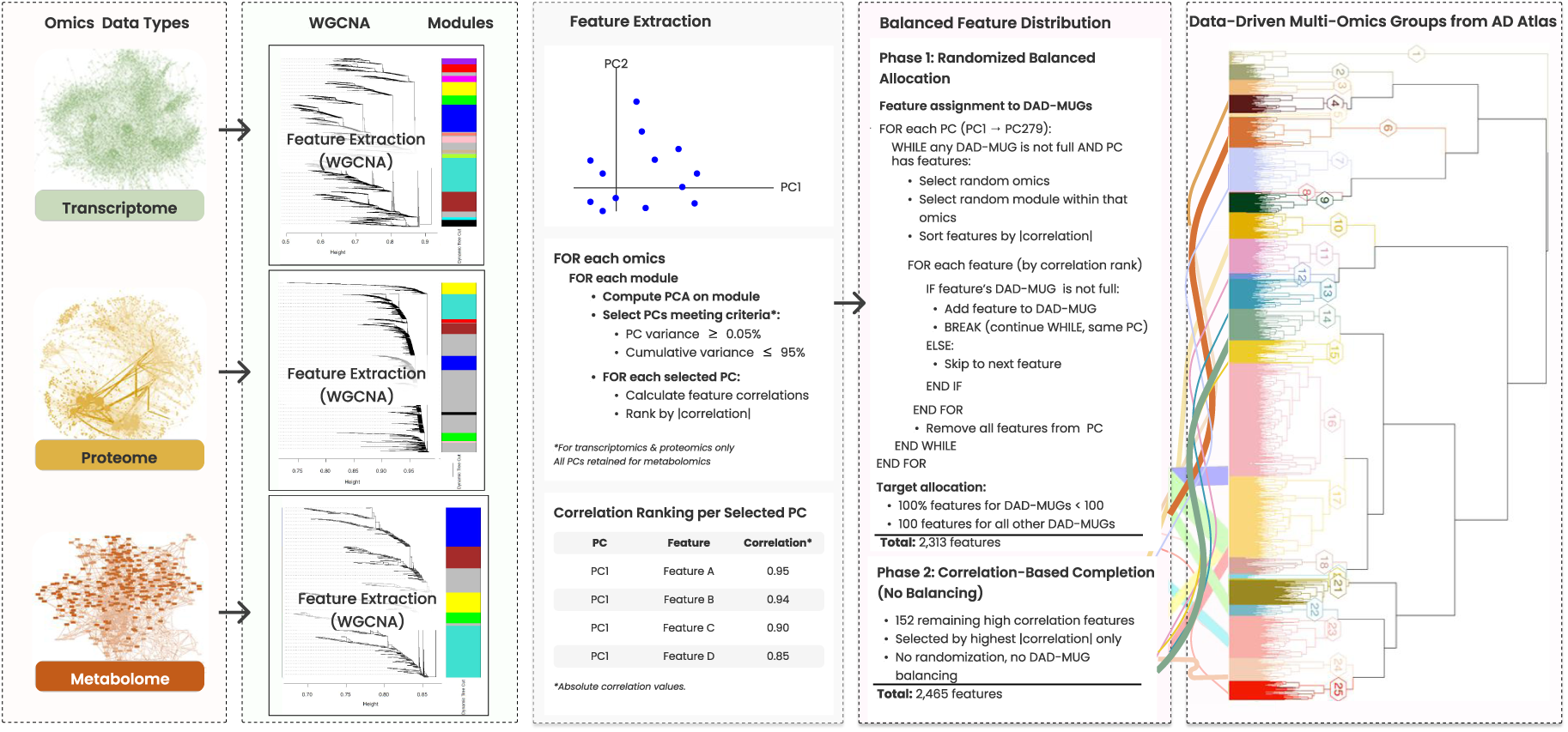
Multi-Omics Feature Extraction and Balanced Distribution Across DAD-MUGs. This figure illustrates the integrated workflow for extracting representative features from three omics types and distributing them in a balanced manner across 25 biologically meaningful functional groups derived from the AD Atlas. **Omics Data Types**. The analysis begins with three molecular datasets: transcriptomics (14,046 genes), proteomics (7,074 proteins) and metabolomics (557 metabolites) from 356 ROS/MAP participants. **WGCNA Modules**. WGCNA identifies co-expression modules within each omics type independently. Features are clustered based on topological overlap matrix (TOM) similarity using signed networks with data-specific soft-thresholding powers, yielding color-coded modules representing distinct co-expression patterns. **Feature Extraction**. Within each WGCNA module, PCA is performed to identify major axes of molecular variation. For transcriptomics and proteomics, PCs are retained based on two stopping criteria: (1) individual PC variance ≥ 0.05% and (2) cumulative variance ≤ 95%. For metabolomics, all PCs are retained to maximize variance coverage given its smaller initial dataset size. Across all three omics types and their respective modules, this yields 279 selected PCs (PC1 through PC279) representing the major patterns of molecular variation. For each selected PC, all module features are ranked by their absolute Pearson correlation with the PC scores, creating correlation-ranked original feature pools for subsequent selection. **Balanced Feature Distribution**. Features are allocated to 25 DAD-MUGs using a two-phase randomized balanced approach (detailed pseudocode in **Supplementary Algorithm**): **Phase 1**. Randomized Balanced Allocation (2,313 features): The algorithm iterates sequentially through the 279 PCs (PC1→ PC279). For each PC, it performs randomized sampling: (1) randomly selects an omics type (transcriptomics, proteomics or metabolomics), (2) randomly selects a WGCNA module within that omics type, (3) identifies features from that module associated with the current PC and sorts them by decreasing absolute correlation and (4) assigns the highest-correlation eligible feature to its corresponding DAD-MUG if that DAD-MUG has not yet reached its predefined target count. Target allocations ensure complete coverage of small DAD-MUGs (9, 15 and 89 features for DAD-MUGs 20, 8 and 5, respectively) and 100 features for all other DAD-MUGs (total: 2,313 features). This randomized strategy prevents systematic bias toward any single omics type or module while respecting biological feature-group assignments and correlation strength. **Phase 2**. Correlation-Based Completion (152 features): After **Phase 1** targets are met, the algorithm adds the top 152 remaining features ranked solely by their maximum absolute correlation across all PCs, without DAD-MUG balancing constraints, yielding a final set of 2,465 features distributed across all 25 DAD-MUGs. **Data-Driven Multi-Omics Groups from AD Atlas**. The right panel shows the hierarchical structure of the 25 DAD-MUGs (colored branches) derived from the network embedding of AD Atlas. Each DAD-MUG represents a functionally coherent biological module significantly enriched for AD-relevant processes. The selected 2,465 features are distributed across these groups, with connections (colored lines) indicating the flow of features from WGCNA modules through PCA-based extraction into their corresponding DAD-MUGs, ensuring balanced representation for downstream autoencoder-based compression and subgroup identification. For complete algorithmic details, see **Supplementary Algorithm**.

We applied standard WGCNA methodology following established procedures: quality control through hierarchical clustering, network construction using dataset-specific soft-thresholding powers, and module detection via dynamic tree cutting with minimum module sizes of 30 features. Complete details are provided in the **Supplementary Material**.

To extract representative features within each identified module for each of the three omics types, we applied a PCA-based approach:

1. PCA was performed on centered and scaled data to identify the principal components (PCs) of variation within each omics module.
2. For transcriptomics and proteomics data, PCs ordered by explained variance were extracted based on two stop criteria:

a. Individual variance threshold: next PC explaining less than 0.05% of the variance.
b. Cumulative variance threshold: extracted PCs collectively explaining more than 95% of the total variance.
3. For metabolomics data, we retained all PCs to capture the complete variance structure within each module. This approach was adopted because metabolomics represented the smallest dataset from the beginning, and we therefore biased the feature selection towards maximum coverage of the variance contained in the metabolomics data.
4. For each retained PC, we calculated correlations between original module features and PC scores. Correlations were used to rank original features for subsequent selection analysis across the three omics types.

Additional details on the selection criteria, implementation, and comparisons with classical, unbalanced WGCNA-based feature extraction, as well as more aggressive feature-reduction settings, are provided in the **Supplementary Material**.

### Multi-Omics Feature Balancing Across DAD-MUGs

Balancing was introduced to mitigate modality- and module-size dominance that can distort unsupervised compression and downstream subgrouping. Following single-omics feature extraction, we applied a two-phase balancing approach to ensure adequate representation of features across the 25 DAD-MUGs from network embedding analysis (**Figure 2**).

#### Phase 1: Randomized feature allocation ensuring minimum feature counts

We allocated features to meet predefined target counts for each DAD-MUG: complete coverage of DAD-MUGs with fewer than 100 features (n = 9, 15 and 89 features for the three smallest DAD-MUGs, respectively) and 100 features for all other DAD-MUGs. For each PC (proceeding sequentially from PC1 - PCN per module, where N denotes the last PC meeting our selection criteria), we iteratively:

1. Randomly selected an omics type (transcriptomics, proteomics or metabolomics).
2. Randomly selected a module within the selected omics type.
3. Identified original features from this module in order of decreasing correlation with the PC.
4. Assigned the first eligible feature to its corresponding DAD-MUG if the DAD-MUG had not yet reached its target count.

This process continued until all DAD-MUGs met their target counts (2,313 total features) or all PCs were processed.

#### Phase 2: Completion of feature allocation

For the remaining 152 PCs, the highest-correlating original features were used to finalize allocation, resulting in 2,465 features across all DAD-MUGs. The allocated 2,465 features were then extracted from the three multi-omics datasets to create lower-dimensional datasets specifically tailored for subsequent model training.

### DAD-MUG-based Expression Scores (DMES)

To extract meaningful patterns from the multi-omics data, we adapted PathME, a sparse autoencoder model originally designed for pathway-based analysis of genomic data ^38^. This unsupervised, multimodal neural network architecture enabled us to transform the previously selected transcriptomic, proteomic, and metabolomic features into a condensed DAD-MUG-based representation. Our adaptations to the model included the removal of the hyperbolic tangent (tanh) activation function in the second hidden layer to address potential output saturation issues and the application of Z-score normalization. As shown in **Supplementary Figure 1**, the model processes the three omics data types through an encoder-decoder architecture that compresses multi-omics information into meaningful representations. By training the adapted model on each of the 25 DAD-MUGs, we generated a condensed score matrix (356 patients by 25 DAD-MUGs) that captures the compressed biological information from the multi-omics data. This matrix provided the foundation for subsequent analysis. Full details regarding model configuration and training procedures are provided in **Supporting Material**.

### Subgroup Identification Analysis

To identify clinically relevant patient subgroups based on DMES, we employed the Subgroup Identification (SGI) software package ^39^. Our analysis followed three steps:

- **Data Preparation:** DMES were normalized via min-max scaling, transforming each feature to a [0, 1] range.
- **Hierarchical Clustering:** We generated a dendrogram using Euclidean distance and Ward’s minimum variance method (ward.D2) ^37^ to group patients by expression similarity. Potential subgroup splits (branch points) were tested for associations only if both resulting subgroups contained ≥10% (minimum n = 36) of the total sample.
- **Association Testing:** For each valid split, we tested associations between subgroups and clinicopathological variables using:

- Ordinal regression (probit link; rms::orm) for ordinal outcomes
- Kruskal-Wallis ^40^ tests for continuous variables
- Fisher’s exact test for binary categorical variables.

Demographic and other covariates were included among the variables tested for subgroup associations using the same procedures as for clinical traits. P-values were adjusted for multiple testing via Bonferroni correction to account for the number of subgroup comparisons tested across all valid dendrogram splits, with significance defined as adjusted p ≤ 0.05.

### Independent Replication

To validate SGI-derived molecular subgroups in the replication cohort lacking metabolomics, we trained nested classifiers in two rounds to predict discovery cohort subgroup labels from transcriptomic and proteomic features and applied these models to this cohort. In each round, we benchmarked multiple feature selection methods and classification algorithms using stratified cross-validation and selected the best-performing combinations for downstream subgroup-trait association testing. All feature selection steps were performed within the cross-validation procedure to avoid information leakage.

In the first round, the selected approach was Recursive Feature Elimination with Random Forest (RFE–RF) for feature selection and weighted Random Forest for four-class classification of the larger clusters in the dendrogram structure (Cluster 2, n = 86; Cluster 4, n = 95; Cluster 6, n = 62; Cluster 7, n = 113). In the second round, forward stepwise selection and LASSO-regularized logistic regression were selected for binary classification to further split Cluster 7 into Clusters 14 (n = 48) and 15 (n = 65). These methods were chosen based on their solid performance and their ability to yield linear, parsimonious models through stepwise variable selection and L1 shrinkage to limit overfitting. Both rounds employed inverse frequency class weighting to account for class imbalance and stratified 5-fold cross-validation to ensure proportional class representation in each fold. Statistical associations between predicted classes and clinical variables in the replication cohort were assessed analogously as for the discovery cohort. For functional annotation purposes, features were ranked by Gini importance to identify the top 50 most discriminative features. Functional enrichment analysis was performed on both the top 50 ranked features and all selected features using Gene Ontology (topGO), KEGG pathways (clusterProfiler) ^41^, Reactome pathways (ReactomePA) ^42^ and Disease Ontology (DOSE) ^43^, with Benjamini-Hochberg correction (FDR < 0.05). The top 50 features underwent AD Atlas- and literature-based annotations for AD associations. Detailed methodology for the machine learning (ML) pipeline and functional enrichment analysis is provided in **Supplementary Material**.

### Variance Decomposition Approaches

To identify and quantify the factors contributing to variation in DMES, we implemented two complementary variance decomposition analyses using the variancePartition package ^44^.

For clinical variable contribution analysis, we used linear mixed models to estimate how clinical and demographic variables contribute to DMES variability, treating categorical factors as random effects and continuous factors as fixed effects.

For multi-omic PCA-based analysis to assess the contributions of the different molecular layers, each omics dataset (transcriptomic, proteomic, metabolomic) was projected into PC space after centering and scaling. All PCs were retained and used as orthogonal predictors in variance partitioning models fitted with standard linear models. Feature-level contributions were derived using Equation 1, which weights normalized squared PC loadings by the variance explained by each PC and sums across PCs:

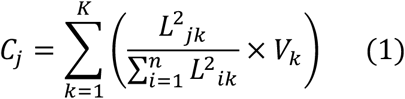

Where:

- *Cj* = contribution score (explained variance) for feature *j*.
- *Ljk*= Loading of feature *j* on principal component *k*.
- *Lik =* Loading of feature *i* on principal component *k* (where *i* indexes all features).
- *n* = total number of features in the omics dataset.
- *K* = number of principal components retained.
- *Vk* = proportion of variance explained by principal component *k*.
- The term 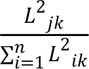 represents the normalized squared loading of feature *j* on PC *k*.

### Differential Association and Functional Enrichment Analysis

To characterize molecular differences between identified subgroups, we performed differential association analysis comparing three key clusters representing distinct disease stages: Controls, At-risk controls and AD cases. Wilcoxon rank-sum tests were applied separately to transcriptomics (14,046 genes), proteomics (7,074 proteins), and metabolomics (557 metabolites) data with Benjamini-Hochberg FDR correction (significance: adjusted p ≤ 0.05).

To identify biological functions distinguishing molecular subgroups, we performed functional enrichment analysis on significant genes and proteins (adjusted p ≤ 0.05) from each comparison against a background universe of all measured genes and proteins. Enrichment was performed using Gene Ontology (Biological Process, Molecular Function, Cellular Component) ^41^, KEGG pathways ^41^, Reactome pathways ^42^, and Disease Ontology ^43^ databases, with Benjamini-Hochberg correction (adjusted p-value < 0.05, q-value < 0.2). Detailed methodology is provided in **Supplementary Material**.

## Results

### Cohort Characteristics

The discovery cohort (n=356 ROS/MAP participants) spanned the full spectrum of cognitive aging, enabling molecular stratification across clinically and neuropathologically diverse individuals (**Table 1**). Participants had a mean age at death of 90.4 years (±6.4 SD) and were predominantly female (68.3%). At the last antemortem visit, 113 participants had no cognitive impairment (NCI), 101 had mild cognitive impairment (MCI), and 142 had dementia. As expected, *APOE* ε4 carrier frequency was higher in dementia (38.0%) than in NCI or MCI (both 16.8%), and both amyloid load and tau tangle burden increased across cognitive status categories.

**Table 1.** Demographic and Clinical Characteristics of the Discovery Cohort by Cognitive Status.

|  | **All (*n* = 356)** | **NCI (*n* = 113)** | **MCI (*n* = 101)** | **Dementia (*n* = 142)** |
| --- | --- | --- | --- | --- |
| **Sex (f/m)** | 243/113 | 74/39 | 71/30 | 98/44 |
| **Age** | 90.38 (±6.39) | 88.54 (±6.62) | 89.96 (±6.22) | 92.15 (±5.88) |
| **APOE4+/-** | 90/266 | 19/94 | 17/84 | 54/88 |
| **Amyloid load** | 4.83 (±4.64) | 3.14 (±3.71) | 4.59 (±4.87) | 6.33 (±4.68) |
| **Tau tangle load** | 6.54 (±7.93) | 3.15 (±3.12) | 5.13 (±5.46) | 10.22 (±10.23) |

NCI, No Cognitive Impairment; MCI, Mild Cognitive Impairment. Values are presented as mean (±SD) or count.

### Summary of Analytical Workflow

We applied a four-stage analytical framework to identify clinically relevant patient subgroups. First, we assigned omics features to 25 DAD-MUGs from the AD Atlas ^29^. Second, we performed feature selection and balancing from multi-omics data across these DAD-MUGs using WGCNA and PCA. Third, we trained DAD-MUG-specific autoencoders to generate condensed DMES for each patient. Finally, we applied subgroup identification and statistical testing to the DMES matrix to identify molecular subgroups with distinct clinical profiles. Results are presented below in the same sequence as the analytical stages shown in Figure 1. Because naïve, unbalanced integration can yield representations dominated by large feature sets rather than molecular structure, we benchmarked unbalanced and balanced strategies and report the selected DMES approach below..

### WGCNA-Based Omics Module Identification and PCA-Based Feature Selection

WGCNA provided a data-driven modular organization of each omics layer to enable downstream feature extraction and network-informed balancing. Across transcriptomics, proteomics, and metabolomics, we identified 30 co-expression modules in total (15, 9, and 6 modules, respectively). Notably, we treated grey modules as regular co-expression modules rather than outgroups, because their variance profiles were comparable to other modules (as reflected by principal component counts and explained variance), suggesting they capture coherent biological signals rather than unassigned noise. Module size distributions and feature counts are summarized in **Supplementary Figure S4.**

To prioritize representative original features within each WGCNA module, we used PCA to capture major axes of within-module variation and ranked module features by correlation with PC scores. For transcriptomics and proteomics modules, PCs were retained until they collectively explained ∼95% of variance (with an additional per-PC minimum variance threshold), whereas for metabolomics modules we retained all PCs to maximize variance coverage given the smaller feature universe. This analysis retained a total of 635 PCs across all modules (186 (29.3%) in transcriptomics, 279 (43.9%) in proteomics and 170 (26.8%) in metabolomics) and yielded a correlation-ranked candidate feature pool across omics layers for subsequent balanced allocation to DAD-MUGs. Detailed PC retention statistics and variance profiles are provided in **Supplementary Figure S5** and **Supplementary Data D1**.

### Feature Balancing Across DAD-MUGs

To prevent large network modules from dominating downstream representations, we applied a two-phase balancing procedure to distribute representative features across the 25 DAD-MUGs. This yielded a final set of 2,465 features drawn from 21,677 mapped omics features. In the first phase, we enforced a target of 100 features for each larger DAD-MUG and complete coverage for the smallest DAD-MUGs to ensure broad network representation for subsequent modeling (2,313 features total; **Figure 3A**). The second phase allocated the remaining 152 features to DAD-MUGs based on correlation ranks without constraints. Feature retention (overall rate = 11.4%) differed by omics layer, reflecting the smaller initial metabolomics feature universe (262/557 metabolites, 724/7,074 proteins, and 1,479/14,046 genes retained; **Supplementary Figure S5A**). The balanced feature set yielded DAD-MUGs with heterogeneous multi-omics composition, including both single-omic and multi-omic groups (**Figure 3A**; **Supplementary Figure S7**). Full feature counts per DAD-MUG and allocations are provided in **Supplementary Figure S7** and the **Supplementary Data D2**.

**Figure 3.**
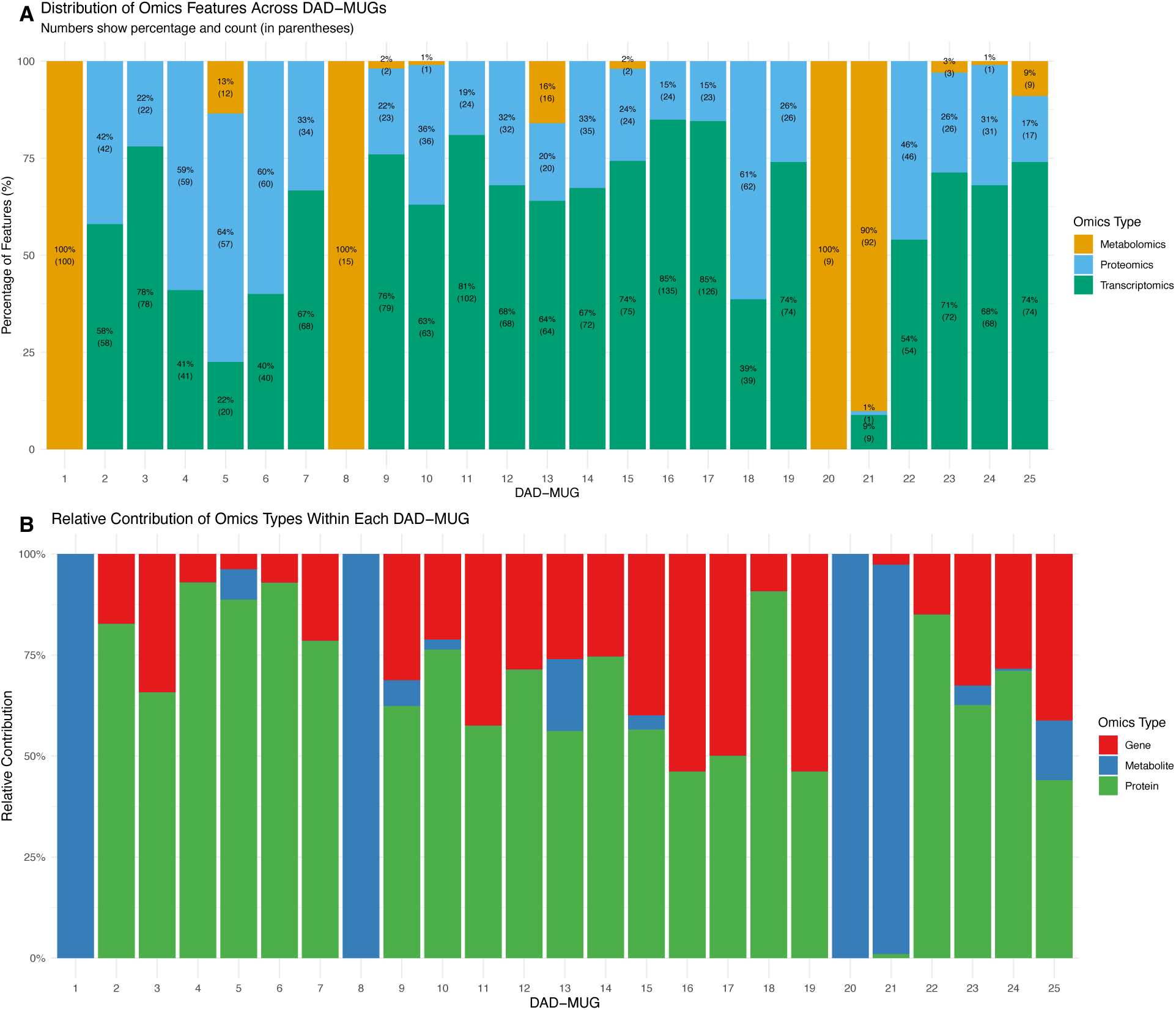
Distribution and relative contribution of multi-omics features across the 25 DAD-MUGs. **(A)** Stacked bar chart showing the percentage and absolute count (in parentheses) of features from each omics type (metabolomics, proteomics and transcriptomics) distributed across the 25 DAD-MUGs after feature balancing. DAD-MUGs 1, 8 and 20 consist entirely of metabolomic features, while DAD-MUG 21 is predominantly metabolomic (90%). DAD-MUGs 4, 5, 6 and 18 have a majority of proteomic features (≥59%). The remaining DAD-MUGs are primarily composed of transcriptomic features, with DAD-MUGs 11, 16 and 17 having the highest proportion (>80%). Each DAD-MUG contains approximately 100 features, except for the three smaller DAD-MUGs (5, 8 and 20). **(B)** Stacked bar plot showing the proportional contribution of each molecular data type (genes, proteins and metabolites) to the variance in DMES across all 25 DAD-MUGs. The y-axis represents the percentage contribution (0-100%) for each omics type, while the x-axis displays the 25 DAD-MUGs. Proteins dominated in 18 DAD-MUGs, while metabolites were exclusive contributors in DAD-MUGs 1, 8 and 20. DAD-MUGs with 0% contribution from a particular omics type indicate the absence of features from that molecular layer in the respective DAD-MUG.

### DAD-MUG-based Expression Scores (DMES) and variance decomposition

To select the DMES approach, we compared unbalanced versus balanced feature selection at two feature-reduction settings because unbalanced multi-omics compression can be biased toward larger feature sets and obscure biologically relevant structure.

Specifically, we evaluated (i) an unbalanced standard WGCNA-based baseline with modest feature reduction, (ii) an unbalanced coupled WGCNA+PCA variant with more aggressive reduction, and balanced feature allocation at (iii) a more aggressive and (iv) a more relaxed feature-reduction rate (**Supplement Text**, Section 1.3). The unbalanced WGCNA+PCA setting produced outlier-prone DMES values and increased sensitivity to demographic covariates (primarily sex, and to a lesser extent age at death), such that these factors dominated DMES variation and subgrouping. Randomized balancing mitigated these effects. However, at the more aggressive reduction setting the smaller feature budget constrained allocation and resulted in uneven representation across DAD-MUGs. We therefore used the balanced strategy with a more relaxed feature reduction rate for all subsequent analyses and refer to the resulting scores as DMES throughout. Clustering reproducibility analyses across candidate subgroup solutions (K=4–6) supported this more relaxed balanced strategy, showing consistently improved stability (higher Jaccard index and adjusted Rand index; **Supplementary Figure S11**).

Variance decomposition of DMES showed that clinical and demographic variables, including sex and age at death, contributed minimally to total variance (**Figure 4**; **Supplementary Data D3**). Decomposition by molecular layer revealed proteomic dominance in 18 of 25 DAD-MUGs, exclusive metabolite contributions in DAD-MUGs 1, 8, and 20 (and 96% contribution in DAD-MUG 21), and dominant transcriptomic contributions in DAD-MUGs 16 and 19 (54% each) (**Figure 3B**; **Supplementary Figure S12**). DAD-MUGs with 0% contribution from a given omics type reflect the absence of features from that layer in the respective DAD-MUG.

**Figure 4.**
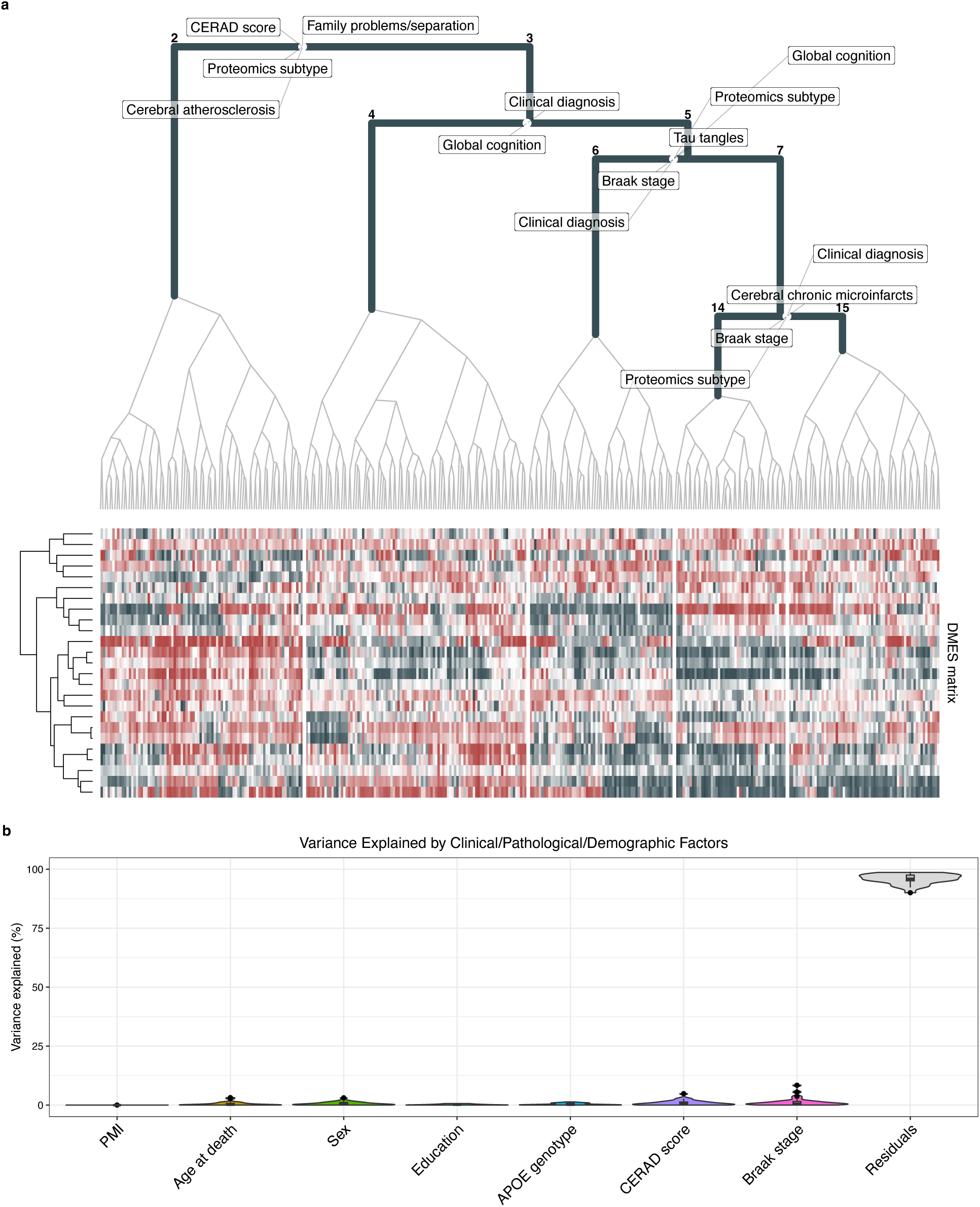
Hierarchical subgroup identification and variance decomposition based on DMES. **(a)** Hierarchical clustering dendrogram of 356 participants based on DMES (Ward’s method, Euclidean distance), with heatmap showing normalized expression levels across 25 DAD-MUGs. Numbers identify clusters at valid splits defined by a minimum size of both clusters of 10% of the total sample (36 samples). Labeled variables at each branching point represent significant (Bonferroni-adjusted p ≤ 0.05) differences distinguishing participant subgroups that emerge from that split and which are visualized in Figure 6. **(b)** Variance decomposition analysis showing the proportion of variance in DMES explained by clinical, pathological, and demographic factors. Most variables explain only a small fraction of variance, with about 92-97% remaining in residuals. This demonstrates that single DMES capture biological variation largely independent of conventional covariates, supporting their utility for molecular subtyping.

### Subgroup Identification Analysis

Using DMES values, we performed subgroup identification (SGI) ^39^ on the ROS/MAP discovery cohort (n = 356), yielding a hierarchical clustering structure with clinically and neuropathologically distinct molecular subgroups (**Figure 4**). Significant subgroup differences were observed for cognitive performance, clinical diagnosis, and neuropathological markers, alongside previously reported molecular AD subtype assignments (**Supplementary Data D4**).

Individuals in Cluster 2 exhibited higher vascular burden and greater exposure to early-life adverse events relative to Cluster 3. Cluster 2 also showed a higher proportion of pathological CERAD ratings (76% with CERAD 1–2 vs. 62% in Cluster 3), although this was not mirrored by quantitative amyloid load measures from immunohistochemistry. Tau measures did not differ between clusters. Interestingly, Cluster 2 contained a higher proportion of individuals with typical AD proteomic signatures and fewer control signatures than Cluster 3, likely mirroring the enrichment of pathological CERAD ratings (**Figure 5a-d**).

**Figure 5.**
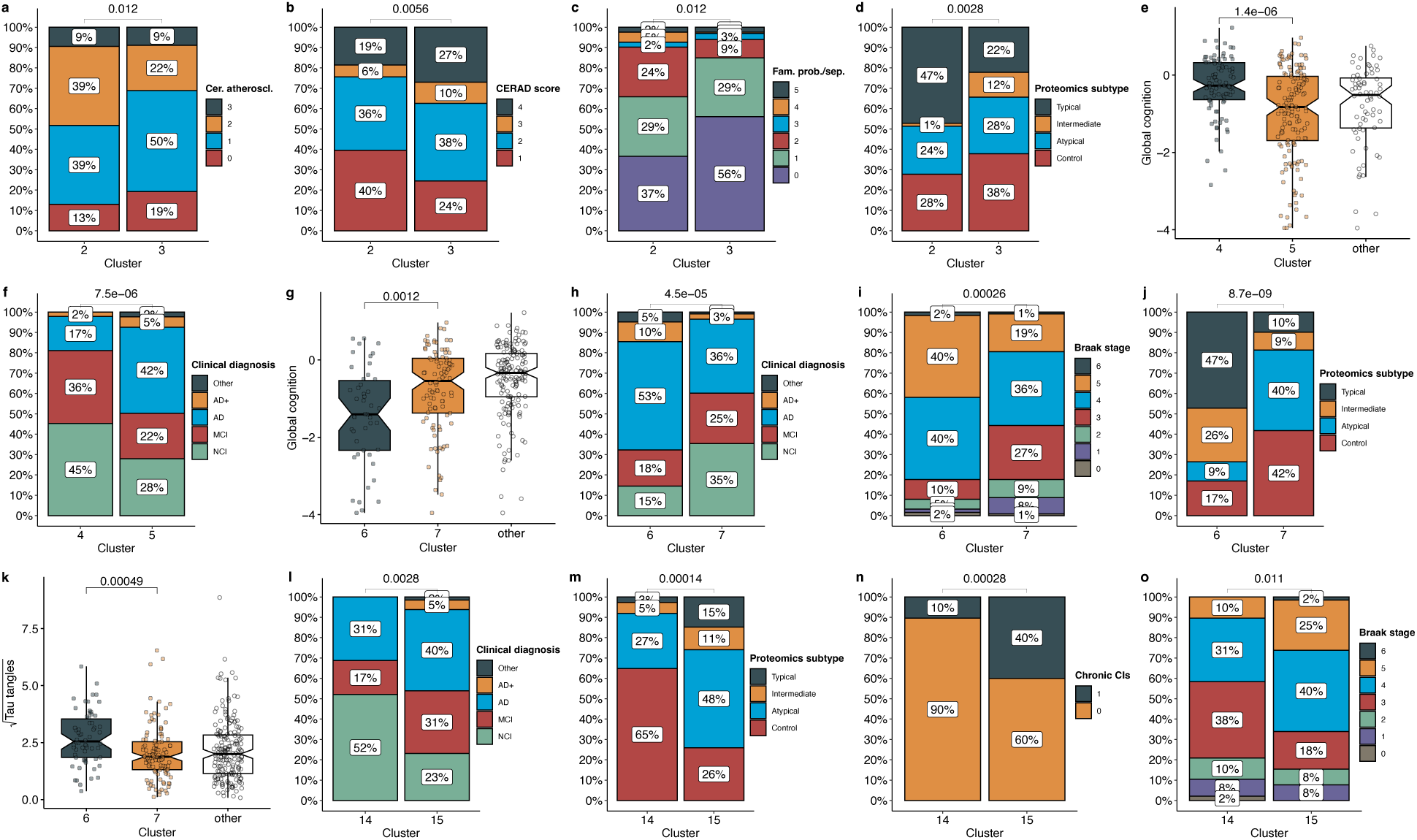
Clinical, cognitive and neuropathological characteristics distinguishing molecular subgroups. Comparative analyses across subgroups identified by SGI. **(a-d)** The primary split differentiates Clusters 2 and 3, with the former exhibiting more severe cerebral atherosclerosis, greater exposure to early-life adversity, lower CERAD scores and a higher proportion of proteomic signatures typical for AD. **(e-f)** Within Cluster 3, Clusters 4 and 5 diverge by cognitive status and clinical diagnosis, with Cluster 4 showing better preserved global cognition and a substantial proportion of cognitively unimpaired individuals. **(g-k)** Further subdivision (Clusters 6 vs. 7) characterizes Cluster 6 by more advanced Braak stages, elevated brain tau tangle burden, a substantially higher prevalence of AD-typical proteomic profiles, worse global cognition, and a large proportion of dementia patients. **(l-o)** Finally, Clusters 14 vs. 15 are distinguished by the number of chronic cerebral microinfarcts, AD proteomic signatures, Braak stage, and rates of dementia.

Cluster 3 was further divided into Clusters 4 and 5, separating a largely cognitively preserved subgroup from a clinically more impaired subgroup (**Figure 5e-f**). Cluster 4 exhibited high global cognition on average and contained a substantial fraction of individuals with no cognitive impairment, rendering it a molecular reference group for downstream stage comparisons. In contrast, Cluster 5 showed significantly lower global cognition and greater cognitive and functional impairment: 47% of individuals had a dementia diagnosis at their last antemortem visit, and only 28% were classified as NCI.

Clusters 6 and 7 emerged from Cluster 5 and diverged significantly by tau pathology, with corresponding differences in cognition, antemortem clinical diagnosis, and proteomic subtype composition (**Figure 5g–k**). Cluster 6 showed a tau-predominant profile with advanced Braak staging (82% stage IV–VI), markedly elevated tau tangle burden, and poorer cognitive outcomes. 63% of individuals had dementia at their last visit. Proteomic subtypes in Cluster 6 were enriched for typical/intermediate AD signatures. In contrast, Cluster 7 exhibited lower tau burden, fewer dementia diagnoses (39%) and proteomic profiles were dominated by atypical/control signatures.

Sub-clustering of Cluster 7 yielded two terminal groups, Clusters 14 and 15, which differed in cognitive diagnosis, proteomic subtype composition, neuropathological staging, and microvascular pathology (**Figure 5l–o**). Cluster 14 contained a higher proportion of cognitively unimpaired individuals (52%) and showed predominantly control-like proteomic subtype assignments (65%) despite multi-omic subgrouping within the AD-related branch, indicating a discordance between proteomics-derived neuropathology signatures and integrated multi-omic stratification. In contrast, Cluster 15 showed fewer cognitively unimpaired individuals (23%) and a higher burden of clinical impairment (46% dementia; 31% MCI), accompanied by more advanced Braak staging, a shift toward atypical proteomic subtypes (48%), and substantially higher prevalence of chronic cerebral microinfarcts (40% vs 10%).

Based on these differences, as well as overall distributions of key indices (global cognition and brain amyloid and tau tangle load; **Figure 6**), we assigned these clusters with the following molecularly grounded clinical subgroup designations:

**Figure 6.**
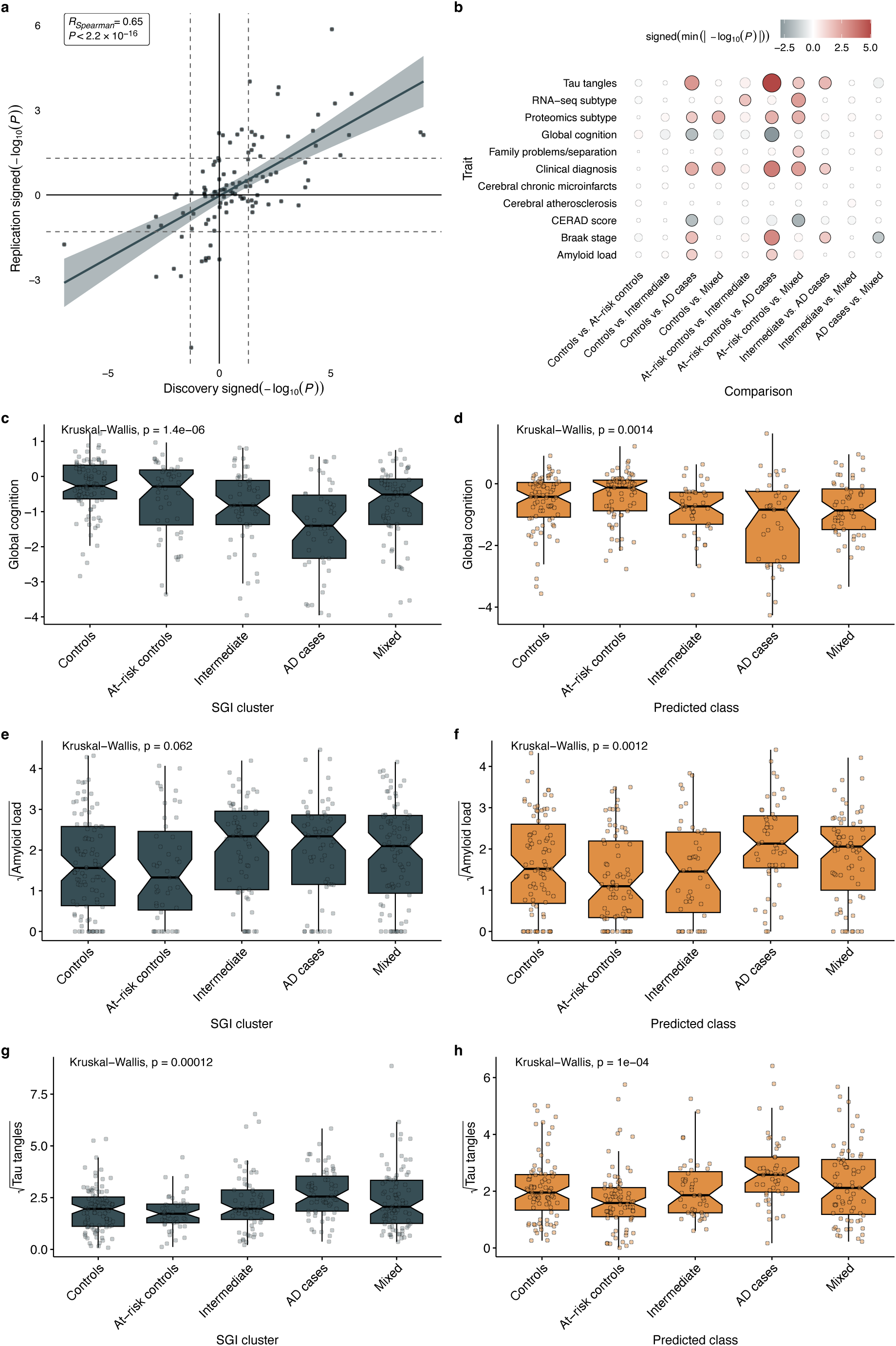
Independent validation of molecular subgroups demonstrates robust clinical and pathological associations. **(a)** Correlation between discovery and replication association statistics across all trait-subgroup comparisons. Each point represents one statistical test (trait × pairwise subgroup comparison). Signed −log10(P) values preserve effect direction and significance, with positive values indicating higher trait levels in the second subgroup relative to the first. Dashed lines indicate the p = 0.05 threshold on log10-scale. The strong positive correlation demonstrates overall reproducibility of subgroup-trait associations. **(b)** Heatmap showing pairwise associations of molecular subgroups with eleven clinicopathological traits reproducible across discovery and replication cohorts. Point size indicates the minimum absolute −log10(P) observed in both cohorts; color indicates effect direction (darkblue: negative association; darkred: positive association). Solid points indicate associations significant in both discovery and replication cohort; transparent points indicate significance in one or no cohort. **(c-d)** Global cognitive function across SGI-identified subgroups in the discovery cohort (**c**, n = 356) and ML-predicted classes in the replication cohort (**d**, n = 327). **(e-f)** Square root-transformed brain amyloid load across molecular subgroups in discovery (**e**) and replication (**f**) cohorts. **(g-h)** Square root-transformed brain tau tangle burden across molecular subgroups in discovery (**g**) and replication (**h**) cohorts.

We designated Cluster 2 as “Mixed” (n=86), defined by molecular separation from the other groups with enrichments for cerebral atherosclerosis, early-life adversity, and advanced CERAD ratings, but without corresponding tau burden; Cluster 4 as “Controls” (n=95), molecularly positioned outside the AD-related branch and characterized by preserved cognition and low dementia prevalence; Cluster 6 as “AD cases” (n=62), marked by substantial tau pathology, reduced cognitive function, and high dementia prevalence; Cluster 14 as “At-risk controls” (n=48), positioned within the AD-related molecular branch yet characterized by preserved cognition and low neuropathological burden with predominantly control-like proteomic subtype assignments; and Cluster 15 as “Intermediate” (n=65), also positioned within the AD-branch but clinicopathologically intermediate between At-risk controls and AD cases, with increased microvascular lesion burden.

### Independent Replication

To validate the clinical relevance of the identified molecular subgroups and identify discriminative molecular features, we trained nested classifiers for the SGI-detected clusters in two rounds using systematic feature selection and classification modeling on dual-omics data (transcriptomics and proteomics). In the first round, Recursive Feature Elimination with Random Forest (RFE-RF) selected 500 discriminative features (27 genes, 473 proteins), and a weighted Random Forest model achieved strong cross-validated performance (83.7% overall accuracy, 88.7% balanced accuracy, Cohen’s κ = 0.779; **Supplementary Table S4**). In the second round, forward selection identified 6 discriminative features (1 gene, 5 proteins) to subdivide Cluster 7 into Clusters 14 and 15, and a LASSO logistic regression model showed excellent performance (AUC = 0.998, balanced accuracy = 96.4%, Cohen’s κ = 0.928; **Supplementary Table S5**). Functional enrichment and annotation analyses of the selected feature sets (top-ranked and full sets) identified biologically coherent pathway signatures and highlighted both previously implicated and putative AD-related candidates (FDR < 0.05; **Supplementary Figure S14** and **S15** and **Supplementary Data D5**).

Applying the trained classifiers to an independent ROS/MAP replication cohort (n = 327; **Supplementary Table S6**) supported generalizability and clinical reproducibility of subgroup characteristics. Across all molecular subgroup comparisons and 11 clinicopathological traits, discovery and replication association statistics showed strong concordance (Spearman ρ = 0.65; **Figure 6a**; **Supplementary Data D6**). Predicted subgroup assignments in the replication cohort recapitulated key clinical and neuropathological gradients, including associations with global cognition, amyloid burden, and tau pathology (**Figure 6c–h**), with consistent effect directions relative to the discovery cohort. Replicated subgroup-trait patterns were most pronounced for tau pathology, cognitive function, and clinical diagnosis (**Figure 6b**). Notably, tau pathology showed particularly robust progressive gradients across molecular subgroups in both discovery (p=1.2×10^−4^) and replication (p=1.0×10^−4^) cohorts, with clear stratification from Controls through At-risk controls and Intermediate states to AD cases.

### Molecular Transitions Across Disease Stages Reveal Progressive Dysfunction

To elucidate molecular changes across subgroup-defined disease stages, we performed differential association analyses across three molecular subgroups: **Controls** (Cluster 4, n=95), **At-risk controls** (Cluster 14, n=48), and **AD cases** (Cluster 6, n=62), leveraging transcriptomic, proteomic, and metabolomic data. We conducted three pairwise comparisons using Wilcoxon rank-sum tests with Benjamini–Hochberg FDR correction (FDR < 0.05): Controls vs At-risk controls (early vulnerability), At-risk controls vs AD cases (disease transition), and Controls vs AD cases (full disease contrast). Differential testing revealed extensive molecular remodeling across all comparisons. In early vulnerability, 7,649 features were significant (173 metabolites, 2,508 proteins, 4,968 genes); the disease transition comparison yielded 11,270 significant features (227 metabolites, 3,318 proteins, 7,725 genes); and the full disease contrast yielded 9,624 significant features (94 metabolites, 3,298 proteins, 6,232 genes; **Supplementary Figure S13**, **Supplementary Data D7**). Functional enrichment analysis of significantly dysregulated genes and proteins revealed three distinct stages of molecular dysfunction (**Figures S16-S18**, **Supplementary Data D8**).

#### Early Molecular Vulnerability (Controls vs. At-risk controls)

This comparison showed molecular signatures consistent with early pathological changes preceding overt cognitive decline. KEGG pathway analysis identified significant enrichment for Alzheimer’s disease and other neurodegenerative pathways (Parkinson’s, Huntington’s, prion disease), indicating AD-like molecular patterns in clinically and neuropathologically unimpaired individuals. GO Biological Process terms were enriched for immune effector processes, synapse organization, and vesicle-mediated transport. Disease Ontology enrichments included respiratory diseases, ischemia, and autoimmune conditions. Cellular Component terms highlighted presynaptic structures and mitochondrial inner membrane, and Molecular Function terms emphasized NADH dehydrogenase and GTPase activity. Reactome analysis further supported neuroinflammatory and immune signaling (e.g., neutrophil degranulation, interleukin signaling, B cell receptor signaling), alongside enrichment for neurotransmitter receptor function, postsynaptic signaling, and mitochondrial translation.

#### Disease Transition (At-risk controls vs. AD cases)

This comparison revealed a strong enrichment of mitochondrial and bioenergetic pathways. This transition contrast yielded the largest number of dysregulated features among the three comparisons (11,270 features). GO Biological Process terms were dominated by oxidative phosphorylation, mitochondrial ATP synthesis coupled electron transport, cellular respiration, and aerobic electron transport chain, while Cellular Component analysis highlighted respiratory chain complexes and NADH dehydrogenase complex. KEGG and Reactome analyses further supported a prominent metabolic/mitochondrial signature, including enrichments for Alzheimer’s disease, oxidative phosphorylation, the tricarboxylic acid (TCA) cycle, and respiratory electron transport. Synapse organization and synaptic vesicle cycle terms remained prominent alongside ribosomal structures. Immune-related enrichments included T-cell receptor signaling, complement and coagulation cascades, and Fc gamma R-mediated phagocytosis.

### Established Disease (Controls vs AD cases)

The full disease contrast showed enrichments consistent with proteostasis impairment and translational dysregulation. GO Biological Process terms were enriched for post-transcriptional regulation of gene expression, regulation of translation, mRNA catabolic process, and mitochondrial translation. Cellular Component terms were dominated by ribosomal structures (ribosome, cytosolic ribosome, mitochondrial ribosome, ribosomal subunits), while Molecular Function terms emphasized translation regulator activity and structural constituent of ribosome. Reactome analysis further highlighted translation-related pathways (eukaryotic translation initiation/termination, nonsense-mediated decay, cap-dependent translation initiation) alongside mitochondrial translation processes. KEGG analysis additionally identified enrichments related to cytoskeletal organization (endocytosis, regulation of actin cytoskeleton, focal adhesion) and several infection-associated pathways, which may reflect innate immune and stress-response signaling.

Complete differential association statistics and enrichment results are provided in **Supplementary Data D7**, **Supplementary Data D8** and **Supplementary Figures S16-S18**.

## Discussion

In this study, we demonstrate that integrating brain multi-omics data with a multi-scale network structure enables stratification of pathological brain aging, yielding clinically meaningful molecular subgroups both along the AD spectrum and a molecularly distinct vascular-enriched mixed profile. Following deep graph representation learning and hierarchical clustering of the multi-scale AD Atlas network to obtain 25 AD-relevant, functional multi-omics groups (DAD-MUGs), we developed an analytical framework combining WGCNA with PCA-based feature extraction, systematic two-phase feature balancing across DAD-MUGs, and DAD-MUG–specific autoencoder models to generate DAD-MUG-based expression scores (DMESs) for patient subgroup identification. Using DMESs, we identified five molecular subgroups that differed in cognition, neuropathology, and clinical diagnosis, including a clinically and neuropathologically unimpaired but molecularly AD-like At-risk controls subgroup and a distinct Mixed subgroup with vascular pathology and early-life adversity signals. These findings support the utility of balanced, network-informed deep learning to uncover molecular heterogeneity that may not be fully captured by clinical diagnosis.

### Balanced deep learning on multi-omics networks

Our approach represents an advanced framework for multi-omics integration in pathological brain aging within a network context. While WGCNA has been extensively used for co-expression network analysis in AD research ^15, 16, 45–49^ and PCA for dimensionality reduction across AD applications including neuroimaging ^50–55^ and transcriptomics ^56, 57^, our combination of (i) network-derived functional grouping (DAD-MUGs), (ii) within-module feature extraction, (iii) systematic feature balancing across network groups, and (iv) DAD-MUG–specific sparse autoencoder compression is, to our knowledge, novel in this context.

This multi-step framework addresses central challenges in multi-omics integration: the curse of dimensionality, unequal feature distribution across biological network groups, and the need to preserve biologically meaningful structure derived from a data-driven molecular network. While WGCNA-, PCA-, and autoencoder-based analyses ^17–25, 58–61^ have each been applied in omics research in AD and beyond, our framework differs by explicitly coupling network-derived multi-omics grouping with systematic feature balancing and group-wise representation learning to support patient stratification.

Our results support this structure in that naïve and/or unbalanced compression can be dominated by large feature sets and yield unstable or confounded representations, whereas balanced selection improved DMES stability and downstream subgroup logic: By enforcing broad representation across all 25 DAD-MUGs while prioritizing informative features, the approach mitigated dominance by large feature sets and improved clustering stability. These findings underscore the value of explicit balancing when integrating multi-omics datasets with disparate feature universes.

Variance decomposition provides support to the interpretation of DMESs as molecular signals that are not driven primarily by conventional covariates. Clinical and demographic variables, including sex and age at death, explained only a small fraction of DMES variance, consistent with the observation that overly aggressive compression increased sensitivity to demographic covariates, whereas the selected balanced strategy mitigated this effect. Further, decomposition by molecular layers largely mirrored the DAD-MUG–specific feature composition, suggesting that DMES capture multi-omics signal in a module-resolved manner rather than collapsing to a single molecular layer.

### Molecular Subgroup Architecture Reveals Heterogeneity in Pathological Brain Aging

Using DMESs, we identified a coherent hierarchical subgroup structure yielding five molecular subgroups – Controls, At-risk controls, Intermediate, AD cases, and a Mixed subgroup – with distinct clinical and neuropathological profiles. A key aspect is the emergence of a structured spectrum in which molecular patterns align with progressive impairment, while also revealing discordant or modifying profiles. The identification of At-risk controls – molecularly AD-like yet cognitively and neuropathologically unimpaired on average – highlights that AD-related molecular signatures can precede conventional clinical and pathological readouts. Conversely, the Mixed subgroup demonstrates that vascular pathology and early-life adverse events can be associated with a distinct molecular profile that differs from other aging groups. This finding aligns with emerging theories linking early-life experiences to later neurodegenerative risk ^62–65^ and suggests that molecular stratification can capture processes relevant to pathological brain aging, including vascular-modified profiles, rather than reflecting a single canonical aging trajectory.

Independent replication supported the generalizability of these subgroup-associated clinical and pathological gradients. Using transcriptomic and proteomic features, nested classification achieved strong performance for multi-class and binary splits, enabling application to an independent ROS/MAP sample. Predicted subgroups in this replication cohort recapitulated discovery patterns across global cognition, amyloid burden, and tau pathology, with consistent effect directions and strong concordance of subgroup-trait association statistics (Spearman ρ = 0.65).

### Single-Omics Signatures Inform but Differ From Multi-Omic Stratification

Previously reported transcriptomics- and proteomics-derived AD subtype assignments provided an additional lens on subgroup biology. Across the subgrouping hierarchy, we observed broad shifts in proteomics subtype distributions consistent with increasing neuropathological burden, and transcriptomic subtypes weakly but reproducibly reflected the shift from At-risk controls to the Intermediate stage. However, we also observed meaningful discordances between transcriptomic and proteomic subtype labels and integrated multi-omic subgrouping. In particular, the At-risk controls subgroup showed predominantly control-like proteomic subtype assignments despite being embedded within the AD-related molecular branch. Similarly, both subtypings characterized the Mixed subgroup as being more similar to typical AD than the At-risk controls. This indicates that integrated multi-omics stratification may capture information not reducible to single-omics signatures alone, which is in line with conclusions from previous multi-omics work in AD ^66^.

### Tau Pathology and Vascular Burden Mark Transitions in Pathological Aging

The separation of intermediate and AD cases subgroups emphasized tau pathology as a key differentiator of advanced disease. The AD cases subgroup was characterized by advanced Braak staging, higher tangle tau burden, and worse cognitive outcomes, whereas subgroups with lower tau burden showed less affected or relatively preserved cognition despite AD-related molecular profiles. This supports the interpretation that tau-associated pathology aligns strongly with later-stage clinical impairment in this stratification, and that molecular subgrouping can help resolve intermediate states within an AD-related branch. Similarly, the intermediate subgroup showed increased microvascular lesion burden (chronic cerebral microinfarcts) alongside worse clinical impairment relative to At-risk controls. Together with the macrovascular (cerebral atherosclerosis) enrichment in the Mixed subgroup, these findings support the view that vascular pathologies can modify clinical expression and subgroup profiles within pathological brain aging. Of note, vascular trait associations were not consistently reproduced in the replication cohort, which may reflect reduced sensitivity for vascular-related molecular signals when metabolomics profiles are unavailable in the validation setting.

### Molecular Transitions Reveal Stage-Specific Mechanisms of Alzheimer’s Disease Progression

Our differential association and functional enrichment analyses provide a stage-resolved view of molecular alterations from controls across the AD-related subgroup spectrum by comparing three molecular subgroups – Controls, At-risk controls, and AD cases.

Early Vulnerability – preclinical molecular signatures in At-risk controls: The comparison of Controls vs. At-risk controls revealed coordinated changes spanning neurodegenerative pathway signatures, synaptic/vesicle processes, immune activation, and early mitochondrial stress. KEGG enrichment for Alzheimer’s disease alongside other neurodegenerative pathways and Reactome enrichment for neuronal system–related pathways suggests engagement of shared neurodegenerative programs in at-risk individuals before overt cognitive decline ^67, 68^. Enrichment of synapse organization and vesicle-mediated transport terms aligns with extensive evidence that synaptic dysfunction is an early and clinically relevant feature of AD ^69–72^. Immune-related GO/Reactome signatures and Disease Ontology enrichments implicating systemic inflammatory states are consistent with models in which peripheral inflammation and neuroimmune activation accompany early vulnerability ^73–76^. Finally, mitochondrial inner membrane and oxidative phosphorylation–related terms indicate early energy metabolism stress or compensatory mitochondrial responses. Together, these patterns support early vulnerability as a coordinated shift across synaptic, immune and metabolic systems.

Disease Transition – broad mitochondrial/bioenergetic disruption at conversion: Molecular differences between At-risk controls to AD cases showed the strongest enrichment of mitochondrial and bioenergetic pathways, consistent with a transition characterized by energy metabolism breakdown. GO and pathway enrichments highlighted oxidative phosphorylation, mitochondrial ATP synthesis–coupled electron transport, and respiratory chain components, with KEGG/Reactome support for TCA cycle and respiratory electron transport. This pattern is consistent with mitochondrial cascade models of AD in which age-related mitochondrial decline contributes centrally to pathogenesis ^77–81^. Importantly, synaptic and vesicle-related terms remained prominent, and translation/ribosome-related signals emerged together with persistent immune activation, suggesting that bioenergetic failure co-occurs with escalating synaptic and proteostasis stress during transition.

Established Disease – translational/proteostatic impairment in advanced pathology: The comparison of Controls vs. AD cases was dominated by translation-related and ribosomal terms, consistent with widespread proteostasis and protein synthesis/turnover dysregulation. Reactome enrichment for translation pathways and GO enrichment for post-transcriptional regulation and mRNA catabolic processes align with the concept of age- and disease-related proteostasis network decline as a hallmark of neurodegeneration ^82–84^. Mitochondrial dysfunction signatures persisted, and cytoskeletal/vesicle transport pathways (e.g., endocytosis, actin cytoskeleton regulation) were additionally enriched.

Collectively, these analyses support a coherent three-stage progression: (i) early synaptic/immune remodeling with emerging AD-related signatures, (ii) transition dominated by mitochondrial/bioenergetic disruption, and (iii) established disease marked by translational/proteostatic impairment with persistent metabolic and immune dysregulation. The repeated enrichment of Alzheimer’s disease pathways in early vulnerability and transition stages supports that the subgroup-defined comparisons capture AD-relevant biology across the disease spectrum.

### Limitations

Several limitations should be acknowledged. The pipeline’s multi-step complexity (network embedding, module-based feature extraction, balancing, autoencoding, and downstream inference) may limit adoption without appropriate computational and statistical expertise. The interpretability of network embedding and autoencoder representations remains inherently limited due to the black-box nature of deep learning approaches. While the autoencoder, for instance, effectively learns complex patterns from multi-omics data, the specific transformations within hidden layers do not directly reveal which molecular features contribute to the learned group-based representations. Although variance decomposition and feature-level analyses support biological interpretation, DMES remain learned representations rather than direct measurements. Finally, subgroup definitions were derived from cross-sectional post-mortem molecular profiles, so inferred stage relationships reflect molecular contrasts rather than longitudinal within-person progression.

### Conclusion and Future Directions

In summary, systematic integration of brain multi-omics data with AD Atlas network structure enables robust identification of clinically meaningful molecular subgroups across pathological brain aging. A key contribution of this work is a network-informed multi-omics stratification framework that explicitly addresses a central challenge in multi-omics integration – unbalanced compression dominated by large feature sets can obscure biologically meaningful structure – by combining data-driven functional network groups (DAD-MUGs), within-module feature extraction, systematic balancing across network groups, and DAD-MUG-specific autoencoder compression into DMES. The resulting subgroup hierarchy captures gradients of pathological aging, supports stage-resolved molecular interpretation of AD progression, and generalizes to an independent replication cohort. The framework is conceptually transferable to other complex diseases where multi-omics imbalance and patient heterogeneity complicate stratification, particularly when domain networks exist to anchor grouping and interpretation.

There are several research directions that emerge from our findings. First, testing subgroup structure and subgroup-trait gradients in additional cohorts – especially those with matched metabolomics – will clarify portability of DMES-based stratification and better resolve vascular-related dysregulation. Second, extending analyses to more diverse populations and additional brain regions will further address generalizability and specificity. Third, mechanistic follow-up of stage-associated signatures – particularly early synaptic/immune remodeling, transition-related mitochondrial disruption, and late-stage proteostasis impairment – will be important for translating subgroup biology into testable hypotheses. Finally, experimental validation of top discriminative molecular features from the replication classifiers, especially less-characterized candidates, could prioritize biomarkers and potential targets for future precision medicine approaches.

## Code availability

All code used for the analyses in this study is available at https://github.com/compneurobio/DAD-MUG.

## Data availability

ROS/MAP data are available via the AD Knowledge Portal (https://adknowledgeportal.org). The AD Knowledge Portal is a platform for accessing data, analyses, and tools generated by the Accelerating Medicines Partnership (AMP-AD) Target Discovery Program and other National Institute on Aging (NIA)-supported programs to enable open-science practices and accelerate translational learning. The data, analyses and tools are shared early in the research cycle without a publication embargo on secondary use. Data is available for general research use according to the following requirements for data access and data attribution (https://adknowledgeportal.synapse.org/Data%20Access). The full complement of clinical, phenotypic, and other data are available through the RADC Research Resource Sharing Hub (https://www.radc.rush.edu/).

For access to content described in this manuscript via the AD Knowledge Portal see: https://doi.org/10.7303/9618239 (ROS/MAP) and https://doi.org/10.7303/9618241 (RNAseq Harmonization Study).

## Acknowledgements

The results published here are in whole or in part based on data obtained from the AD Knowledge Portal (https://adknowledgeportal.org).

Study data were provided by the Rush Alzheimer’s Disease Center, Rush University Medical Center, Chicago. Data collection was supported through funding by NIA grants P30AG10161 (ROS), R01AG15819 (ROSMAP; genomics and RNAseq), R01AG17917 (MAP), R01AG30146, R01AG36042 (5hC methylation, ATACseq), RC2AG036547 (H3K9Ac), R01AG36836 (RNAseq), R01AG48015 (monocyte RNAseq) RF1AG57473 (single nucleus RNAseq), U01AG32984 (genomic and whole exome sequencing), U01AG46152 (ROSMAP AMP-AD, targeted proteomics), U01AG46161(TMT proteomics), U01AG61356 (whole genome sequencing, targeted proteomics, ROSMAP AMP-AD), the Illinois Department of Public Health (ROSMAP), and the Translational Genomics Research Institute (genomic). Additional phenotypic data can be requested at www.radc.rush.edu.

**Metabolomics:** Metabolomics data is provided by the Alzheimer’s Disease Metabolomics Consortium (ADMC) and funded wholly or in part by the following grants and supplements thereto: NIA R01AG046171, RF1AG051550, RF1AG057452, R01AG059093, RF1AG058942, U01AG061359, U19AG063744 and FNIH: #DAOU16AMPA awarded to Dr. Kaddurah-Daouk at Duke University in partnership with a large number of academic institutions. As such, the investigators within the ADMC, not listed specifically in this publication’s author’s list, provided data along with its pre-processing and prepared it for analysis, but did not participate in analysis or writing of this manuscript. A complete listing of ADMC investigators can be found at: https://sites.duke.edu/adnimetab/team/. The Metabolon datasets were generated at Metabolon and pre-processed by the ADMC.

**TMT Proteomics:** Study data were provided through the Accelerating Medicine Partnership for AD (U01AG046161 and U01AG061357) based on samples provided by the Rush Alzheimer’s Disease Center, Rush University Medical Center, Chicago. Data collection was supported through funding by NIA grants P30AG10161, R01AG15819, R01AG17917, R01AG30146, R01AG36836, U01AG32984, U01AG46152, the Illinois Department of Public Health, and the Translational Genomics Research Institute.

**RNAseq Harmonization Study:** Data generation was supported by the following NIH grants: P30AG10161, P30AG72975, R01AG15819, R01AG17917, R01AG036836, U01AG46152, U01AG61356, U01AG046139, P50 AG016574, R01 AG032990, U01AG046139, R01AG018023, U01AG006576, U01AG006786, R01AG025711, R01AG017216, R01AG003949, R01NS080820, U24NS072026, P30AG19610, U01AG046170, RF1AG057440, and U24AG061340, and the Cure PSP, Mayo and Michael J Fox foundations, Arizona Department of Health Services and the Arizona Biomedical Research Commission. We thank the participants of the Religious Order Study and Memory and Aging projects for the generous donation, the Sun Health Research Institute Brain and Body Donation Program, the Mayo Clinic Brain Bank, and the Mount Sinai/JJ Peters VA Medical Center NIH Brain and Tissue Repository. Data and analysis contributing investigators include Nilüfer Ertekin-Taner, Steven Younkin (Mayo Clinic, Jacksonville, FL), Todd Golde (University of Florida), Nathan Price (Institute for Systems Biology), David Bennett, Christopher Gaiteri (Rush University), Philip De Jager (Columbia University), Bin Zhang, Eric Schadt, Michelle Ehrlich, Vahram Haroutunian, Sam Gandy (Icahn School of Medicine at Mount Sinai), Koichi Iijima (National Center for Geriatrics and Gerontology, Japan), Scott Noggle (New York Stem Cell Foundation), Lara Mangravite (Sage Bionetworks).

## Author contributions

**Conceptualization:** GK, MA; **Data Curation:** YANN, MAU, GK, MA; **Formal Analysis:** YANN, MA; **Funding Acquisition:** NTS, RKD, GK, MA; **Methodology:** YANN, GK, MA; **Software:** YANN, MA; **Supervision:** GK, MA; **Visualization:** YANN, GK, MA; **Writing – original draft:** YANN, MA; **Writing – review & editing:** All authors.

## Competing interests

RKD, GK, MA are inventors on patent applications on the use of metabolomics in diseases of the central nervous system. They also hold equity in Chymia LLC, which had no role in this work. RKD is co-founder and holds equity in Metabolon, Inc., which had no role in this work. NTS is a co-founder and consultant of Emtherapro, and co-founder of Arc Proteomics and StitchRx. All other authors declare no competing interests.

## Notes

### Author Declarations

IRB of Duke University Health System gave ethical approval for this work.

