## Supplementary Material for "Balanced deep learning on multi-omics networks identifies molecular subgroups of pathological brain aging"

### 1. Feature Selection Using WGCNA and PCA

We utilized the Weighted Gene Co-expression Network Analysis (WGCNA) R package [1] and Principal Component Analysis (PCA) for multi-omics feature selection to identify representative features and reduce dimensionality across our datasets. This analysis was performed to select features for systematic balancing across 25 data-driven multi-omics groups (DAD-MUGs) identified through deep graph representation learning and unsupervised clustering of AD Atlas [2]. The complete workflow is illustrated in **Figure 2**.

#### 1.1 WGCNA-Based Module Identification

##### 1.1.1 Quality Control

To ensure robust downstream analysis, we performed quality control to assess data quality. First, we applied the goodSamplesGenes function to evaluate samples and features for excessive missing values or zero variance. We then performed hierarchical clustering using Euclidean distances and average linkage to assess sample distributions. PCA further aided in quality control by visualizing the first two principal components. These quality control steps ensured the reliability of downstream analyses across all three omics datasets.

##### 1.1.2 Soft-Thresholding Power Selection

Optimal soft-thresholding powers were determined using the pickSoftThreshold function, which evaluates a range of candidate powers to approximate scale-free network topology **(Figure S2**, left column). For each dataset, powers between 1 and 10 were initially assessed, extending the range up to 40 for proteomics and metabolomics and up to 100 for transcriptomics. Power selection was guided by two criteria:

- Scale-free topology fit index ($R^2$): evaluated to assess approximation to scale-free topology
- Mean connectivity: evaluated to ensure adequate network density **(Figure S2**, right column).

Although the automated procedure suggested higher soft-thresholding powers to maximize scale-free topology fit, lower powers were selected (4 for transcriptomics, 7 for proteomics, and 3 for metabolomics), as higher powers led to over-sparsified networks and excessive assignment of features to the grey module, whereas the selected powers produced more interconnected networks, reduced the number of unassigned features in the grey module, and improved biological interpretability.

##### 1.1.3 Network Construction and Module Detection

Signed co-expression networks were constructed for each omics dataset. Adjacency matrices were calculated using the adjacency() function with type="signed" and the dataset-specific soft powers determined in the previous step. These matrices were then converted to topological overlap matrices (TOM) using the TOMsimilarity() function. The dissimilarity TOM (1-TOM) was then used for hierarchical clustering to identify modules.

##### 1.1.4 Module Detection Using Dynamic Tree Cutting

We performed hierarchical clustering on the dissimilarity TOM using the average linkage method, resulting in feature dendrograms for each dataset (**Figure S3**). These dendrograms were then processed using the cutreeDynamic function with parameters shown in **Table S1**.

**Table S1.** Parameters used for dynamic tree cutting in module detection.

| **Parameter** | **Value** |
|---|---|
| Method | hybrid |
| Deep split | 0 |
| PAM stage | FALSE |
| Minimum module size | 30 |

The resulting module assignments were converted to color labels using the labels2colors function.

#### 1.2 Unbalanced PCA-Based Feature Selection Algorithm

Our feature selection approach within modules combined PCA with correlation analysis through the following algorithm:

1. For each identified module:
    a. Extract the module-specific expression data
    b. Perform PCA with centering and scaling
    c. Calculate the variance explained by each PC, both individually and cumulatively
    d. Select PCs based on the following thresholds:

            i. Individual PC variance ≥ 0.05%
            ii. Continue adding PCs until the cumulative variance reaches ≥ 95%
        e. For selected PCs, calculate the correlation between each feature in the module and the PC scores
2. Compile the selected features across all modules and datasets

Exception: For metabolomics data, all PCs were retained because the lower dimensionality (557 vs. 14,046 and 7,074 features for transcriptomics and proteomics, respectively) results in variance being concentrated in fewer components, with threshold-based filtering having minimal impact on the final PC set.

#### 1.3 Evolution of Feature Selection Approaches

We evaluated multiple feature selection strategies before selecting the expanded balanced approach described in the main paper.

##### 1.3.1 Initial Unbalanced Approach

In our first implementation, we applied WGCNA following the general procedure described in Section 1 (Feature Selection Using WGCNA and PCA), but with different parameter settings. The optimal soft-thresholding powers were automatically determined by the algorithm's scale-free topology fit index (82 for transcriptomics and 14 for proteomics).
Feature selection was based solely on module membership: we retained all features assigned to functional modules (excluding grey/unassigned features) without subsequent balancing across DAD-MUGs. For metabolomics, given the comparatively small number of features, all 557 metabolites were included without applying WGCNA.
This approach yielded 3,125 genes, 3,035 proteins and 557 metabolites for a total of 6,717 features. The resulting unbalanced distribution of features across the 25 DAD-MUGs is illustrated in **Figure S8**. The initial unbalanced approach was excluded because the large number of features compromised biological coherence in the resulting AD subtyping.

While this approach retained many features, we sought to further refine selection to focus on the most informative signals. This led to an intermediate feature selection approach that identified features most highly correlated with principal components explaining the majority of variance in each module, thereby reducing dimensionality while preserving essential biological information.

##### 1.3.2 Refined Feature Selection Approach

In this refined approach, we used WGCNA with fixed power thresholds (4 for transcriptomics, 7 for proteomics, and 3 for metabolomics) as described in Section 1 (Feature Selection Using WGCNA and PCA). This time, all modules were included (even the grey “unassigned” module), and the WGCNA procedure was extended to the metabolomics dataset.

For each module, we performed PCA to determine the number of principal components (PCs) required to explain at least 80% of the cumulative variance. For each PC, we calculated the correlation between PC scores and the original feature values, and selected the single feature with the maximum correlation, representing the feature that best captured that dimension of variation.

This procedure identified representative features from each module, resulting in 160 genes, 477 proteins, and 110 metabolites (747 features total). While the feature count was substantially reduced, the distribution across DAD-MUGs remained highly imbalanced (**Figure S9**), with some DAD-MUGs containing over 100 features and others fewer than 10.

Analysis of DAD-MUG-based expression scores (DMES) produced by our autoencoder model trained on this 747-feature set revealed two critical limitations:

- Extreme values in DMES: Some feature DAD-MUGs produced unusually high or low compressed scores, whereas others showed minimal variation.
- Biological bias: Certain biological factors became disproportionately dominant in the DMES. This bias was particularly evident in relation to sex, where the DMES at split points (clusters 2 and 3) showed a clear separation pattern ($p = 8.07 \times 10^{-88}$). Variance decomposition analysis quantified this effect, revealing that sex explained a disproportionately high percentage of variance in the DMES (max = 96.05%).

These limitations necessitated a balanced feature selection approach to ensure proportional representation across all DAD-MUGs.

##### 1.3.3 Compact Balanced Approach

In this implementation, we applied the same WGCNA, PCA, and balancing methodology as described in the main Methods section, but with more stringent selection criteria. Higher individual variance thresholds (0.68% vs. 0.05%) and lower cumulative variance thresholds (85% vs. 95%) were used for PC selection.

Additionally, smaller target counts were applied in the balancing procedure (15 features per DAD-MUG instead of 100, with 9 features for the smallest DAD-MUG). Following our two-phase balancing approach, Phase 1 allocated features to meet these targets (369

features), while Phase 2 added 130 highly correlated features to complete the allocation process.
This resulted in 499 total features (319 genes, 117 proteins, and 63 metabolites) compared to 2,465 in the final expanded balanced approach. The compact balanced approach achieved a more proportional distribution of features across all 25 DAD-MUGs (**Figure S10**).

##### 1.3.4 Expanded Balanced Approach (Final)

The expanded balanced approach was selected for all subsequent analyses.

### 2. DAD-MUG-based Expression Scores (DMES) Generation

In this work, we adapted the PathME sparse autoencoder model [3] to analyze multi-omics data from AD patients. PathME was chosen for its ability to perform dimensionality reduction while preserving biological relevance, allowing us to transform transcriptomic, proteomic and metabolomic profiles into a DAD-MUG-based representation.

#### 2.1 Key modifications to the Original Model

Several principal adjustments were made to optimize PathME for our application:

1. **Data and structural adaptation:** The original PathME maps genomic features (gene expression, miRNA expression, DNA methylation, and CNVs) to biological pathways from databases such as NCI, KEGG, and Reactome. In contrast, our approach uses AD Atlas [2] DAD-MUGs instead of pathway-based organization. We also integrated transcriptomic (T), proteomic (P), and metabolomic (M) measurements, combining multiple molecular levels rather than only genomic features.
2. **Activation function adjustment:** The hyperbolic tangent (tanh) activation function was removed from the second hidden layer (the bottleneck layer with a single node). This modification addresses the potential for gradient saturation, which occurs when inputs to tanh become extreme, pushing outputs toward asymptotic values (-1 or 1). Such saturation significantly diminishes gradient magnitude during backpropagation [4], which can impair model training, especially when dealing with datasets that contain highly varied features, such as multi-omics data.
3. **Weight initialization constraint**: While the original model explored multiple initialization strategies (normal, uniform, Xavier, and He), we exclusively used

Xavier initialization. This maintains consistent variance of activations across layers, helping prevent both vanishing and exploding gradients [4].

4. **Normalization strategy:** We replaced min-max normalization with Z-score normalization, which standardizes each feature by subtracting the mean and dividing by the standard deviation. This stabilizes training and ensures consistent scaling across multi-omics datasets.

#### 2.2 Model Architecture

As illustrated in **Figure S1**, our adapted model features an encoder-decoder structure:

- **Encoder**: The input layer accepts three separate data matrices representing transcriptomic (T), proteomic (P) and metabolomic (M) measurements for each patient. Each data type is processed through dedicated segments in the first hidden layer, where tanh activation functions are applied. These segments then feed into a single-node second hidden layer (without tanh activation), creating a unified compressed representation for each DAD-MUG.
- **Bottleneck layer**: This single-node layer represents the reduced-dimension embedding that captures the essential biological signal from all three omics types.
- **Decoder**: The bottleneck representation expands back through three parallel segments that reconstruct the original data dimensions, mirroring the encoder structure in reverse.

#### 2.3 Hyperparameter Optimization

We used Bayesian optimization (Tree Parzen Estimator, TPE) to efficiently explore the hyperparameter space, performing 30 evaluation cycles per DAD-MUG. Model performance was assessed using cross-validation with reconstruction error as the evaluation metric. The optimized hyperparameters are summarized in **Table S2**, and the final selected values for each DAD-MUG are in **Table S3**.

**Table S2.** Hyperparameters optimized in the multi-omics autoencoder model.

| **Hyperparameter** | **Description** | **Search Range** |
|---|---|---|
| Hidden layer dimensions | Number of neurons in each hidden layer | Varied based on DAD-MUG size |
| Mini-batch size | Number of samples per batch | {4, 8, 16} |
| Learning rate | Step size for model updates | $e^{-5}$ to $e^{-1}$ (log-uniform) |
| Sparsity penalty (λ) | Regularization for sparsity | {0, $e^{-8}$ to $e^{-1}$ (log-uniform)} |

| | | |
|---|---|---|
| Dropout keep probability | Probability of retaining neurons during training | {1, 0.5 to 1 (uniform)} |
| Optimizer | Optimization algorithm | {Adam, Nadam, SGD, Momentum, RMSProp} |
| Weight initializer | Method for initializing weights | Xavier |
| Alpha (scaling factor) | Scaling factor for loss terms | {0, 0 to 1 (uniform)} |

**Table S3.** Optimal Hyperparameter Sets for Each-DAD-MUG. DAD-MUG size represents the total number of genes, proteins, and metabolites in each DAD-MUG from the AD atlas network [2]. Each DAD-MUG may include all three omics features (genes, proteins, and metabolites), a combination of two features (genes and proteins) or just one feature (e.g., only metabolites as in DAD-MUG 8). The columns Input T, Input P, and Input M indicate the number of input features in the transcriptomic (T), proteomic (P) and metabolomic (M) data, respectively. The columns Hidden T, Hidden P and Hidden M represent the number of nodes in the first hidden layer that connect to the input layer for each i-th DAD-MUG during model training. Alpha (α) and Lambda (λ) represent regularization parameters.

| DAD-MUG number | DAD-MUG size | Input T | Input P | Input M | Hidden T | Hidden P | Hidden M | Batch Size | Alpha | Lambda | Learn Rate |
|---|---|---|---|---|---|---|---|---|---|---|---|
| 1 | 301 | 0 | 0 | 100 | - | - | 41 | 4 | 0 | 0 | 0.026 |
| 2 | 484 | 58 | 42 | 0 | 4 | 21 | - | 8 | 0.302 | 0 | 0.036 |
| 3 | 459 | 78 | 22 | 0 | 9 | 3 | - | 16 | 0 | 0 | 0.014 |
| 4 | 610 | 41 | 59 | 0 | 13 | 29 | - | 8 | 0.142 | 0 | 0.036 |
| 5 | 89 | 20 | 57 | 12 | 8 | 2 | 3 | 4 | 0 | 0.002 | 0.059 |
| 6 | 1135 | 40 | 60 | 0 | 12 | 24 | - | 16 | 0 | 0 | 0.10 |
| 7 | 1663 | 68 | 34 | 0 | 27 | 16 | - | 16 | 0 | 0 | 0.012 |
| 8 | 15 | 0 | 0 | 15 | - | - | 5 | 8 | 0.953 | 0 | 0.065 |
| 9 | 679 | 79 | 23 | 2 | 17 | 11 | 1 | 4 | 0 | 0 | 0.093 |
| 10 | 1020 | 63 | 36 | 1 | 12 | 8 | - | 8 | 0 | 0.003 | 0.032 |
| 11 | 1317 | 102 | 24 | 0 | 26 | 12 | - | 4 | 0 | 0 | 0.009 |
| 12 | 211 | 68 | 32 | 0 | 22 | 2 | - | 16 | 0 | 0.00038 | 0.098 |
| 13 | 989 | 64 | 20 | 16 | 32 | 5 | 1 | 16 | 0.805 | 0 | 0.039 |
| 14 | 1208 | 72 | 35 | 0 | 34 | 12 | - | 16 | 0 | 0.00037 | 0.025 |

| | | | | | | | | | | | |
|---|---|---|---|---|---|---|---|---|---|---|---|
| 15 | 697 | 75 | 24 | 2 | 4 | 10 | 1 | 16 | 0.270 | 0 | 0.053 |
| 16 | 4892 | 135 | 24 | 0 | 26 | 10 | - | 16 | 0.662 | 0 | 0.057 |
| 17 | 3273 | 126 | 23 | 0 | 43 | 7 | - | 8 | 0 | 0 | 0.041 |
| 18 | 744 | 39 | 62 | 0 | 17 | 11 | - | 8 | 0.226 | 0 | 0.057 |
| 19 | 113 | 74 | 26 | 0 | 7 | 10 | - | 8 | 0.027 | 0 | 0.035 |
| 20 | 9 | 0 | 0 | 9 | - | - | 3 | 8 | 0 | 0 | 0.09 |
| 21 | 573 | 9 | 1 | 92 | 3 | - | 25 | 16 | 0.894 | 0 | 0.013 |
| 22 | 261 | 54 | 46 | 0 | 2 | 4 | - | 8 | 0.128 | 0 | 0.01 |
| 23 | 1313 | 72 | 26 | 3 | 10 | 5 | 1 | 8 | 0.513 | 0 | 0.01 |
| 24 | 760 | 68 | 31 | 1 | 24 | 12 | - | 8 | 0 | 0 | 0.025 |
| 25 | 473 | 74 | 17 | 9 | 27 | 5 | 2 | 4 | 0 | 0 | 0.043 |

#### 2.4 Loss Function and Regularization Strategy

The loss function combines reconstruction accuracy with sparsity constraints at both DAD-MUG and individual feature levels. Specifically, we employed a composite loss that integrates:

- Mean squared error between input and reconstructed data to ensure accurate representation
- Group-lasso penalty (L2 regularization) to promote collective sparsity across features from the same omics layer
- Individual lasso penalty (L1 regularization) to identify the most informative individual measurements

#### 2.5 Training Process

Network weights were initialized using the Xavier method, which sets initial values based on the dimensions of each layer to maintain consistent variance throughout the network. This initialization strategy helps prevent gradient vanishing/exploding problems during training [4]. Each DAD-MUG model underwent training for up to 1,000 epochs, with early stopping implemented to prevent overfitting. Training was halted when the validation loss showed no improvement for 20 consecutive epochs. The convergence patterns for all

DAD-MUGs demonstrated successful minimization of the cost function, as shown in **Figure S6**.

#### 2.6 DMES Extraction

After training models for all 25 DAD-MUGs sequentially, we extracted the activation value from the bottleneck layer for each patient-DAD-MUG combination. These values represent the compressed biological signals specific to each DAD-MUG.
By concatenating these activations, we created a 356 × 25 matrix (patients × DAD-MUGs), which provides a dimensionally reduced representation of the original multi-omics data. This matrix served as the foundation for subsequent AD subtype identification analyses.

### 3. Evaluation of Clustering Consistency and Reproducibility

We implemented a systematic approach to evaluate the consistency and reproducibility of clustering results between our two balanced feature selection strategies (Compact balanced approach and Expanded balanced approach, see Section 1.3.3 and 1.3.4 of this Supplementary Text).
For each balanced implementation, we generated 10 distinct feature sets by applying the same balancing procedure with different random seeds. The adapted PATHME autoencoder model was trained on each feature set, producing 10 DMES matrices per approach. Hierarchical clustering was then performed on each DMES matrix using Euclidean distance and Ward's minimum variance method (ward.D2) [5], across a range of cluster numbers (K = 2–6).
To quantify clustering consistency, we calculated two complementary metrics:

1. The Jaccard similarity coefficient [6]: the ratio of the intersection to the union of sample pairs assigned to the same cluster across two different clusterings
2. The Adjusted Rand Index (ARI) [7], which measures the agreement between two partitions while adjusting for chance, was calculated using the R package mclust.

We performed all pairwise comparisons between matrices within each approach (45 comparisons per approach), computing both Jaccard and ARI values for each comparison and cluster number.

### 4. Independent Replication and Functional Enrichment Analysis

#### 4.1 Feature Selection and Classification Model Development

To validate the molecular subgroups identified through SGI analysis, we trained nested classifiers in two rounds using transcriptomics and proteomics data. Metabolomics was excluded due to lack of corresponding data in the validation cohort. In both rounds, we followed a two-step approach: (1) feature selection to identify discriminative molecular features and (2) classification model training using the selected features. Prior to final model selection, multiple feature selection methods and classification algorithms were systematically benchmarked using cross-validation to identify optimal combinations based on performance metrics.

**Round 1: Four-Class Classification**
We performed four-class classification for the larger clusters in the dendrogram structure: Cluster 2 (n=86), Cluster 4 (n=95), Cluster 6 (n=62), and Cluster 7 (n=113).

**Feature Selection:** Multiple feature selection approaches spanning filter, embedded and wrapper methods were systematically evaluated using cross-validation. Recursive Feature Elimination with Random Forest (RFE-RF) was selected based on superior accuracy and  applied to identify discriminative features for the four clusters. Using  the pre-processed dual-omics dataset (transcriptomics: 14,046 genes; proteomics: 7,074 proteins), the algorithm evaluated five candidate feature set sizes (100, 200, 500, 1000 and 1500) through 5-fold cross-validation with fixed folds. Feature importance was ranked by Gini importance, with the optimal subset of 500 features (27 genes, 473 proteins) selected based on cross-validation performance.

**Classification Model Training:** Multiple classification algorithms were benchmarked, and Weighted Random Forest achieved the highest balanced accuracy for four-class discrimination . The classifier was trained using the 500 selected features, with class weights set inversely proportional to class frequencies to address class imbalance in the discovery cohort (Cluster 4 n=95, Cluster 2 n=86, Cluster 6 n=62, Cluster 7 n=113). The model was configured with 500 trees and the mtry parameter was optimized through grid search (values: 2, 5, 10, 15). Model performance was evaluated using stratified 5-fold cross-validation to ensure proportional class representation in each fold.
The final model achieved a balanced accuracy of 88.7% (Cohen's kappa = 0.779) in cross-validation, indicating robust discrimination across all four molecular subtypes (**Table S4**).

**Round 2: Binary Classification of Cluster 7**
In the second round, we trained and applied a binary classifier to split Cluster 7 into Clusters 14 (n=48) and 15 (n=65).
**Feature Selection:** Given the limited sample size (n=113) and elevated overfitting risk, multiple feature selection approaches were evaluated. Forward selection with Bayesian Information Criterion (BIC) was selected based on its strong binary discrimination performance, stepwise parsimony, and linear approach. The method was applied to Cluster 7 samples using the same dual-omics data, identifying 6 discriminative features (1 gene, 5 proteins).
**Classification Model Training:** Multiple classification algorithms were benchmarked. LASSO logistic regression was selected based on optimal AUC, its linear and interpretable framework, and L1 regularization properties, which perform coefficient shrinkage toward zero to prevent overfitting and automatically select features – critical for stable performance with small sample sizes. The model was trained with class weights inversely proportional to class frequencies (48 vs 65) to account for class imbalance. It used L1 regularization (alpha = 1) with lambda values (0.001, 0.01, 0.1, 1) optimized via cross-validation. Performance was evaluated using stratified 5-fold cross-validation to ensure proportional class representation in each fold, with AUC as the primary metric.
The final binary model achieved exceptional performance with an AUC of 0.998, balanced accuracy of 96.4%, and Cohen's kappa of 0.928 in cross-validation, demonstrating near-perfect discrimination between Clusters 14 and 15 (**Table S5**).
**Validation**
The trained models from both rounds were applied sequentially to an independent validation cohort (n=327 samples) to assess generalizability and clinical validity. The Round 1 model (RFE-RF + Weighted Random Forest) first assigned samples to Clusters 2, 4, 6, or 7. Subsequently, the Round 2 model (forward selection + LASSO Logistic Regression) was applied only to samples predicted as Cluster 7, subdividing them into Clusters 14 and 15. Statistical associations between predicted classes and clinical variables in the validation cohort were assessed.

#### 4.2 Functional Enrichment Analysis Methodology

**Gene Set Preparation:** The top 50 most discriminative features (ranked by Gini importance from the weighted Random Forest model) and all 500 selected features were analyzed separately. Feature identifiers were converted to gene symbols using BioMart to enable cross-database enrichment analysis.
**Multi-Database Enrichment Strategy:** We employed a comprehensive enrichment analysis using four complementary approaches:

- **Gene Ontology (GO):** topGO package [8] with weight01 algorithm (accounts for GO hierarchy), Fisher's exact test, Benjamini-Hochberg FDR correction

- **KEGG Pathways:** clusterProfiler::enrichKEGG [9] with FDR correction
- **Reactome Pathways:** ReactomePA::enrichPathway [10] with FDR correction
- **Disease Ontology:** DOSE::enrichDO [11] with FDR correction

**Statistical Rigor:** Multiple testing correction was applied using Benjamini-Hochberg method (FDR < 0.05 considered significant).
**Visualization:** Complete enrichment results for both top 50 features and all 500 features are presented in **Figures S14-S15**, showing significant biological pathways, molecular functions, and disease associations.

**Table S4.** Performance of Weighted Random Forest with RFE-RF Feature Selection for Molecular Subtype Classification

| **Metric Catogory** | **Specific Metric** | **Value** |
|---|---|---|
| **Overall Performance** | | |
| | Cross-validation Accuracy | 83.7% |
| | Balanced Accuracy | 88.7% |
| | Cohen's Kappa | 0.779 |
| | Macro F1-Score | 0.830 |
| | Multiclass AUC | 0.956 |
| **Features Selected** | | |
| | Total Features | 500 |
| | Genes | 27 |
| | Proteins | 473 |
| **Class-Specific Sensitivity** | | |
| | Cluster 4 | 81.1% |
| | Cluster 2 | 88.4% |
| | Cluster 6 | 74.2% |
| | Cluster 7 | 87.6% |
| **Class-Specific Specificity** | | |
| | Cluster 4 | 95.0% |
| | Cluster 2 | 94.4% |
| | Cluster 6 | 96.3% |

| | | |
|---|---|---|
| | Cluster 7 | 92.2% |
| **Model Configuration** | | |
| | Algorithm | Weighted Random Forest |
| | Feature Selection | RFE-RF |
| | Number of Trees | 500 |
| | Validation Method | 5-fold stratified CV |

**Table S5.** Performance of LASSO Logistic Regression with Forward Selection for Binary Classification of Cluster 7

| **Metric Catogory** | **Specific Metric** | **Value** |
|---|---|---|
| **Overall Performance** | | |
| | Cross-validation Accuracy | 96.5% |
| | Balanced Accuracy | 96.4% |
| | Cohen's Kappa | 0.928 |
| | AUC | 0.998 |
| | F1-Score | 0.958 |
| | Sensitivity | 95.8% |
| | Specificity | 96.9% |
| **Features Selected** | | |
| | Total Features | 6 |
| | Genes | 1 |
| | Proteins | 5 |
| **Class-Specific Sensitivity** | | |
| | Cluster 14 | 95.8% |
| | Cluster 15 | 95.8% |
| **Class-Specific Specificity** | | |
| | Cluster 14 | 96.9% |
| | Cluster 15 | 96.9% |
| **Model Configuration** | | |

| | | |
|---|---|---|
| | Algorithm | LASSO Logistic Regression |
| | Feature Selection | Forward Selection (BIC) |
| | Regularization | L1 (alpha = 1) |
| | Lambda Range | 0.001 - 1 |
| | Validation Method | 5-fold stratified CV |

### 5. Differential Association and Functional Enrichment Analysis

#### 5.1 Differential Association Analysis

**Sample Selection and Grouping.** Three molecular subgroups were selected for differential analysis based on their distinct clinical and neuropathological profiles identified through Subgroup Identification (SGI) analysis: Control (Cluster 4, n=95), At-risk Control (Cluster 14, n=48) and disease-relevant state (Cluster 6, n=62).

**Statistical Testing.** For each of three pairwise comparisons (Cluster 4 vs 14, Cluster 14 vs 6, Cluster 4 vs 6), differential association testing was performed independently on transcriptomics (14,046 genes), proteomics (7,074 proteins) and metabolomics (557 metabolites) datasets using the two-sided Wilcoxon rank-sum test. Features with fewer than two observations in either group were excluded from testing. P-values were adjusted for multiple testing using the Benjamini-Hochberg false discovery rate (FDR) method, with FDR < 0.05 considered statistically significant. Summary statistics (mean, median, log2 fold change) were calculated for all significant features.

#### 5.2 Functional Enrichment Analysis

**Gene Set Preparation.** For each pairwise comparison, significant genes (FDR < 0.05) from transcriptomics and proteins from proteomics analyses were combined at the gene symbol level to create unified gene lists capturing both transcriptional and post-transcriptional regulation. Gene symbols were converted to Entrez Gene IDs using the bitr function from clusterProfiler (version 4.11.0.1) with the org.Hs.eg.db human genome annotation database (version 3.18.0). Only successfully mapped genes were included in downstream enrichment analyses.

**Background Gene Universe.** A background gene universe was constructed from all genes and proteins measured across transcriptomics (14,046 genes) and proteomics

(7,074 proteins) datasets. After gene symbol conversion to Entrez IDs, this yielded 13,671 unique Entrez IDs used as the statistical background for all enrichment tests.

**Enrichment Databases and Parameters.** Over-representation analysis was performed using four complementary annotation databases: Gene Ontology (GO; Biological Process, Molecular Function, Cellular Component) [7] tested using enrichGO, KEGG pathways tested using enrichKEGG [7], Reactome pathways tested using enrichPathway from ReactomePA (version 1.46.0) [8] and Disease Ontology tested using enrichDO from DOSE (version 3.28.2) [9]. All enrichment analyses used Benjamini-Hochberg multiple testing correction with significance thresholds of adjusted p-value < 0.05 and q-value < 0.2. For each enriched term, the following metrics were reported: gene ratio (proportion of input genes in the term), background ratio (proportion of background genes in the term), nominal p-value, adjusted p-value, q-value, gene count and contributing gene IDs.

**Output and Reporting.** Complete results for all features tested in differential association analysis, including p-values, adjusted p-values and fold changes, are provided in **Supplementary Data D7**. Complete enrichment results for all databases and comparisons, including all significantly enriched terms with associated statistics and gene lists, are provided in **Supplementary Data D8**.

**Table S6.** Demographic and Clinical Characteristics of the Replication Cohort by Cognitive Status.

| | **All** ***(n = 327)*** | **NCI** ***(n = 123)*** | **MCI** ***(n = 81)*** | **Dementia** ***(n = 123)*** |
|---|---|---|---|---|
| **Sex (f/m)** | 200 / 127 | 67 / 56 | 51 / 30 | 82 / 41 |
| **Age at death** | 88.30 (±6.69) | 86.44 (±6.64) | 89.32 (±6.52) | 89.48 (±6.48) |
| **APOE4+/-** | 76 / 251 | 20 / 103 | 15 / 66 | 41 / 82 |
| **Amyloid** | 3.97 (±4.11) | 3.11 (±3.84) | 4.01 (±4.28) | 4.78 (±4.12) |
| **Tau** | 5.88 (±6.65) | 2.93 (±2.63) | 4.92 (±4.50) | 9.42 (±8.68) |

NCI, No Cognitive Impairment; MCI, Mild Cognitive Impairment. Values are presented as mean (±SD) or count.

### Supplementary Data

**Data D1. Per-module PCA feature correlation statistics.** For each WGCNA module across transcriptomics, proteomics and metabolomics, features are listed alongside their assigned principal component (PC), the variance explained by that PC and the cumulative total variance explained across retained PCs within the module. PCs were retained until ~95% of within-module variance was explained (transcriptomics and proteomics) or all PCs were retained (metabolomics).

**Data D2. Complete feature balancing results across DAD-MUGs.** For each of the 2,465 selected features across transcriptomics, proteomics and metabolomics, feature identifier, assigned DAD-MUG, omics type, WGCNA module, principal component (PC), correlation with PC scores, absolute correlation and allocation phase are reported (Phase 1: randomized allocation to meet target counts; Phase 2: high-correlation feature completion).

**Data D3. Variance decomposition of DAD-MUG-based Expression Scores (DMES) by clinical and demographic variables.** For each of the 25 DAD-MUGs, the proportion of variance in the corresponding DMES explained by post-mortem interval (PMI), age at death, sex, education, APOE genotype, CERAD score and Braak stage is reported, alongside residual variance.

**Data D4. Pairwise clinical association results across hierarchical subgroup splits.** For each valid dendrogram split identified by the SGI algorithm, pairwise associations between subgroups and clinical variables are reported, including trait, compared cluster pair (cid1, cid2), raw and Bonferroni-adjusted p-values, test statistic and statistical test applied.

**Data D5. Complete functional enrichment results for discriminative feature sets.** Enrichment analysis results across Gene Ontology (Biological Process, Cellular Component, Molecular Function), KEGG pathways, Reactome pathways and Disease Ontology are provided in separate sheets for the top 50 and all 500 RFE-RF selected features.

**Data D6. Subgroup-trait association statistics for discovery and replication cohorts.** For each clinicopathological trait and pairwise subgroup comparison, association statistics, p-values and effect directions are reported for both the ROS/MAP discovery cohort (n = 356) and the independent replication cohort (n = 327).

**Data D7. Complete differential association statistics across pairwise subgroup comparisons.** For each feature across transcriptomics, proteomics and metabolomics, fold change, p-value and Benjamini-Hochberg-adjusted p-value are reported for three pairwise comparisons: Controls vs. At-risk controls (Cluster 4 vs. Cluster 14), At-risk controls vs. AD cases (Cluster 14 vs. Cluster 6) and Controls vs. AD cases (Cluster 4 vs. Cluster 6).

**Data D8. Complete functional enrichment results for pairwise subgroup comparisons.** Enrichment analysis results across Gene Ontology (Biological Process, Cellular Component, Molecular Function), KEGG pathways, Reactome pathways and Disease Ontology are provided in separate sheets for each of the three pairwise comparisons (Cluster 4 vs. Cluster 14, Cluster 14 vs. Cluster 6, Cluster 4 vs. Cluster 6).

### Supplementary Figures

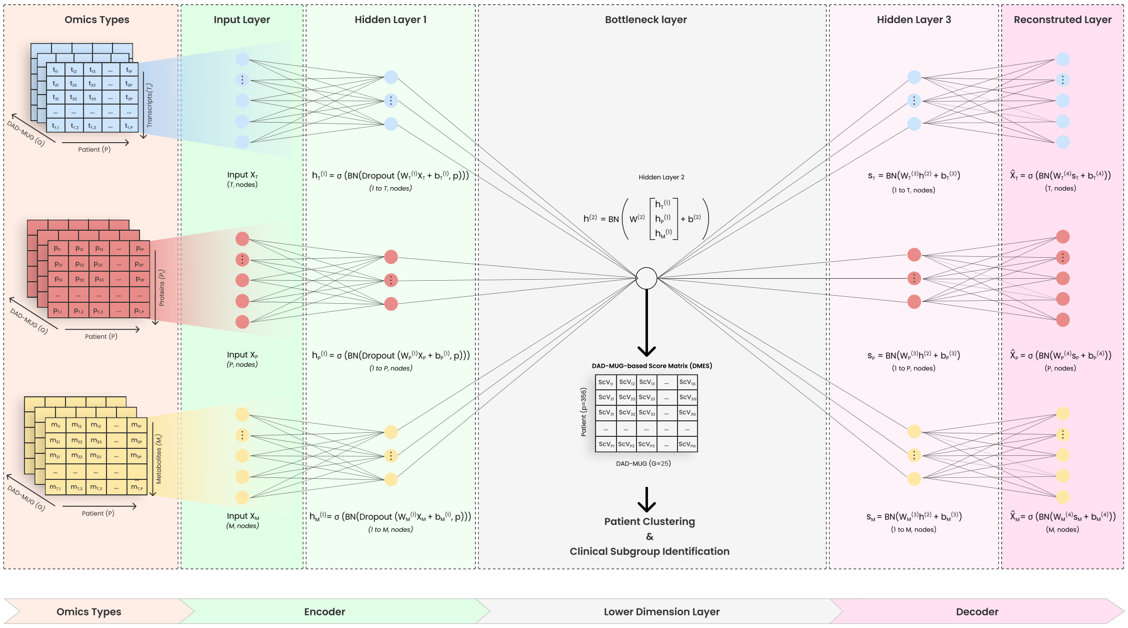

**Figure S1. Autoencoder architecture designed for multi-omics data dimensionality reduction**. The model incorporates three distinct data types: transcriptomics, proteomics  and metabolomics. The network begins with separate input channels for each omics type that feed into the encoder section. In the first hidden layer, each data type is processed independently with tangent activation and batch normalization. These processed signals converge at the bottleneck layer, creating a compressed single-node representation that captures essential biological information. The decoder then expands this compressed signal back through a third hidden layer to reconstruct the original inputs. Upon completion of model training across the 25 DAD-MUGs, we extracted the bottleneck values for each patient-DAD-MUG combination, producing a matrix with dimensions of 25 columns (representing DAD-MUGs) by 356 rows (representing samples). This dimensionally-reduced representation was subsequently utilized for subgroup identification and other downstream analyses.

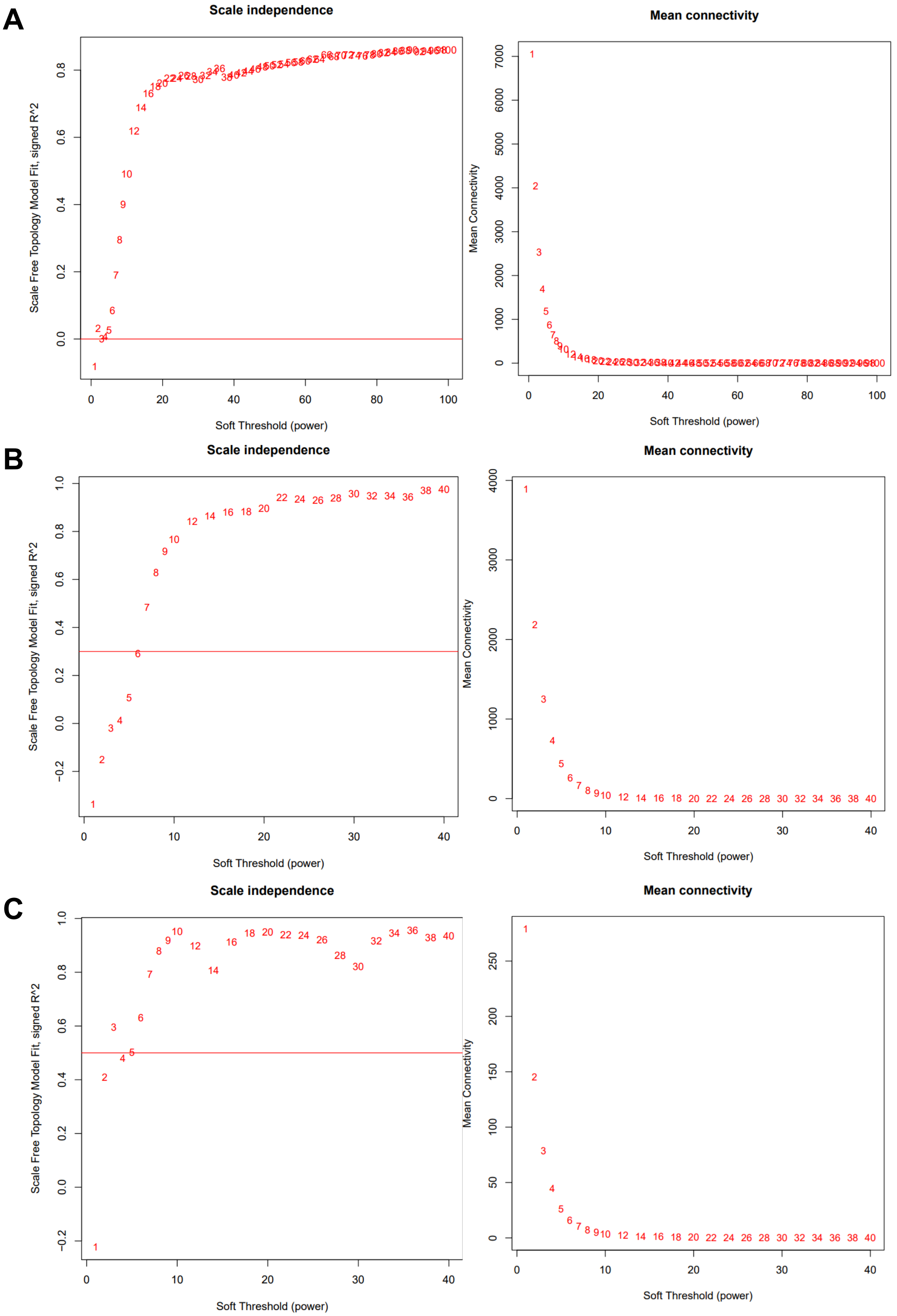
A
Scale independence
Scale Free Topology Model Fit, signed R^2
Soft Threshold (power)
Mean connectivity
Mean Connectivity
Soft Threshold (power)
B
Scale independence
Scale Free Topology Model Fit, signed R^2
Soft Threshold (power)
Mean connectivity
Mean Connectivity
Soft Threshold (power)
C
Scale independence
Scale Free Topology Model Fit, signed R^2
Soft Threshold (power)
Mean connectivity
Mean Connectivity
Soft Threshold (power)

**Figure S2: WGCNA scale-free topology analysis for soft-thresholding power selection.** The left column shows scale independence analysis (Scale Free Topology Model Fit, signed $R^2$ vs. soft-thresholding power) for transcriptomics (A), proteomics (B) and metabolomics (C). The horizontal red lines indicate the $R^2$ thresholds used for power selection (0.0 for transcriptomics, 0.3 for proteomics and 0.5 for metabolomics). The right column shows corresponding mean connectivity analysis at different soft-thresholding powers. Selected powers of 4 (transcriptomics), 7 (proteomics) and 3 (metabolomics) balanced network connectivity with approximate scale-free topology properties. While automated selection suggested higher thresholds, lower powers were chosen to prioritize module assignment and biological interpretability, reducing the number of unassigned features in the grey module.

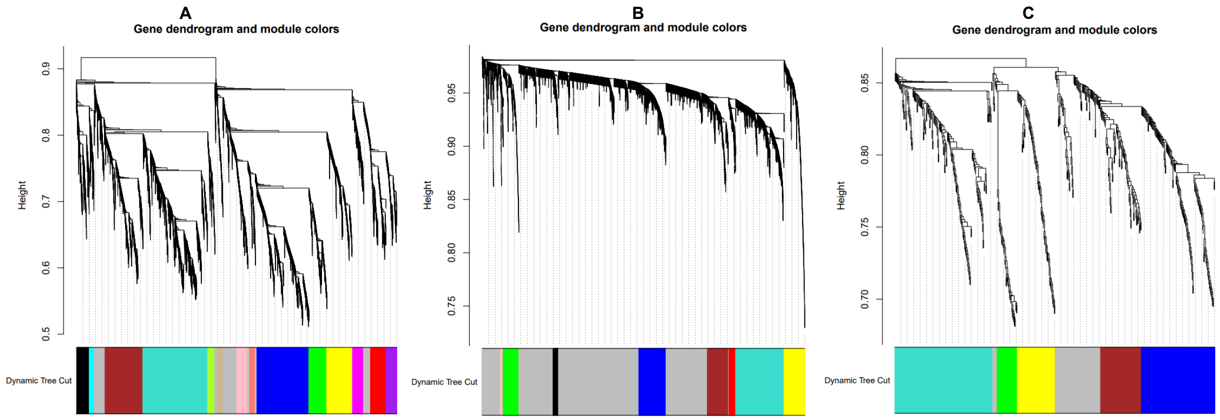

**Figure S3: Hierarchical clustering dendrograms for module identification.** Dendrograms show hierarchical clustering of features based on topological overlap matrices (TOM) derived from adjacency matrices for transcriptomics (A), proteomics (B) and metabolomics (C) datasets. Colored bars below each dendrogram represent module assignments identified through dynamic tree cutting (minimum module size = 30, deep split = 0, using the hybrid method). Each identified module is assigned a unique color, with grey representing unassigned features. The transcriptomics analysis (A) identified 15 modules, proteomics analysis (B) identified 9 modules and metabolomics analysis (C) identified 6 modules.

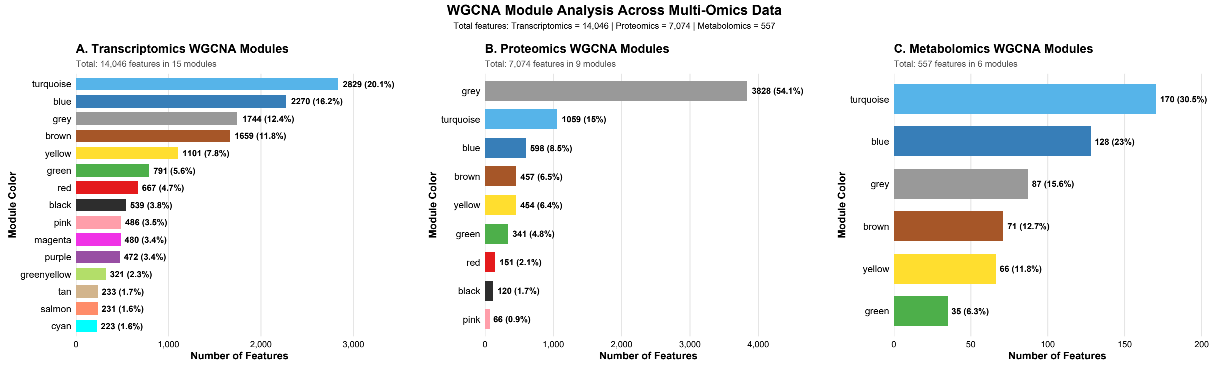

**Figure S4: WGCNA module analysis across multi-omics data.** Bar charts showing the number of features in each module identified by WGCNA analysis. (A) Transcriptomics modules: 14,046 genes distributed across 15 modules, with turquoise (2,829 genes, 20.1%) and blue (2,270 genes, 16.2%) modules containing the largest number of genes. (B) Proteomics modules: 7,074 proteins distributed across 9 modules, with the grey module containing the majority (3,828 proteins, 54.1%). (C) Metabolomics modules: 557 metabolites distributed across 6 modules, with the turquoise module containing the largest number (170 metabolites, 30.5%). Bars are colored according to the standard WGCNA module color assignment.

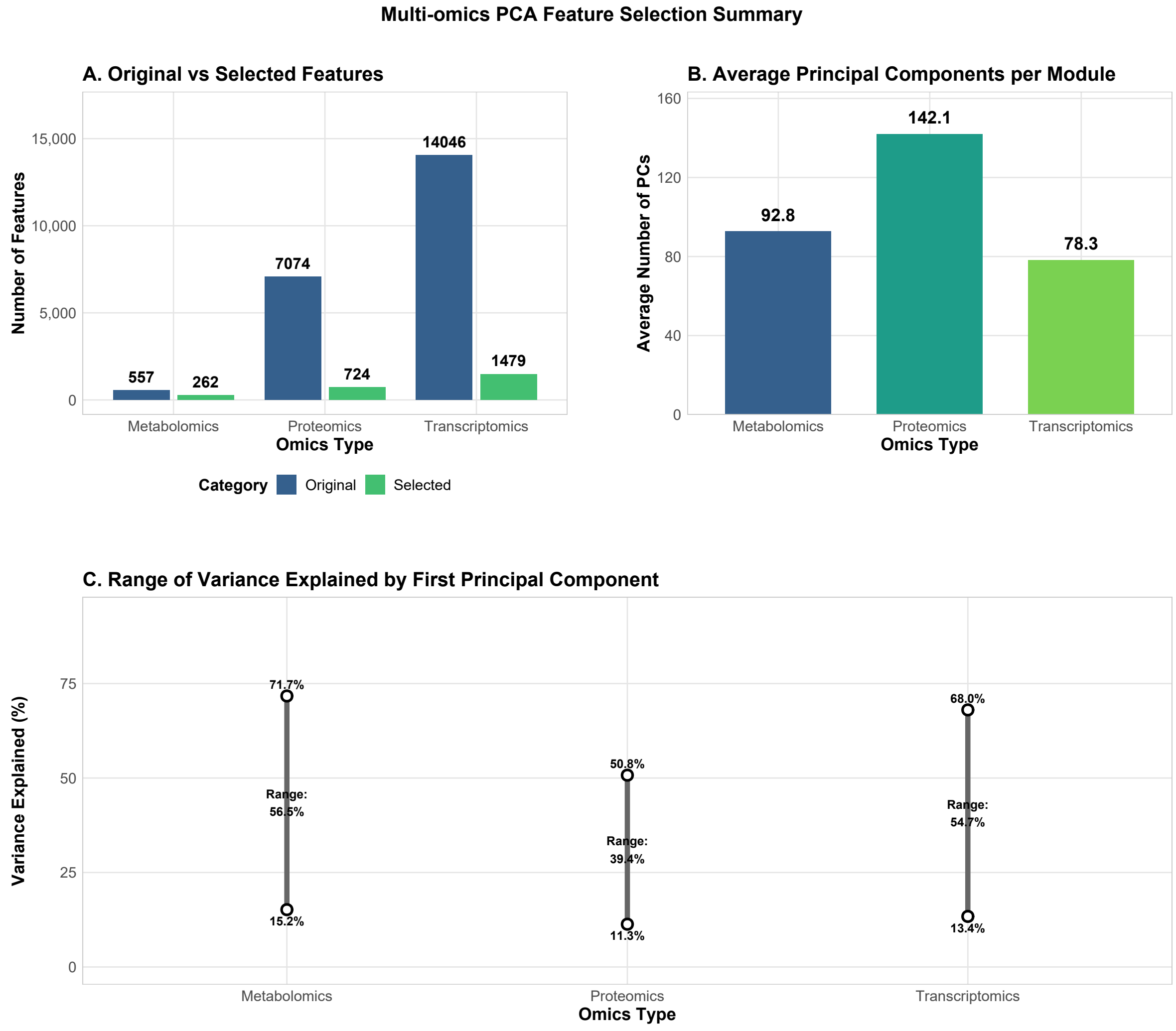

**Figure S5: Summary of PCA-Based feature selection in multi-omics data.** (A) Comparison of original feature counts versus selected features across omics types. Original feature counts were 14,046 genes (transcriptomics), 7,074 proteins (proteomics) and 557 metabolites (metabolomics), from which 1,479, 724 and 262 representative features were selected, respectively. (B) Average number of principal components (PCs) per module for each omics type. Proteomics modules required the highest number of PCs (142.1), followed by metabolomics (92.8) and transcriptomics (78.3). (C) Range of variance explained by the first principal component (PC1) across modules. Each line represents the span from minimum to maximum variance explained by PC1 in modules of each omics type. Metabolomics showed the widest range (56.5%), from 15.2% to 71.7%, followed by transcriptomics (54.7%, from 13.4% to 68.0%) and proteomics (39.4%, from 11.3% to 50.8%).

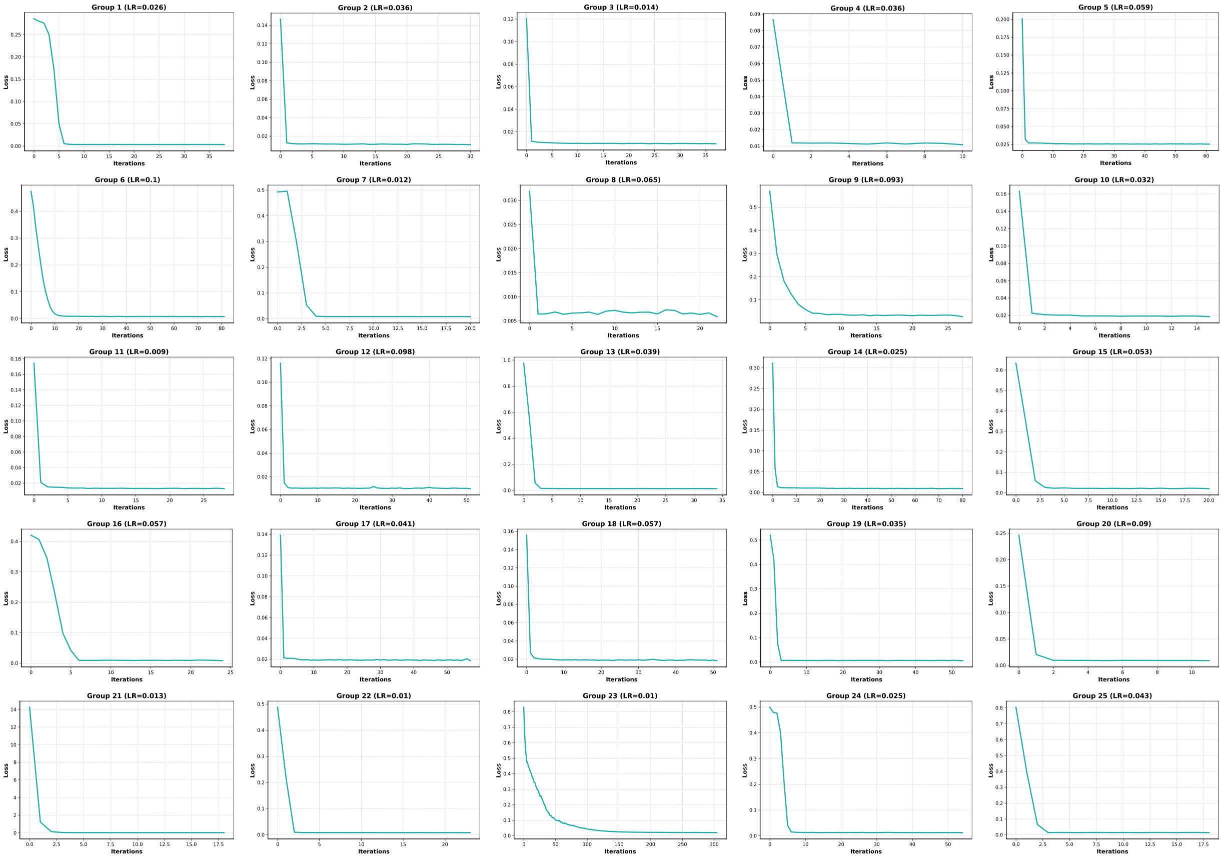

**Figure S6. Training loss trajectories for multi-omics autoencoder across DAD-MUGs.** The plots illustrate the progression of loss function values throughout the autoencoder training procedure for all 25 DAD-MUGs (Groups 1-25 in the figure refer to individual DAD-MUG). The horizontal axis represents training epochs while the vertical axis displays the composite loss values incorporating reconstruction error and regularization components. During each epoch, model parameters are optimized using the complete multi-omics dataset for the respective DAD-MUG. Some DAD-MUGs (e.g., DAD-MUGs 4, 10) complete training within 20 epochs due to early stopping activation, where the loss function achieves its optimal value and remains unchanged for 20 subsequent epochs. Specifically, DAD-MUG 4 attains minimal loss around epoch 2, and since no further improvement occurs over the following 20 epochs, the visualization displays only the initial 10 epochs of meaningful optimization.

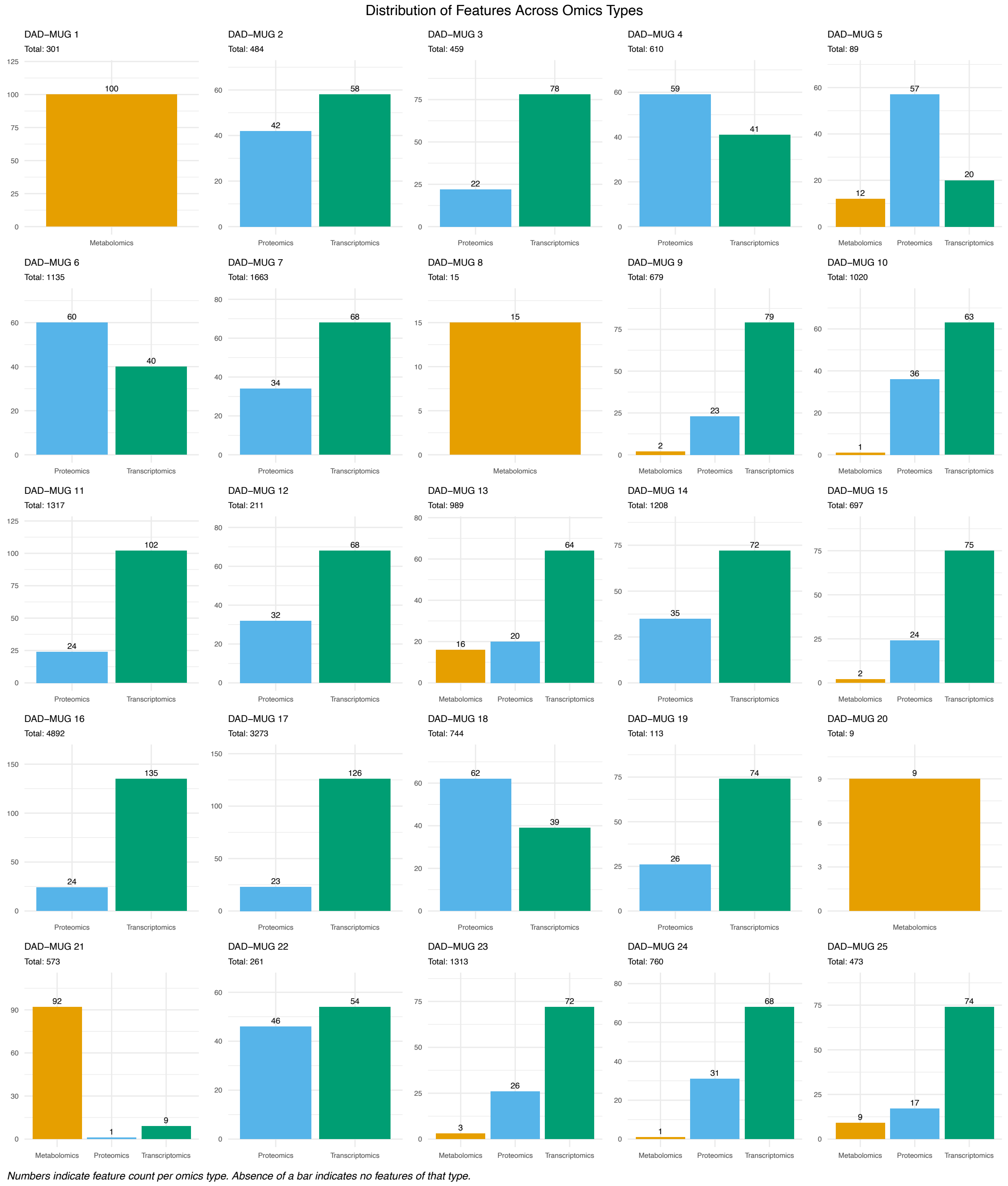

**Figure S7: Absolute feature counts distributed across omics types for each DAD-MUG.** Bar charts display the number of features from each omics type (metabolomics, proteomics and transcriptomics) allocated to each of the 25 DAD-MUGs. The total number of features originally available in each group is shown below each DAD-MUG number.

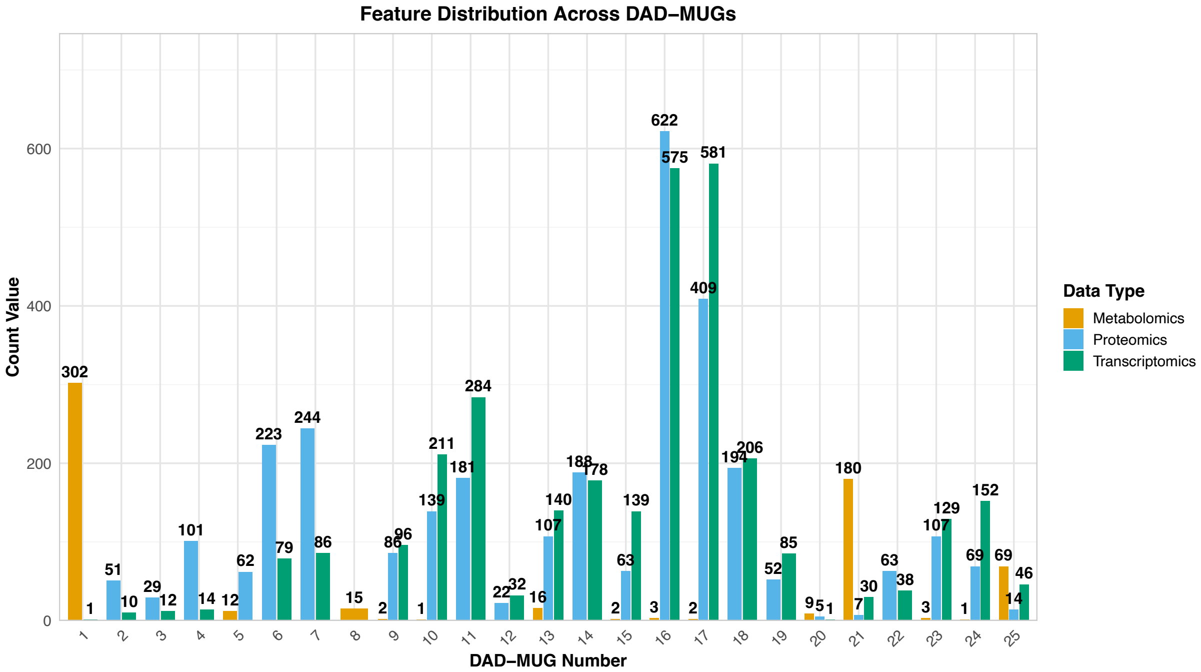

**Figure S8: Distribution of features across the 25 DAD-MUGs using the initial unbalanced approach.** The stacked bar plot displays transcriptomics, proteomics and metabolomics features per DAD-MUG, showing a markedly unbalanced distribution. Some DAD-MUGs contain many features (e.g., 1200 total features in DAD-MUG 16, 992 in DAD-MUG 17, 366 in DAD-MUG 14), while others have substantially fewer (e.g., 61 features in DAD-MUG 2, 41 in DAD-MUG 3, 54 in DAD-MUG 12).

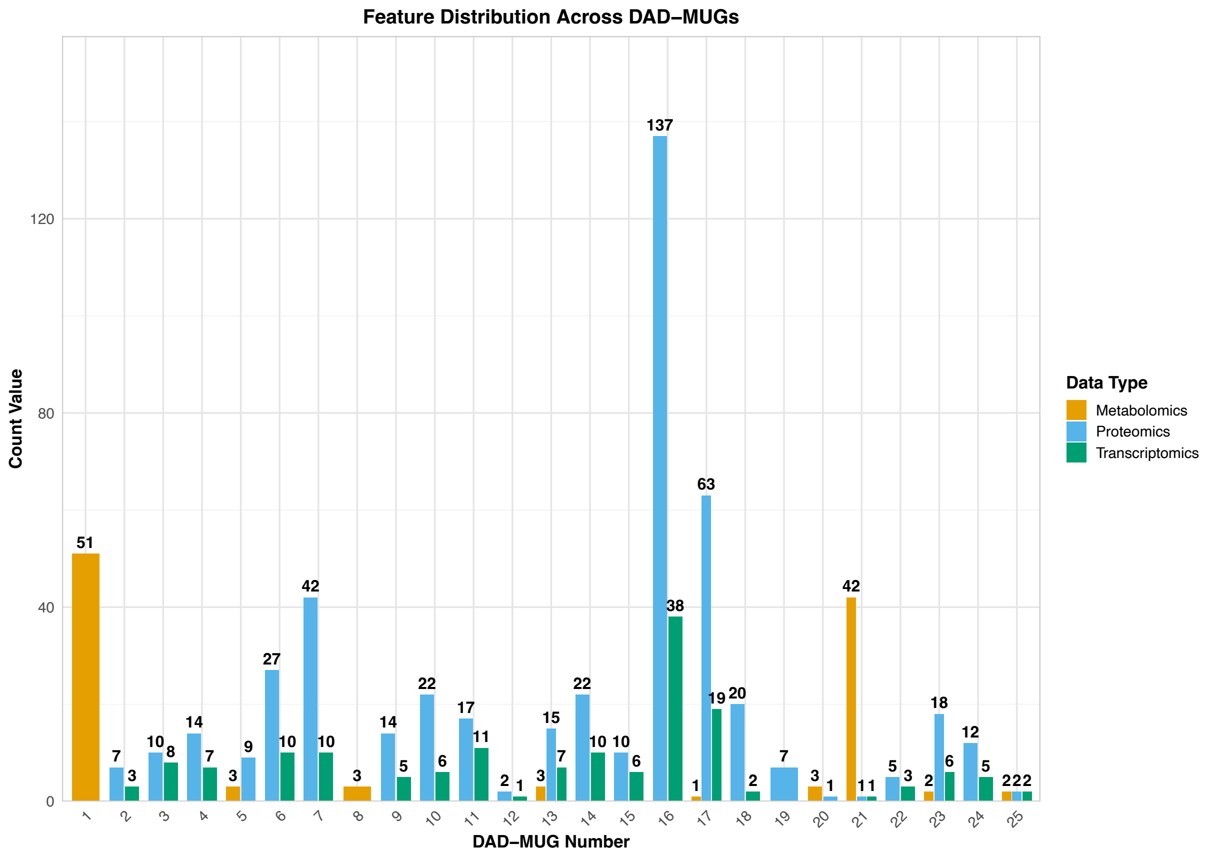

**Figure S9: Distribution of features across the 25 DAD-MUGs using the intermediate feature selection approach.** The stacked bar plot displays transcriptomics, proteomics and metabolomics features per DAD-MUG, showing that even after reducing the total feature count to 747, considerable imbalance remains. Some DAD-MUGs contain many features (e.g., 175 total features in DAD-MUG 16, 83 in DAD-MUG 17), while others have substantially fewer (e.g., 6 features in DAD-MUG 25, 10 in DAD-MUG 2).

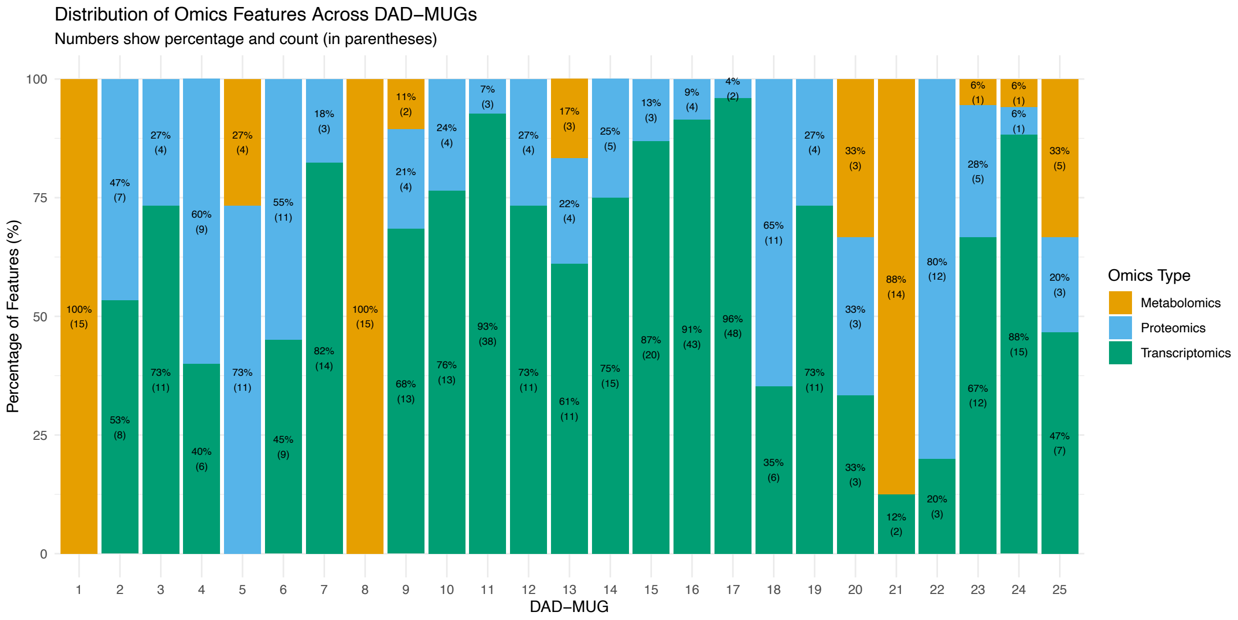

**Figure S10: Distribution of omics features across the 25 DAD-MUGS using the compact balanced approach.** Stacked bar plot shows the percentage and count of transcriptomics, proteomics, and metabolomics features per DAD-MUG, illustrating the more proportional distribution achieved through the balancing procedure described in the text.

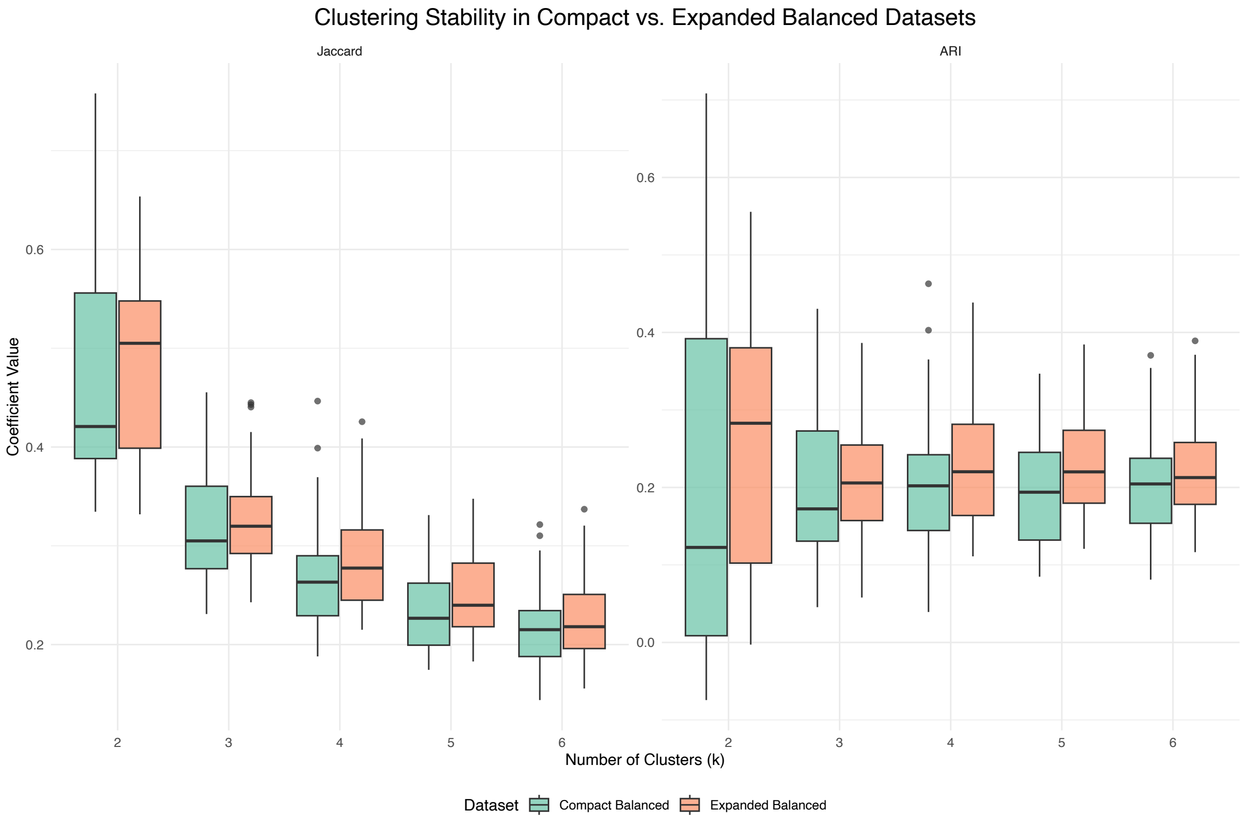

**Figure S11: Comparison of clustering consistency between balanced feature selection approaches.** Boxplots show the distribution of Jaccard similarity coefficients (left) and Adjusted Rand Index values (right) from pairwise comparisons among 10 different DMES matrices for each approach. The expanded balanced approach (containing 2,465 features) demonstrated superior clustering consistency compared to the compact balanced approach (containing approximately 499 features), particularly for K=4, K=5, and K=6. Each boxplot represents 45 pairwise comparisons within a single dataset for a specific cluster number (K).

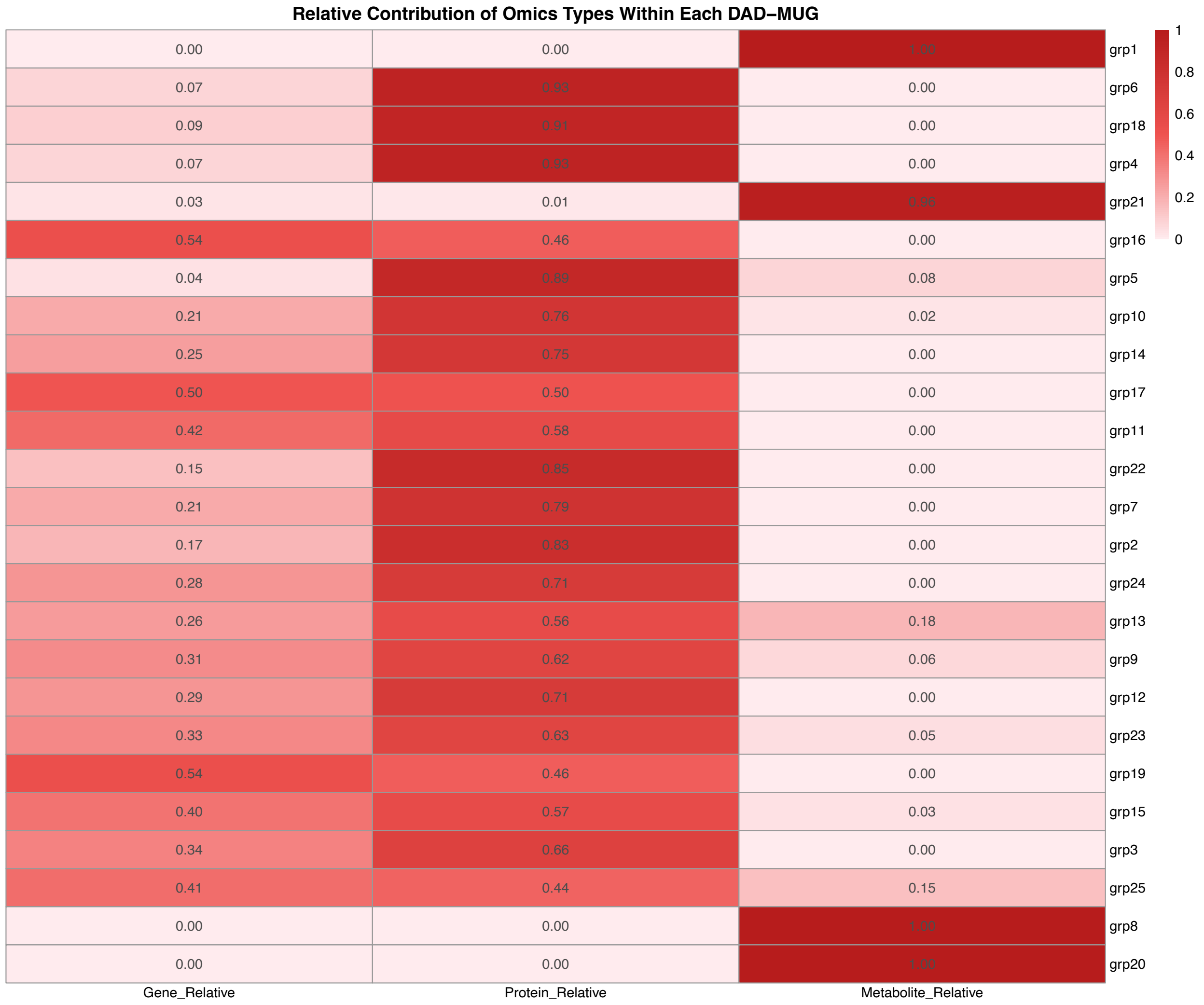

**Figure S12. Heatmap of relative omics contributions to DAD-MUGs.** Heatmap displaying the relative contributions of genes, proteins and metabolites to each of the 25 DAD-MUGs, shown as proportions ranging from 0 to 1. Color intensity corresponds to contribution magnitude, with values displayed in each cell. A value of 0 indicates the absence of that omics type in the respective DAD-MUG, while 1 indicates that omics type comprises the entire DAD-MUG.

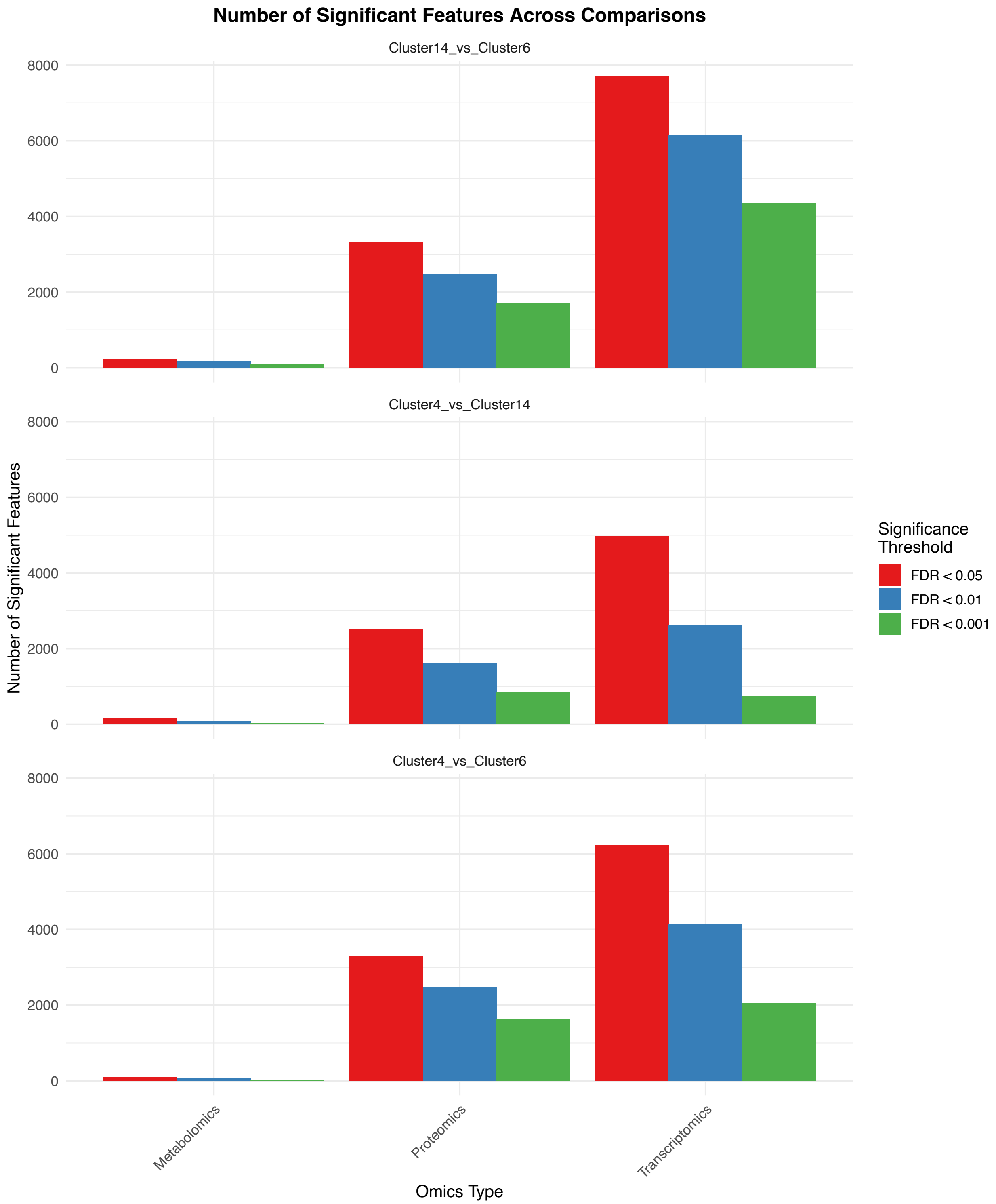

**Figure S13. Number of Significant Features Across Molecular Subgroup Comparisons.** Bar plot showing the distribution of significantly dysregulated features (FDR <0.05, < 0.01 and < 0.001) across three pairwise comparisons and three omics types. The disease transition stage (Cluster 14 vs Cluster 6) exhibited the highest number of dysregulated features across all omics types, reflecting extensive molecular remodeling during progression from at-risk to disease state. Transcriptomics showed the largest number of significant features in all comparisons, followed by proteomics and metabolomics. Color coding: red = FDR < 0.05, blue = FDR < 0.01, green = FDR < 0.001.

### Captions for supplementary figures provided separately

**Figure S14. Functional enrichment analysis of the top 50 discriminative features.** Enrichment results for the 50 highest-ranked features (by Gini importance from the weighted Random Forest model) across Gene Ontology Biological Process, Cellular Component, Molecular Function, KEGG pathways, Reactome pathways and Disease Ontology.

**Figure S15. Functional enrichment analysis of all 500 discriminative features.** Enrichment results for the complete set of 500 RFE-RF selected features across Gene Ontology Biological Process, Cellular Component, Molecular Function, KEGG pathways, Reactome pathways and Disease Ontology.

**Figure S16. Functional enrichment analysis for Controls vs. At-risk controls (Cluster 4 vs. Cluster 14).** Over-representation analysis results for significantly dysregulated genes and proteins across Gene Ontology Biological Process (GO-BP), Cellular Component (GO-CC), Molecular Function (GO-MF), KEGG pathways, Reactome pathways and Disease Ontology.

**Figure S17. Functional enrichment analysis for At-risk controls vs. AD cases (Cluster 14 vs. Cluster 6).** Over-representation analysis results for significantly dysregulated genes and proteins across Gene Ontology Biological Process (GO-BP), Cellular Component (GO-CC), Molecular Function (GO-MF), KEGG pathways and Reactome pathways.

**Figure S18. Functional enrichment analysis for Controls vs. AD cases (Cluster 4 vs. Cluster 6).** Over-representation analysis results for significantly dysregulated genes and proteins across Gene Ontology Biological Process (GO-BP), Cellular Component (GO-CC), Molecular Function (GO-MF), KEGG pathways and Reactome pathways.

### Supplementary Algorithm

#### Algorithm: Multi-Omics Feature Selection and Balancing

##### Notation

**Sets and Indices:**

- $F$: Set of all features across three omics types
- $G = \{1, 2, \ldots, 25\}$: Set of biological groups (DAD-MUGs)
- $O = \{T, P, M\}$: Set of omics types (Transcriptomics, Proteomics, Metabolomics)
- $M(o)$: Set of modules for omics type $o$, where $o \in O$
- $PC(i)$: The $i$-th principal component, where $i = 1, \ldots, N$
- $N$: Last PC meeting selection criteria

**Functions and Parameters:**

- $\rho(f, PC(i), m)$: Correlation of feature $f$ with $PC(i)$ in module $m$
- $|\rho(f, PC(i), m)|$: Absolute correlation value
- $g(f)$: Group assignment function mapping feature $f$ to group number in $G$
- $T(g)$: Target number of features for group $g$
- $c(g)$: Current count of selected features in group $g$

**Algorithm Constants:**

- $S$: Set of selected features, where $|S| = 2465$
- $n_1 = 2313$: Phase 1 target (total features)
- $n_2 = 152$: Phase 2 target (additional features)

##### Target Allocation

The target $T(g)$ for each group $g$ is defined as:

$$T(g) = \begin{cases} 9, & \text{if } g = 20 \\ 15, & \text{if } g = 8 \\ 89, & \text{if } g = 5 \\ 100, & \text{otherwise} \end{cases} \tag{1}$$

where $\sum_{g=1}^{25} T(g) = 2313$.

##### Algorithm

**Input:**

- Correlation matrix $C = \{\rho(f, PC(i), m) : f \in F, i \in [1, N], m \in M(o), o \in O\}$
- Group assignments $g(f)$ for all $f \in F$

**Output:**

- $S \subset F$ with $|S| = 2465$

**Algorithm 1:** Multi-Omics Feature Selection with Group Balancing

**Initialize:** $S \leftarrow \emptyset$, $c(g) \leftarrow 0$ for all $g \in G$;

// Phase 1: Balanced Selection ($n_1 = 2313$ features)

**for** $i = 1$ **to** $N$ **do**

    **if** $\sum_{g \in G} c(g) \geq n_1$ **then**

        **break**;

    $F_i \leftarrow \{f \in F \setminus S : f$ is associated with $PC(i)\}$;

    **while** $F_i \neq \emptyset$ ***and*** $\sum_{g \in G} c(g) < n_1$ **do**

        // Step 1: Random omics selection

        $o^* \leftarrow$ Random sample from $\{o \in O : \exists m \in M(o), \exists f \in F_i$ associated with $(f, m, o)\}$;

        // Step 2: Random module selection

        $m^* \leftarrow$ Random sample from $\{m \in M(o^*) : \exists f \in F_i$ associated with $(f, m, o^*)\}$;

        // Step 3: Correlation-ordered candidate selection

        $F_{i,m^*} \leftarrow \{f \in F_i : f$ is associated with $(PC(i), m^*, o^*)\}$;

        Sort $F_{i,m^*}$ by $|\rho(f, PC(i), m^*)|$ in descending order;

        // Step 4: Feature assignment with group constraint

        **foreach** $f \in F_{i,m^*}$ **do**

            **if** $c(g(f)) < T(g(f))$ **then**

                $S \leftarrow S \cup \{f\}$;

                $c(g(f)) \leftarrow c(g(f)) + 1$;

                **break**;

        // Step 5: Remove processed features

        $F_i \leftarrow F_i \setminus (F_{i,m^*} \cup \{\text{features already in } S\})$;

// Phase 2: Top Remaining Features ($n_2 = 152$ features)

$F_{\text{remaining}} \leftarrow F \setminus S$;

// Sort all remaining features by their maximum absolute correlation

**foreach** $f \in F_{remaining}$ **do**

    $\text{max\_corr}(f) \leftarrow \max_{i,m} |\rho(f, PC(i), m(f))|$;

    where $m(f)$ is the module yielding maximum correlation for $f$;

Sort $F_{\text{remaining}}$ by $\text{max\_corr}(f)$ in descending order;

$S \leftarrow S \cup \{\text{top } n_2 \text{ features from } F_{\text{remaining}}\}$;

**return** $S$
