## Supplementary figures and images for "Balanced deep learning on multi-omics networks identifies molecular subgroups of pathological brain aging"

### Figure S14.pdf

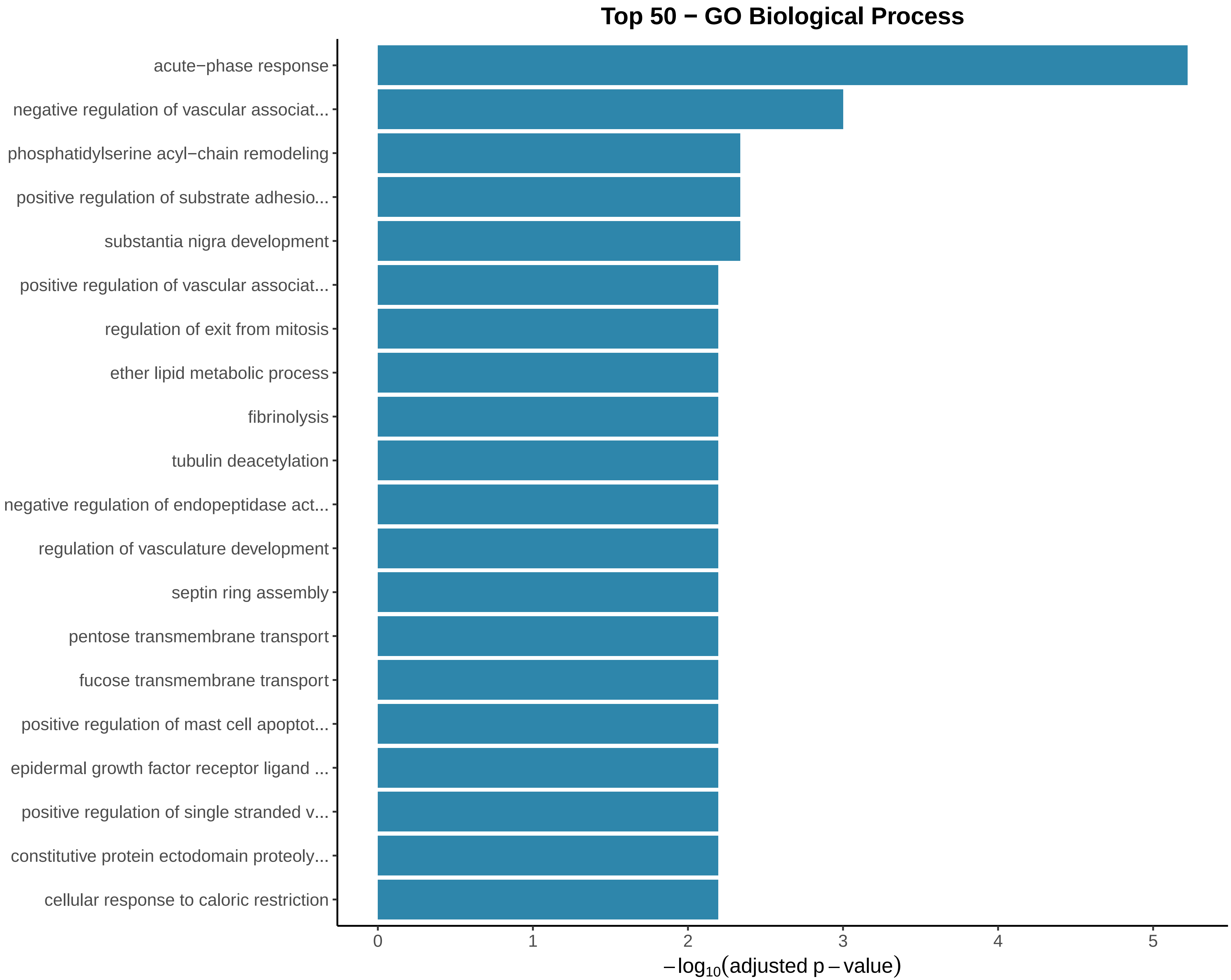

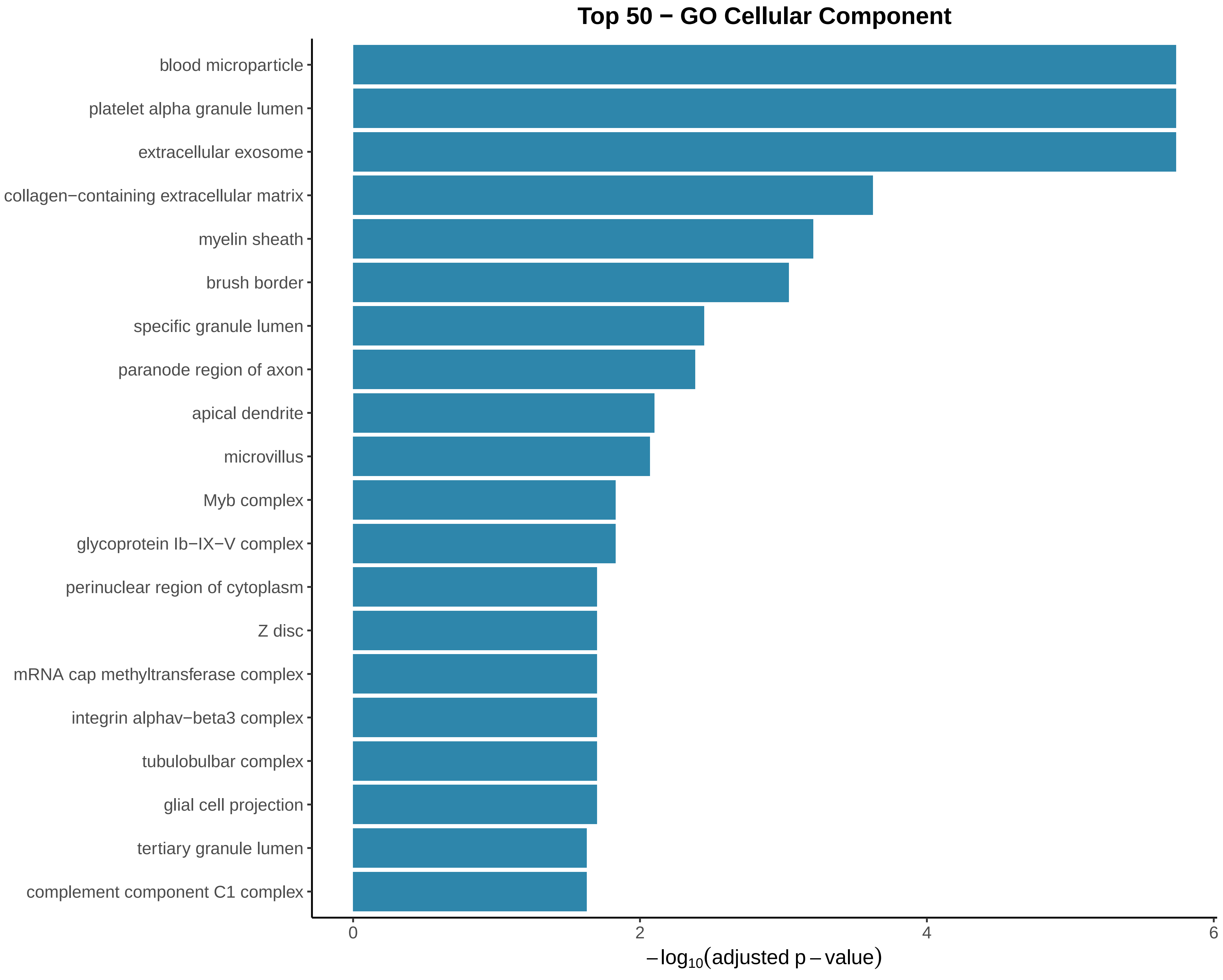

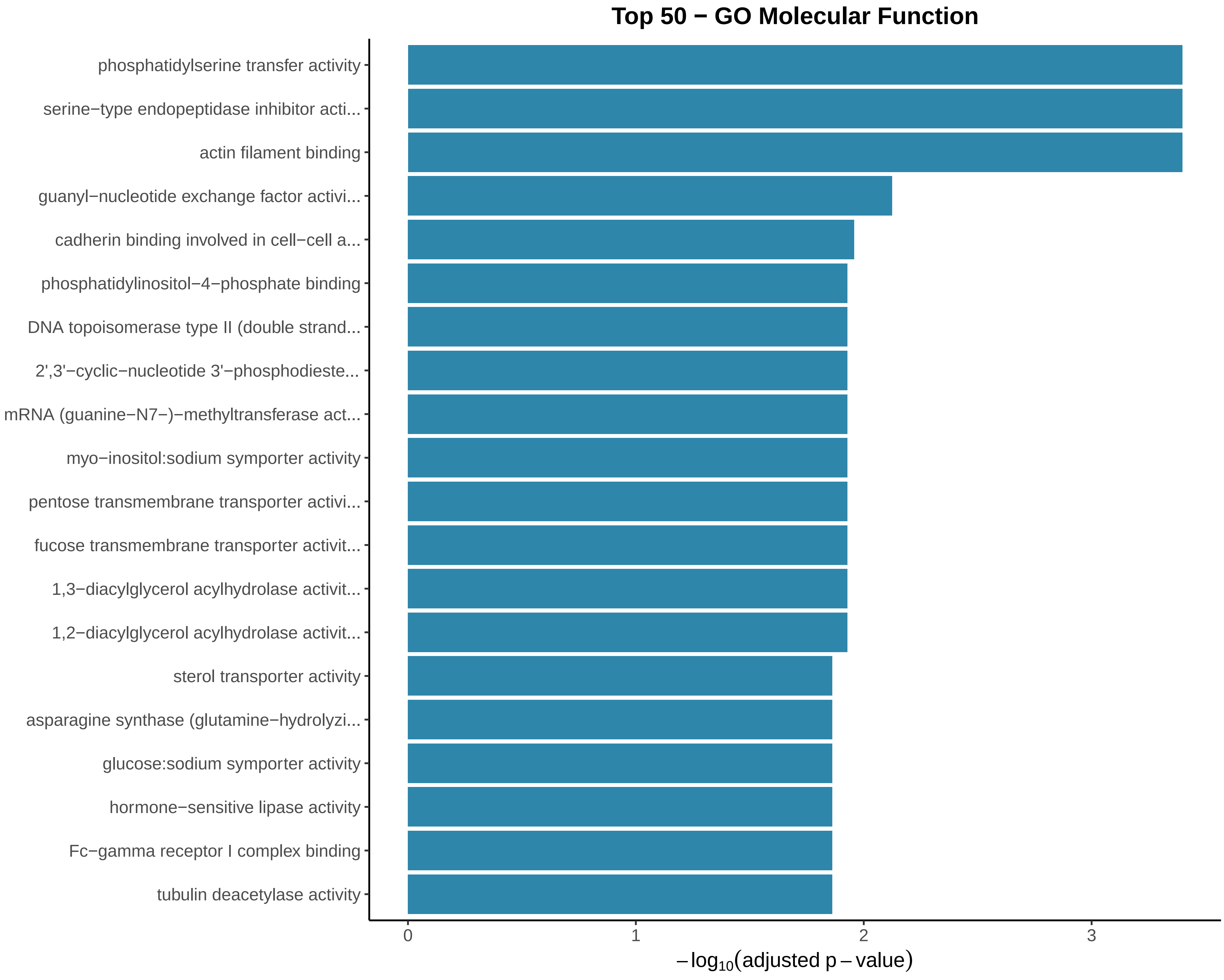

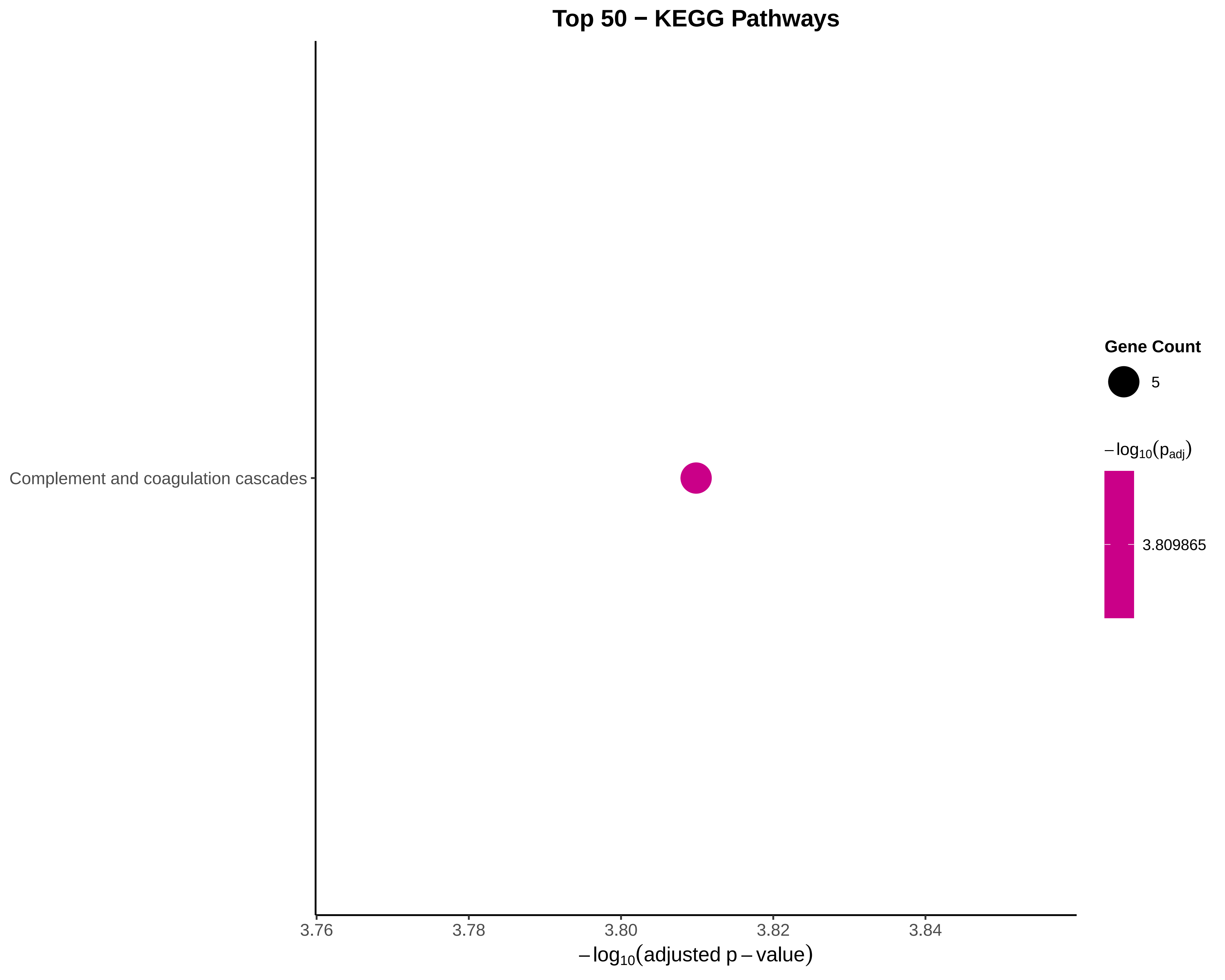

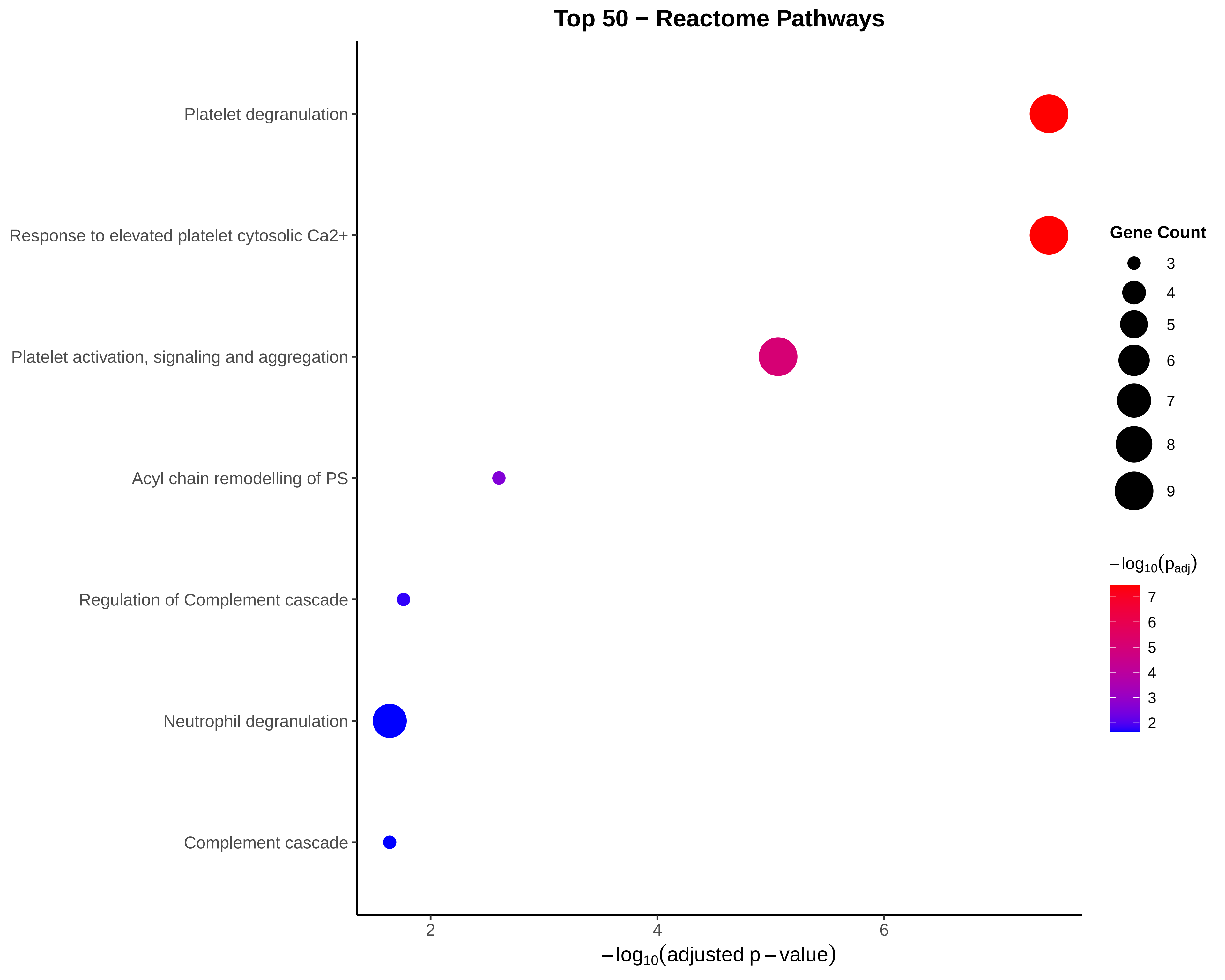

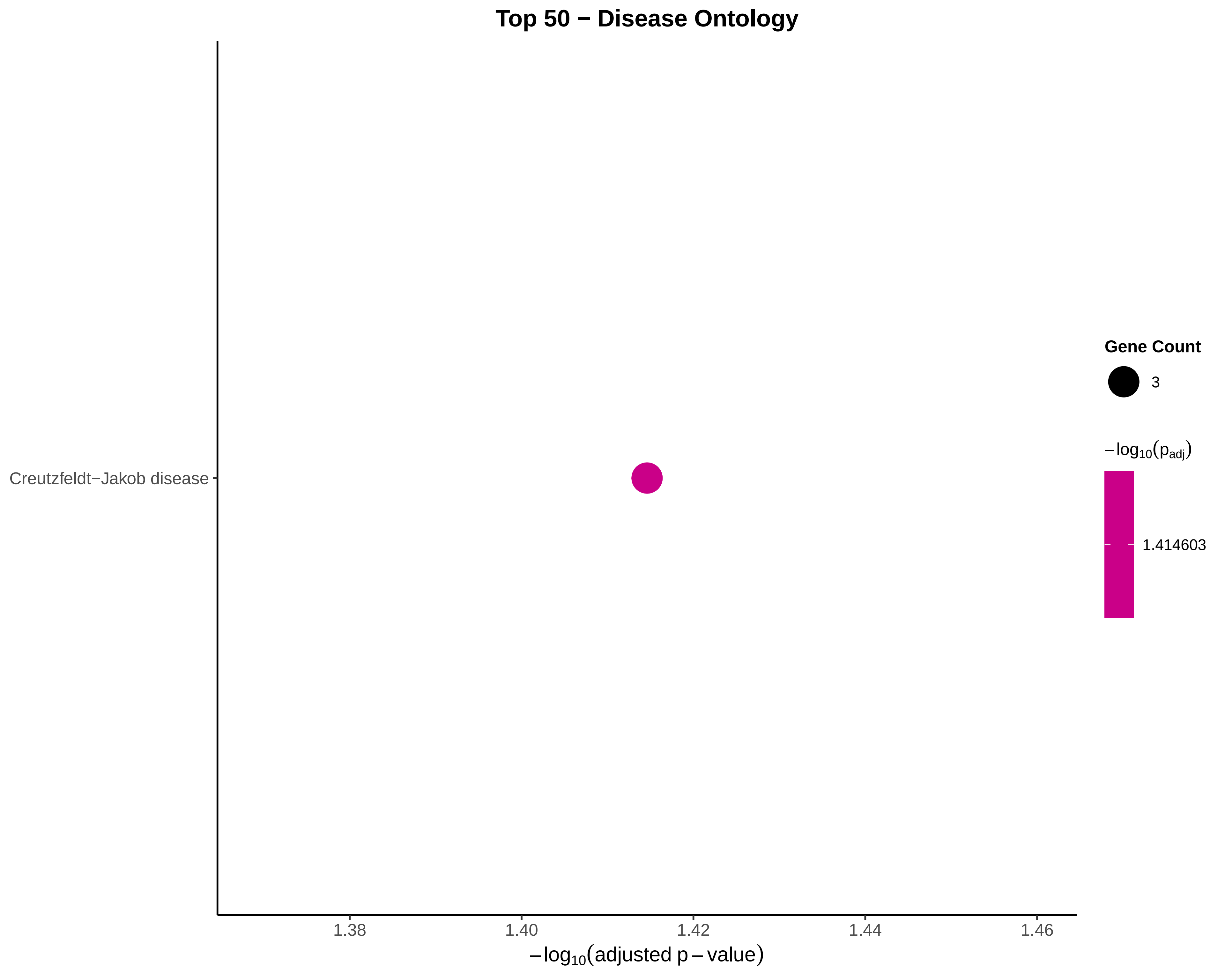

### Figure S15.pdf

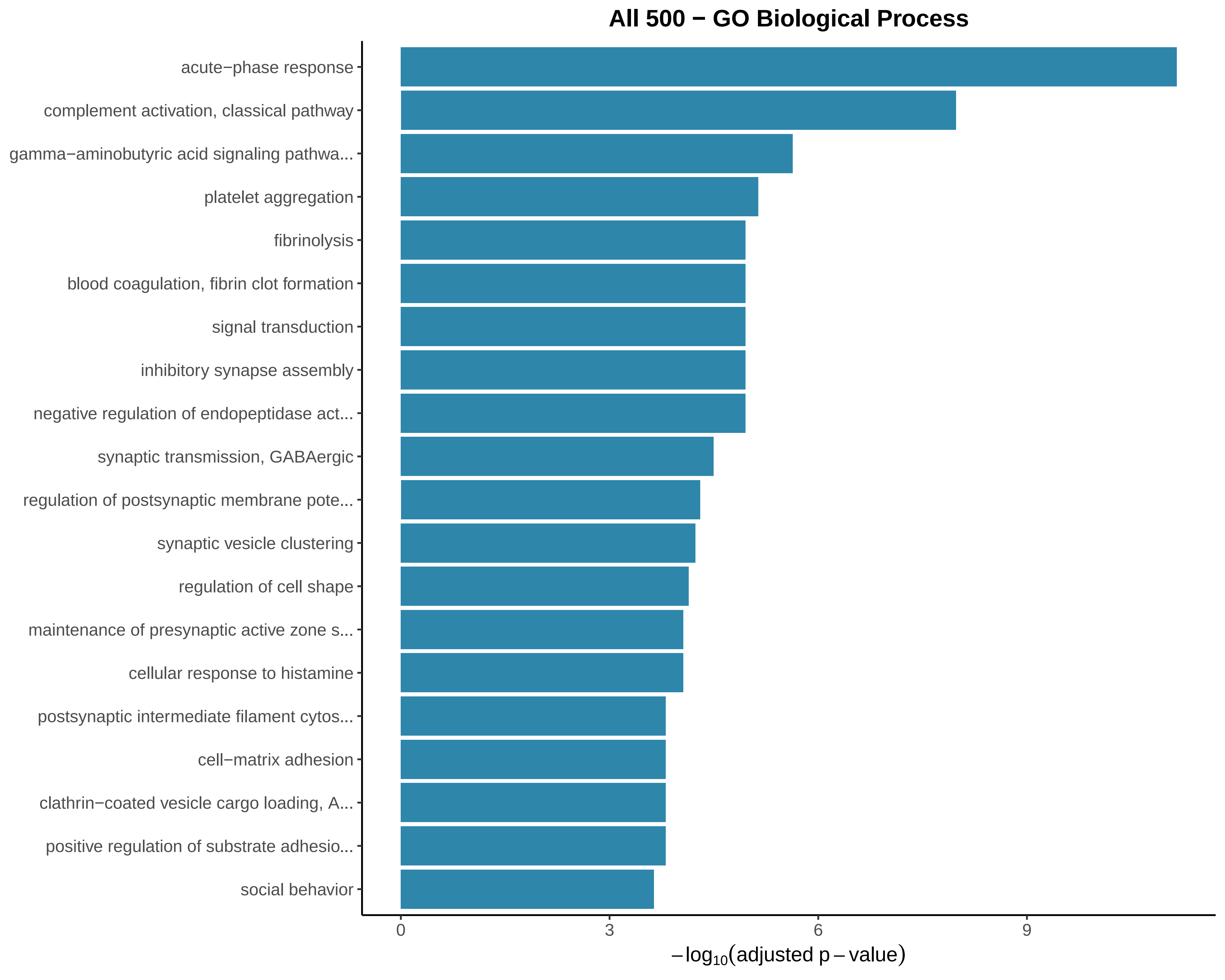

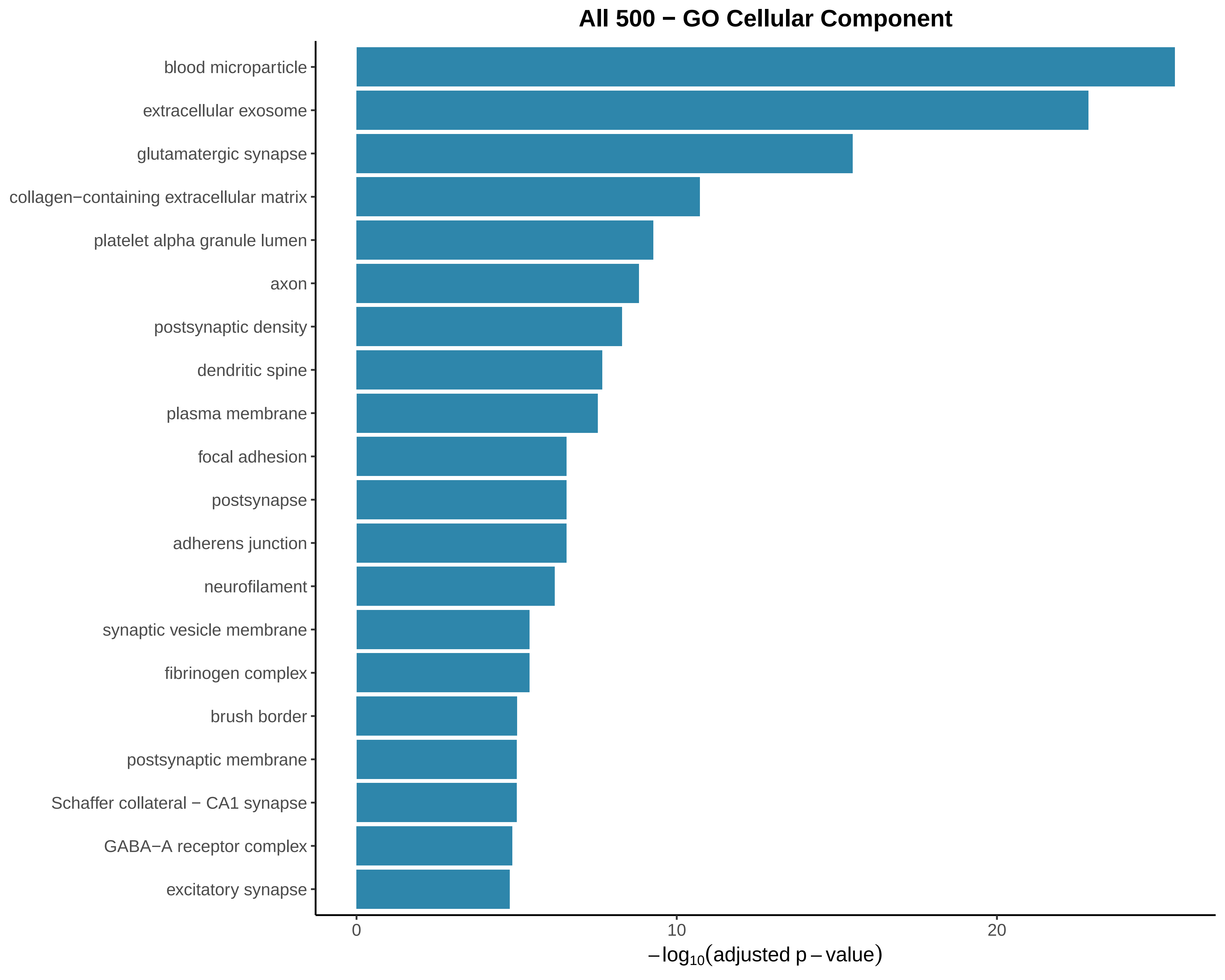

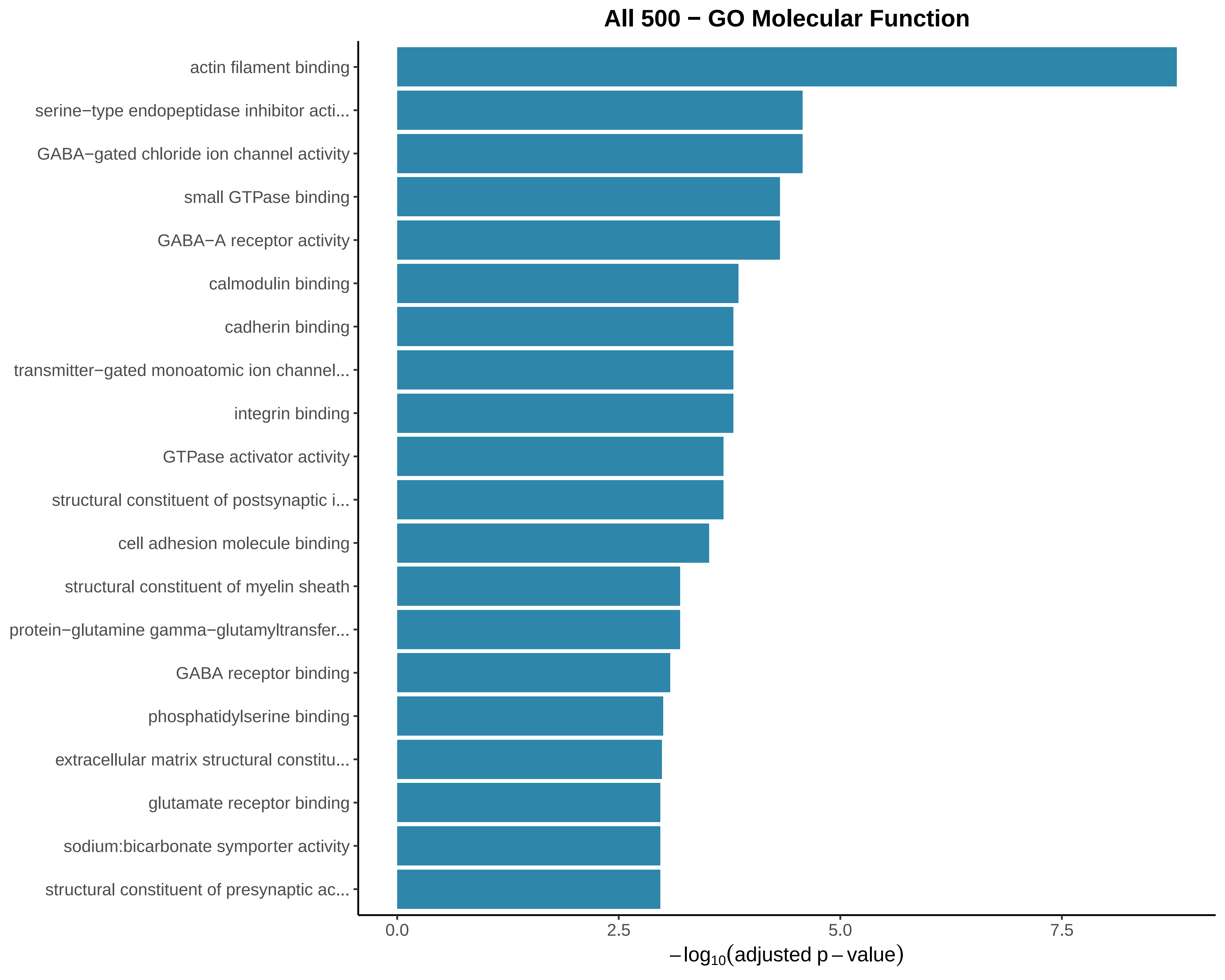

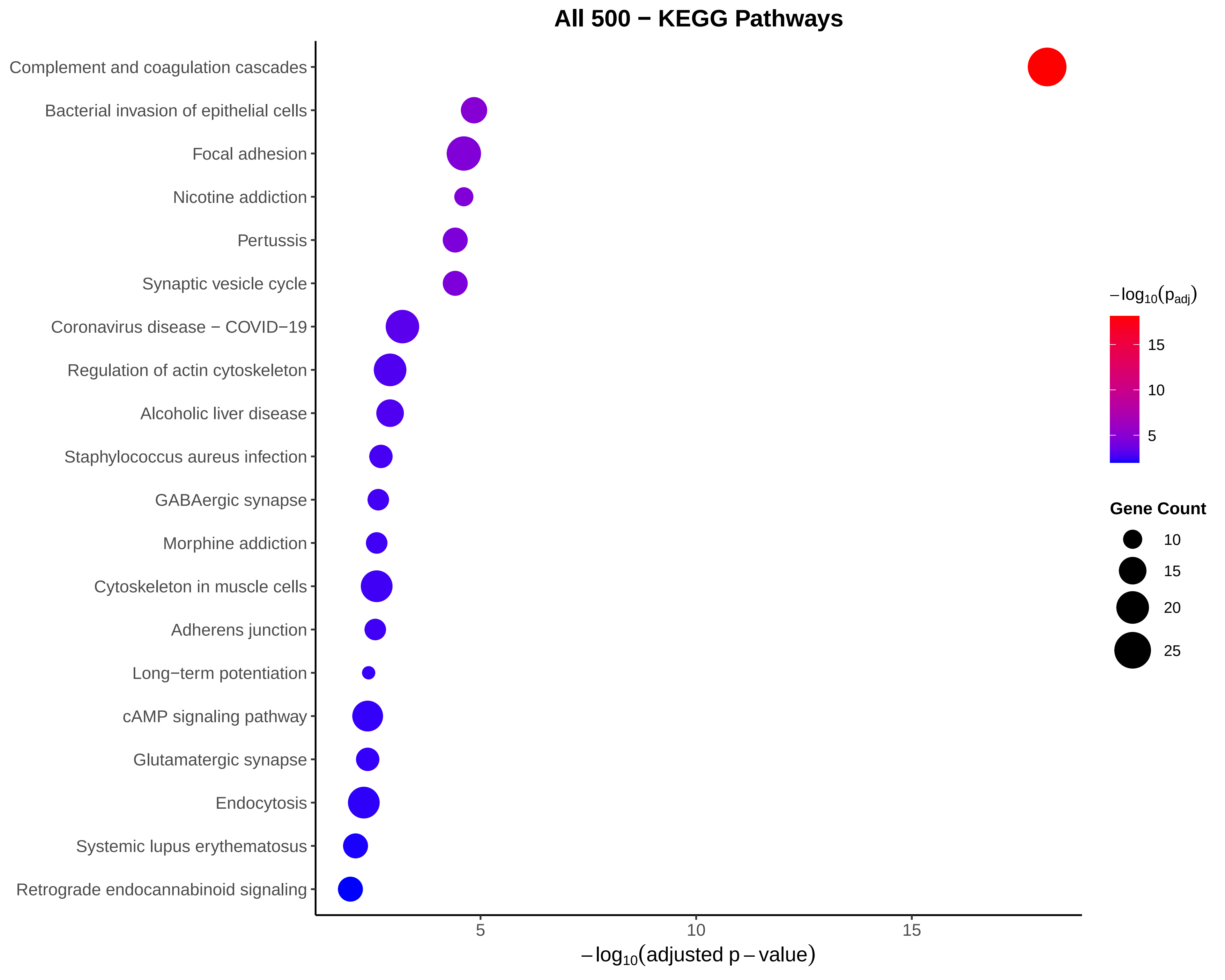

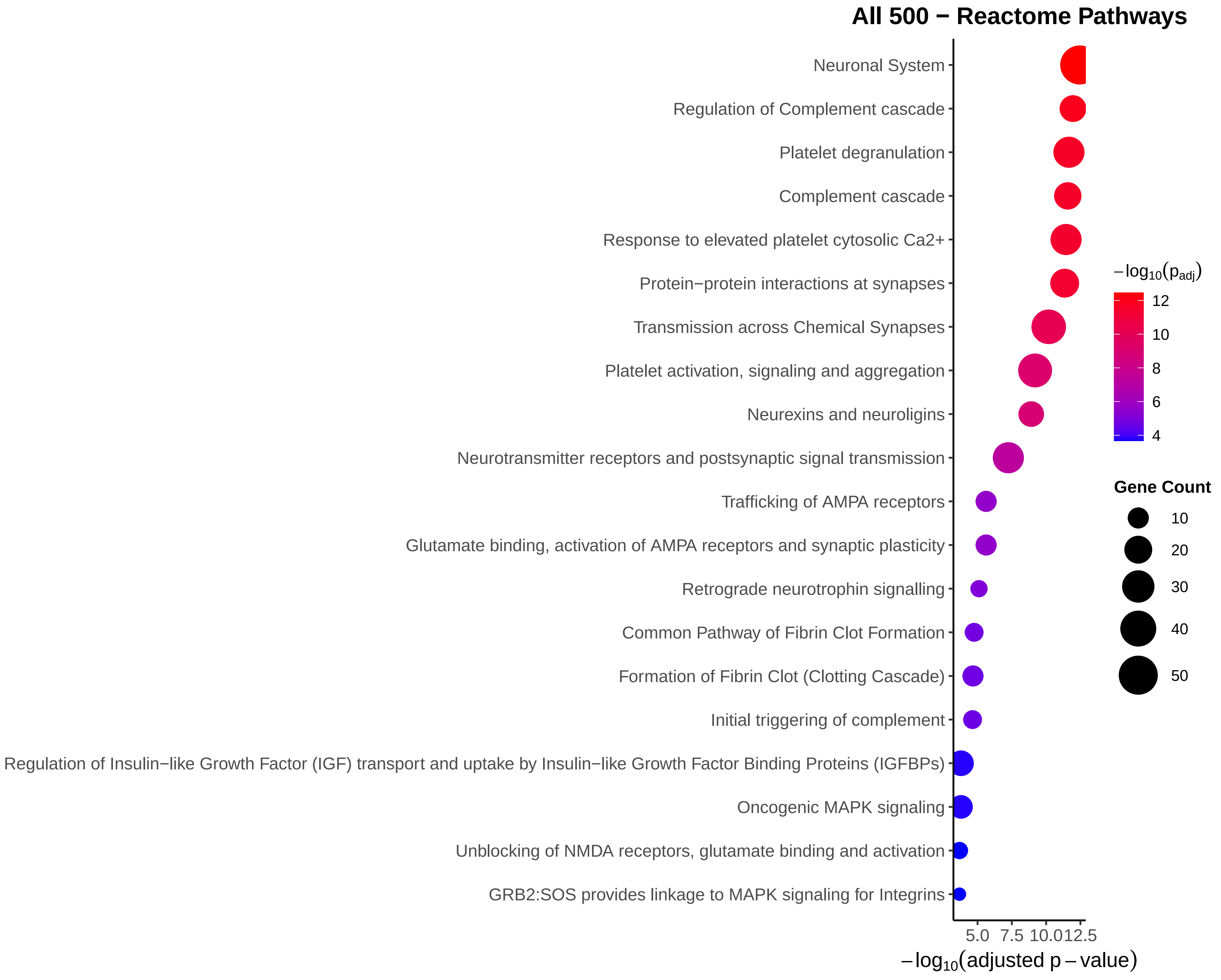

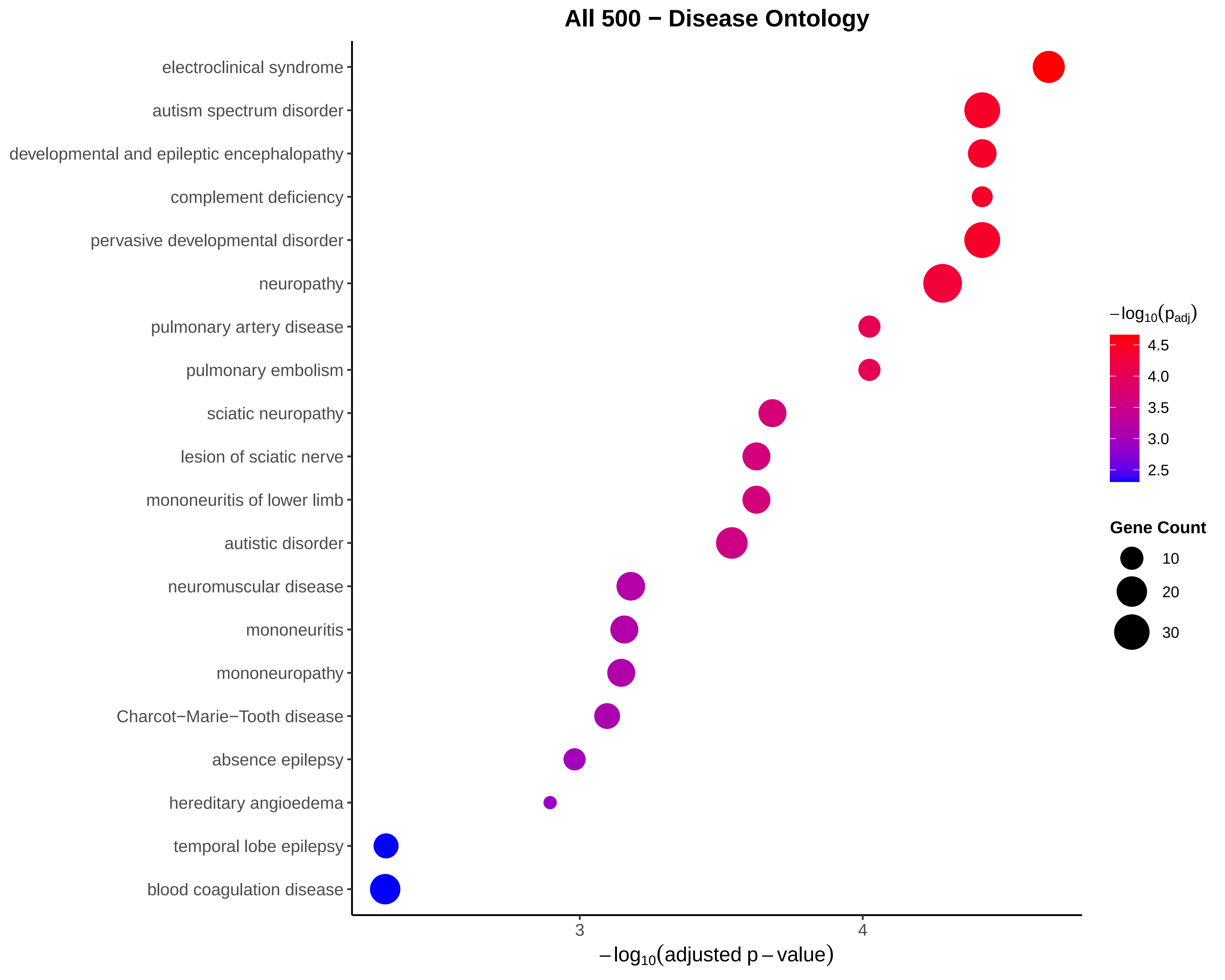

### Figure S16.pdf

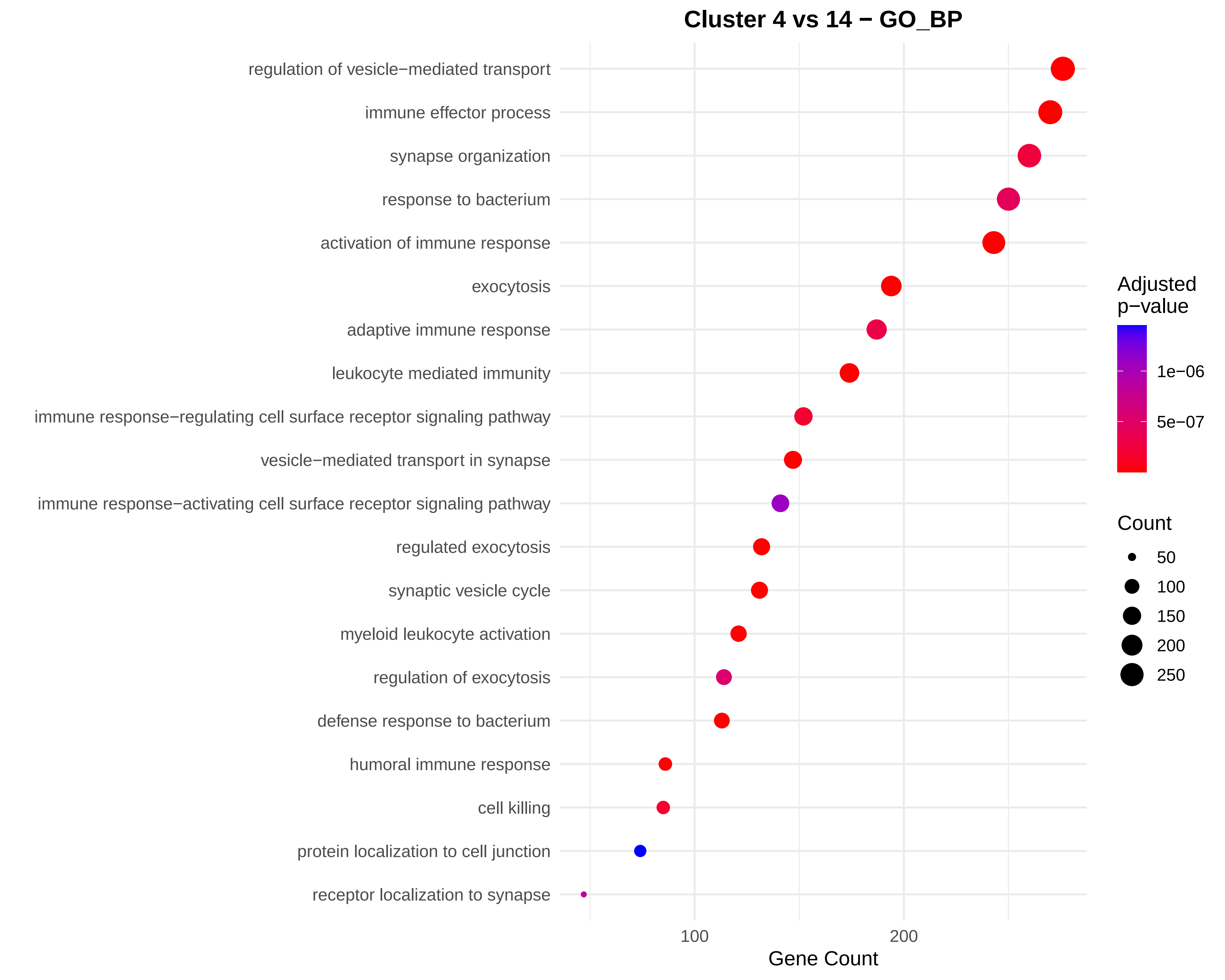

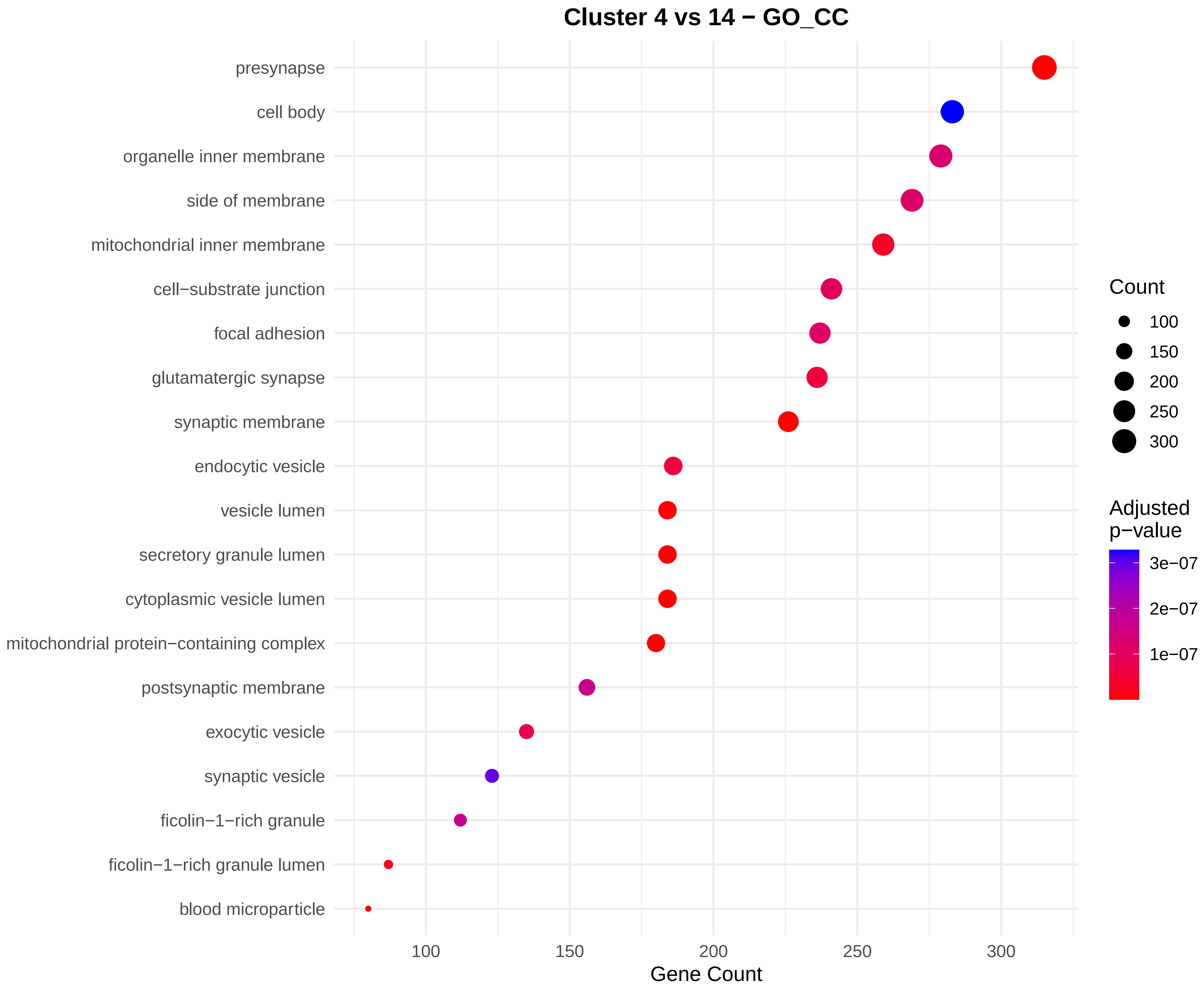

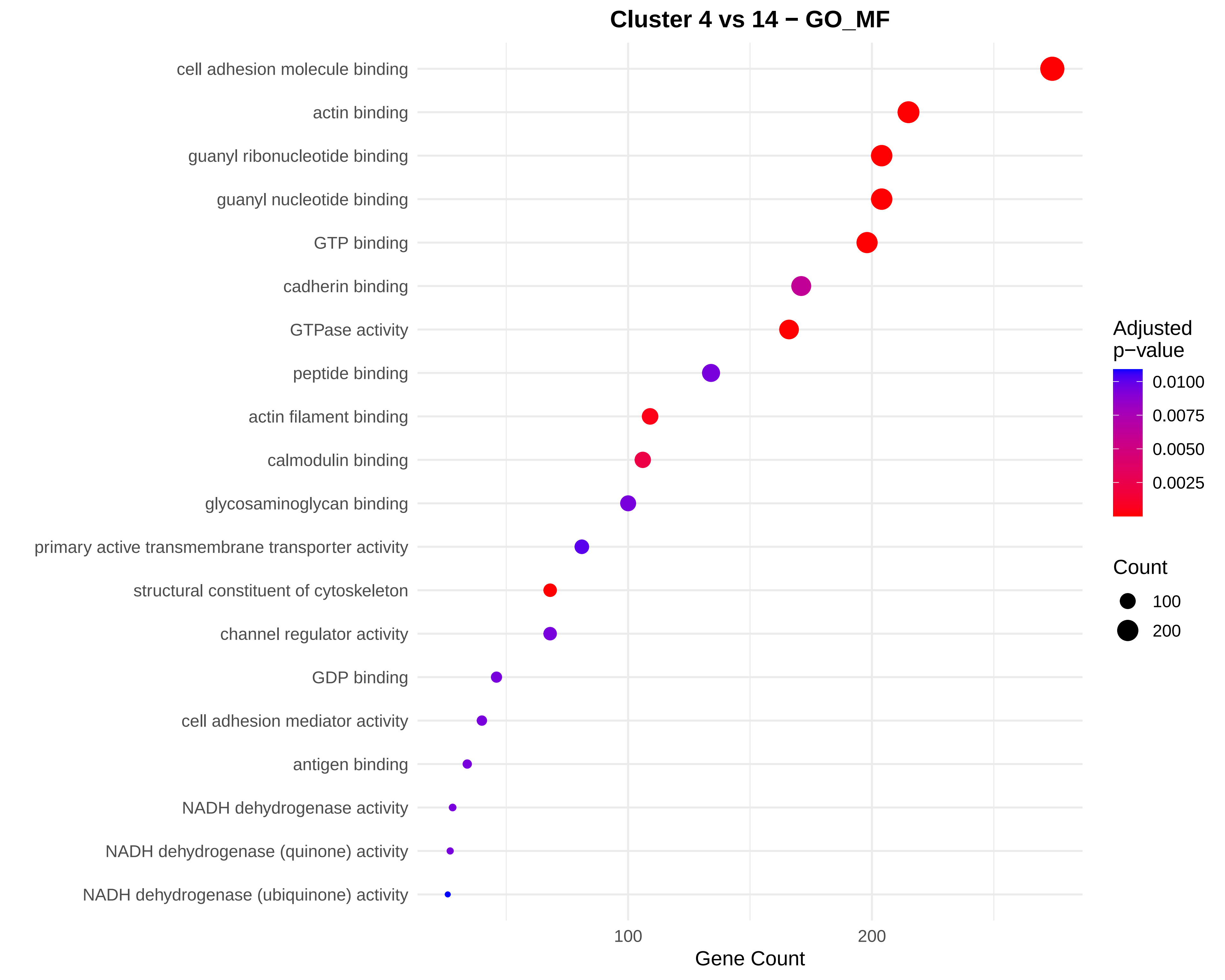

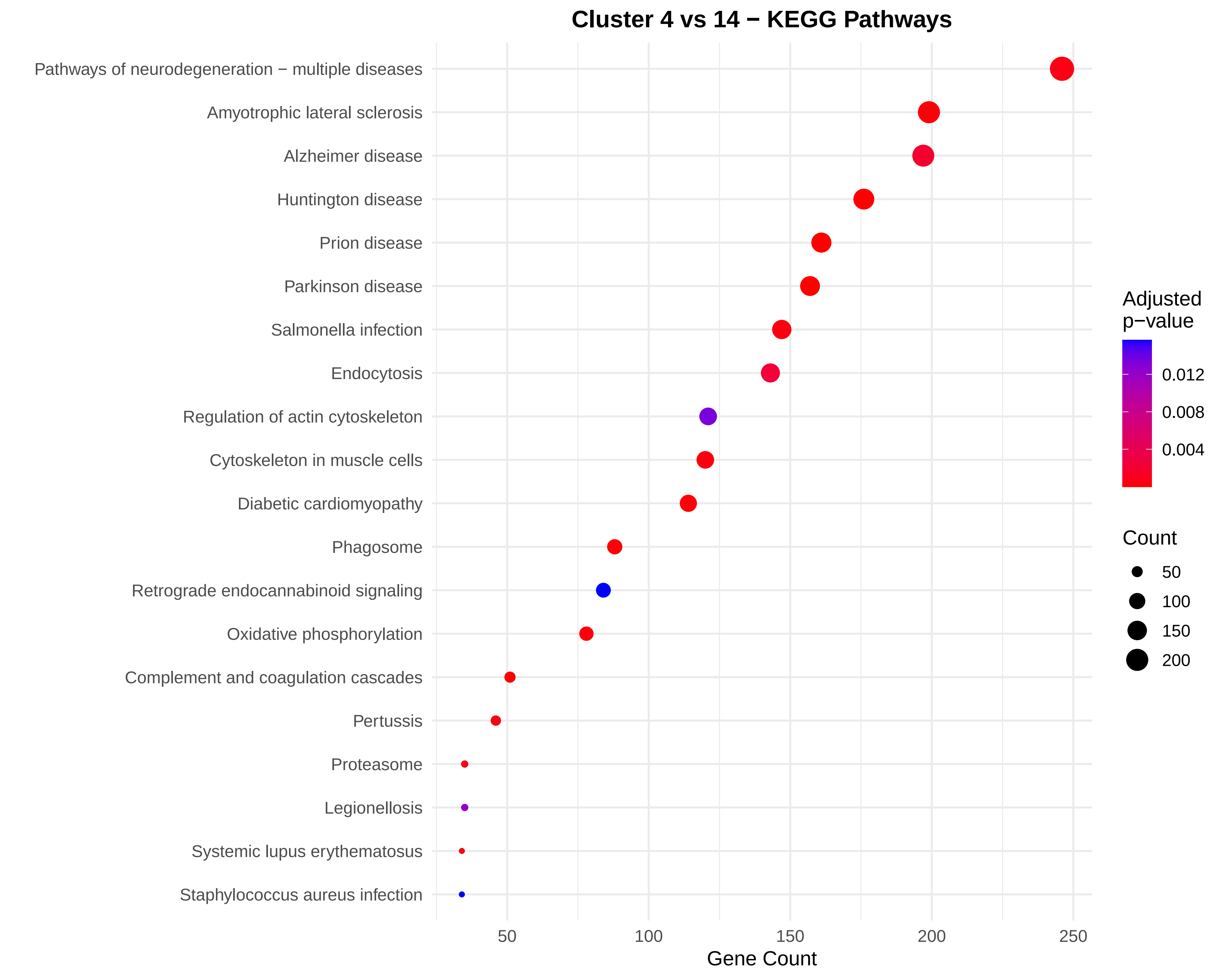

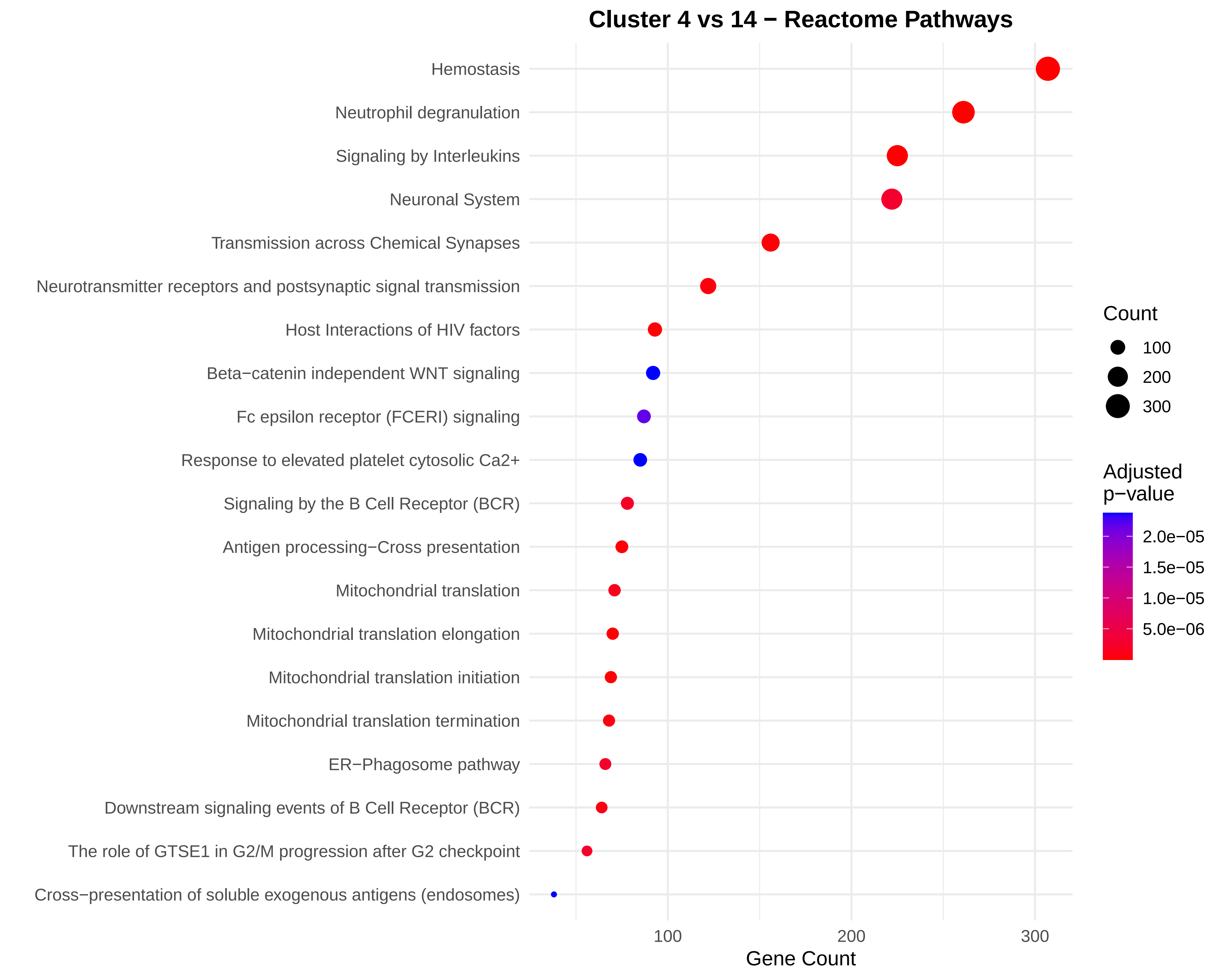

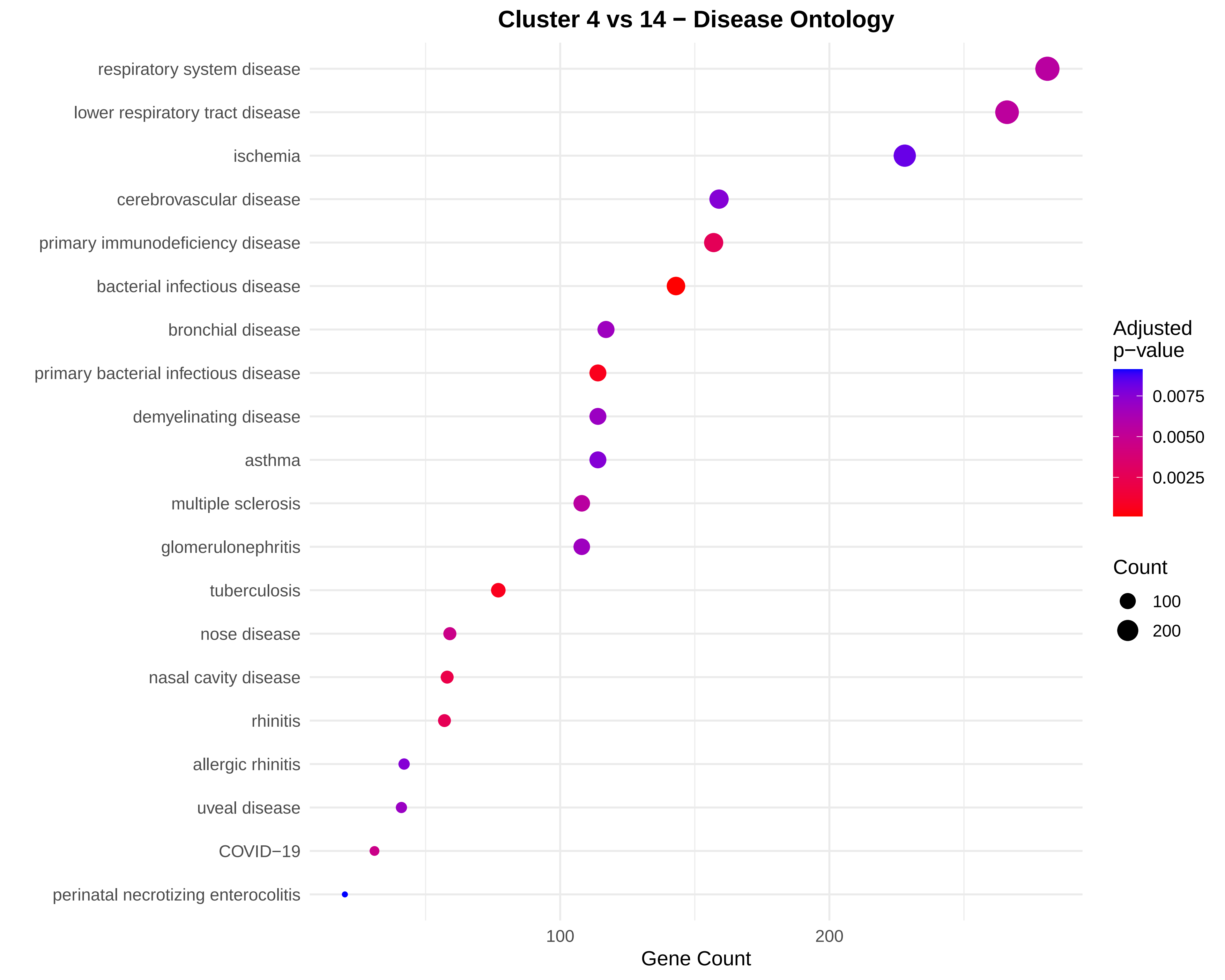

### Figure S17.pdf

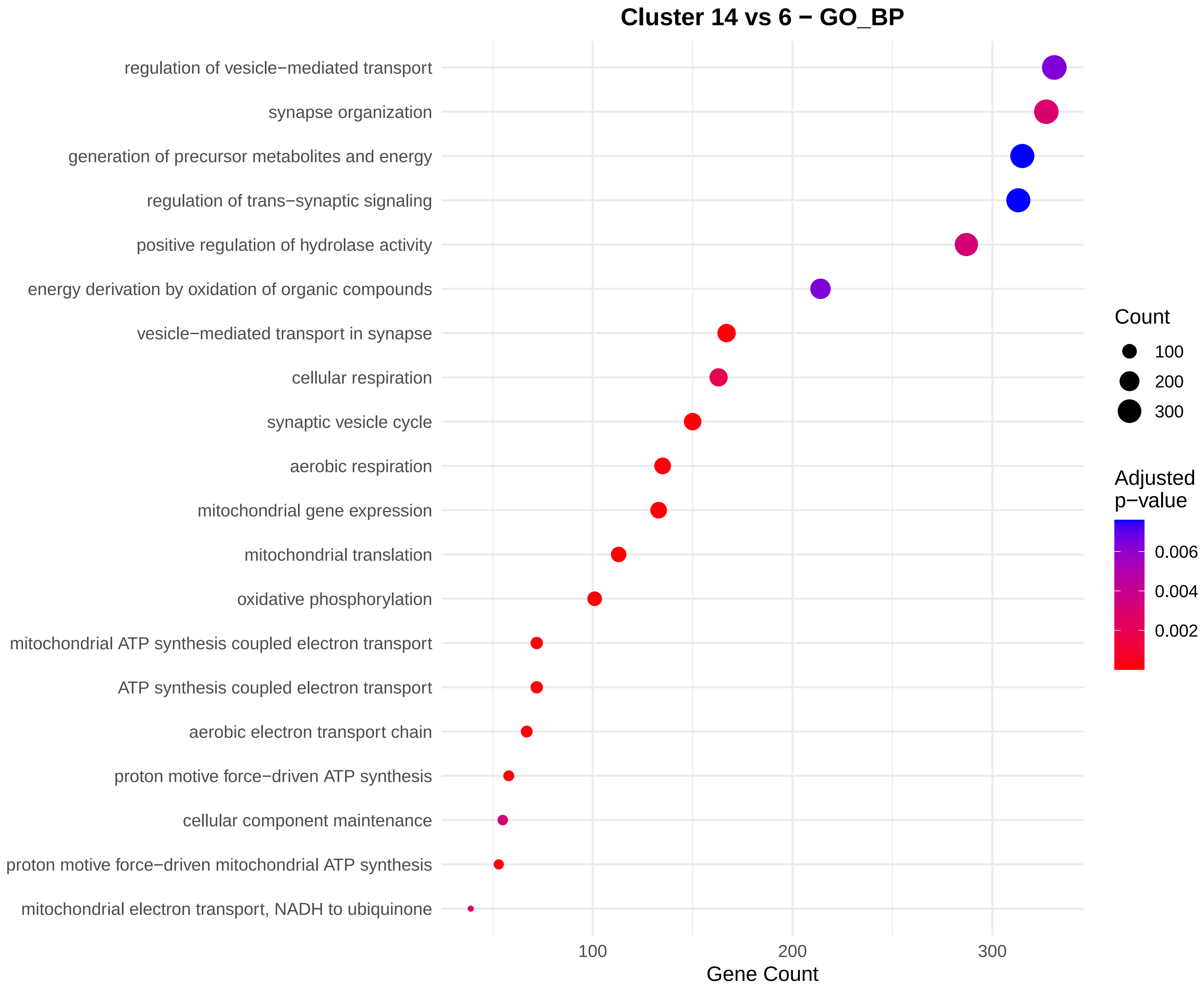

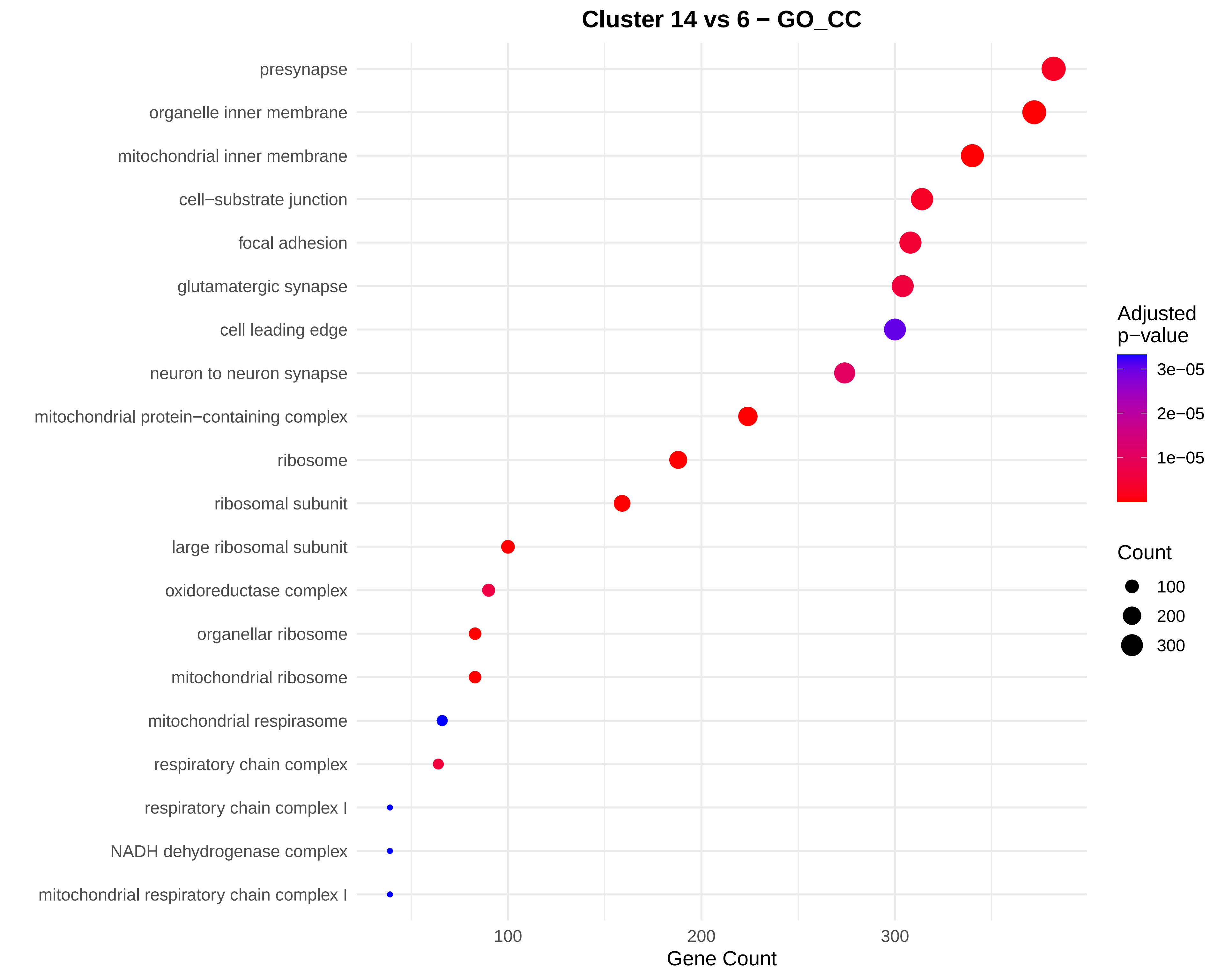

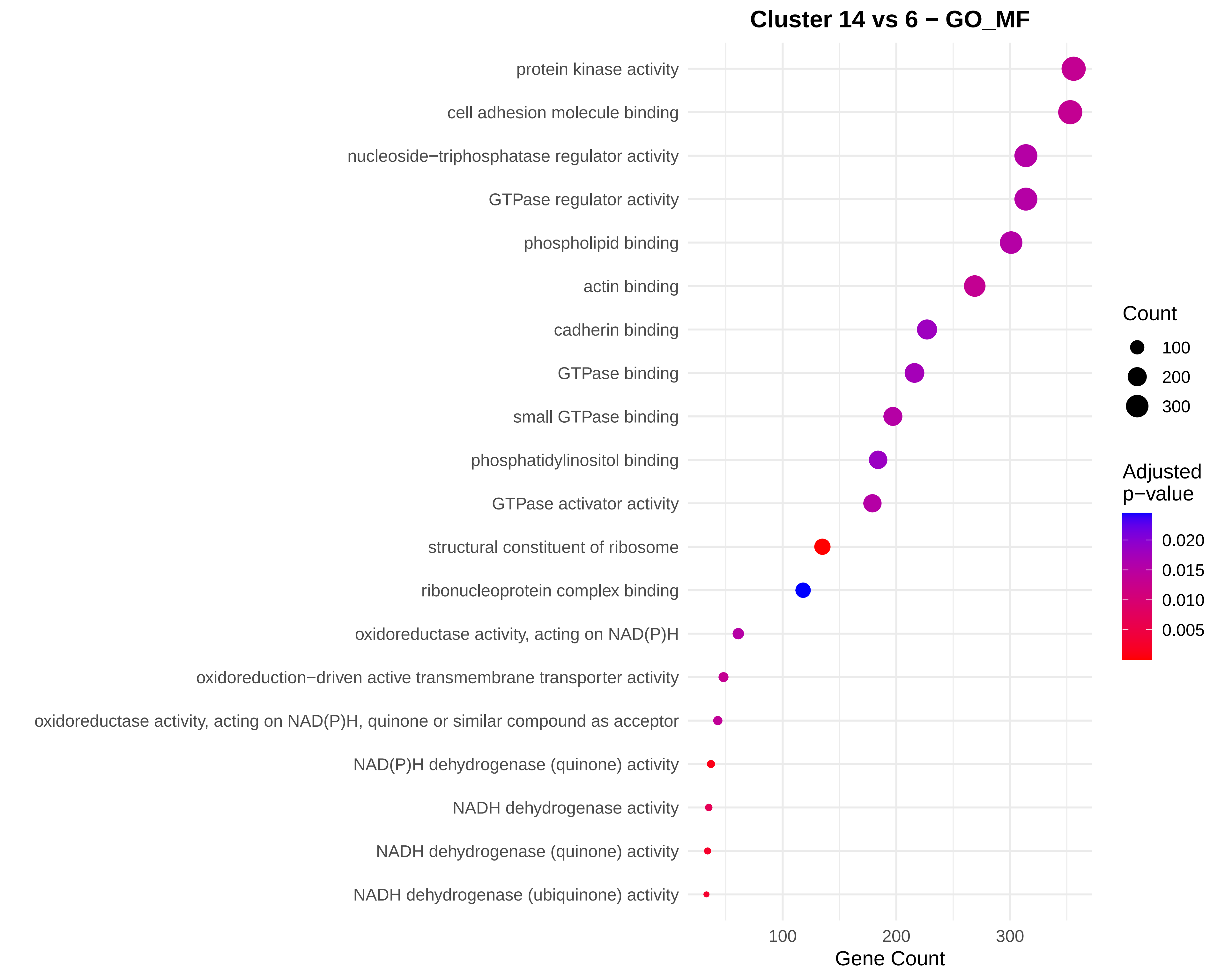

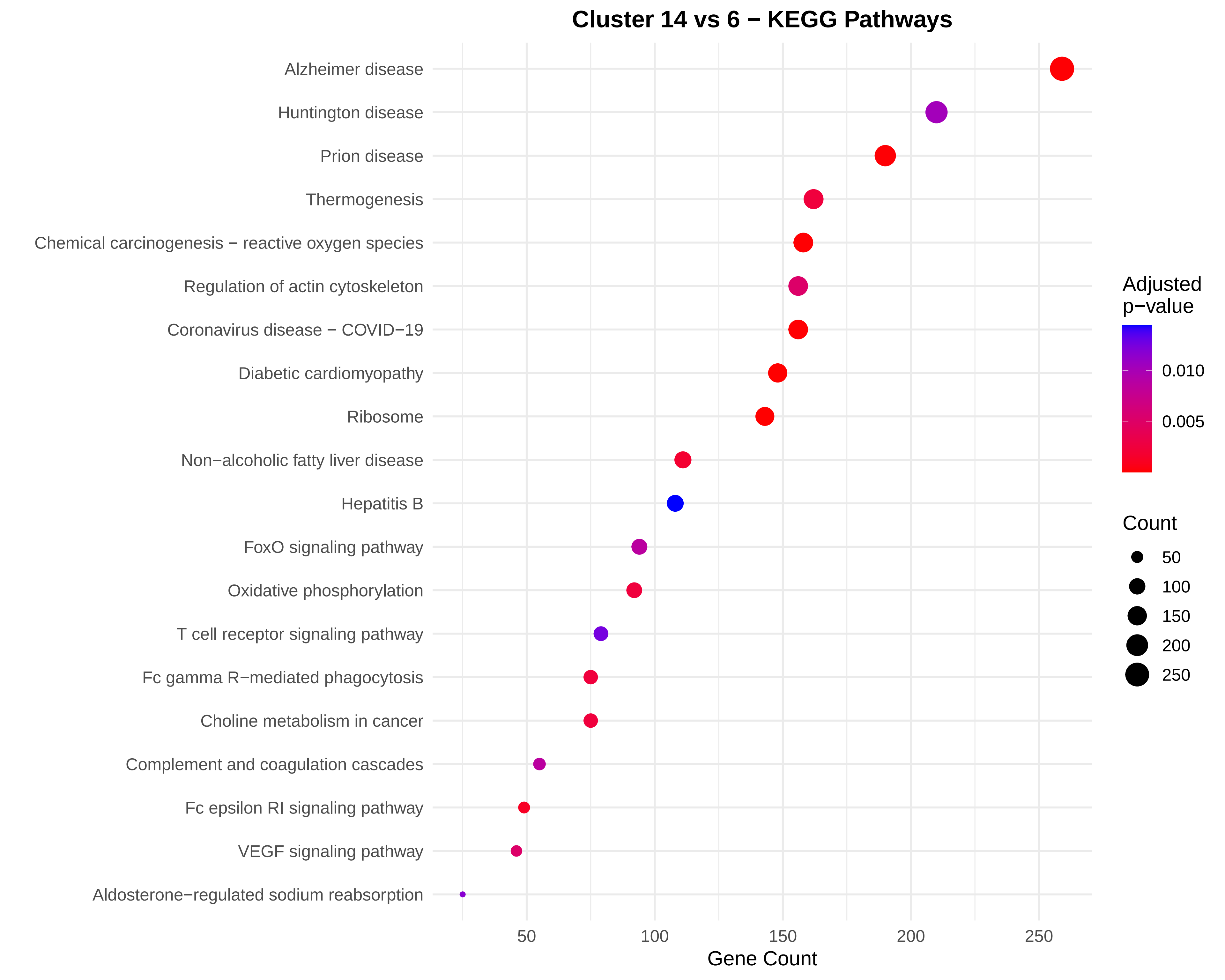

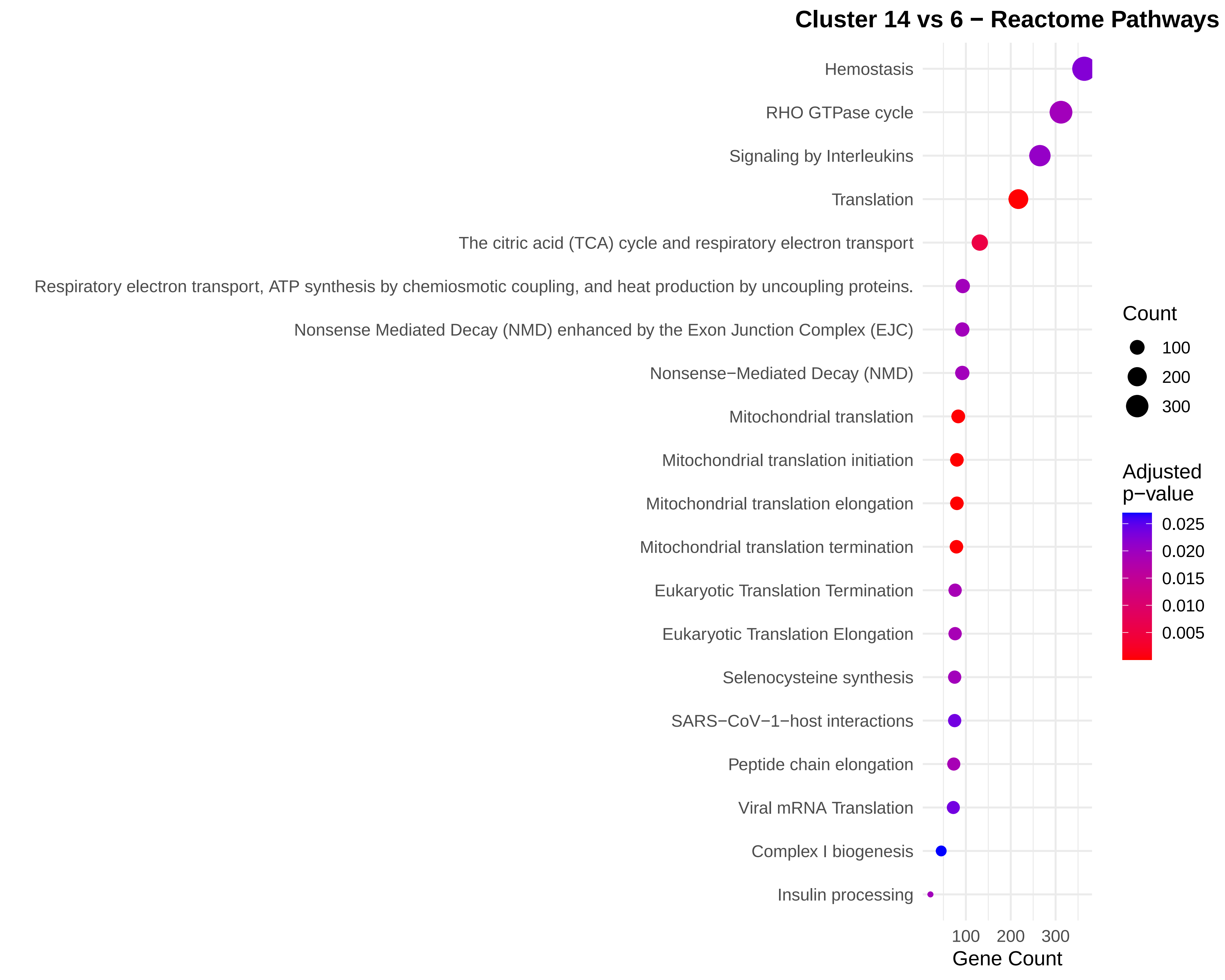

### Figure S18.pdf

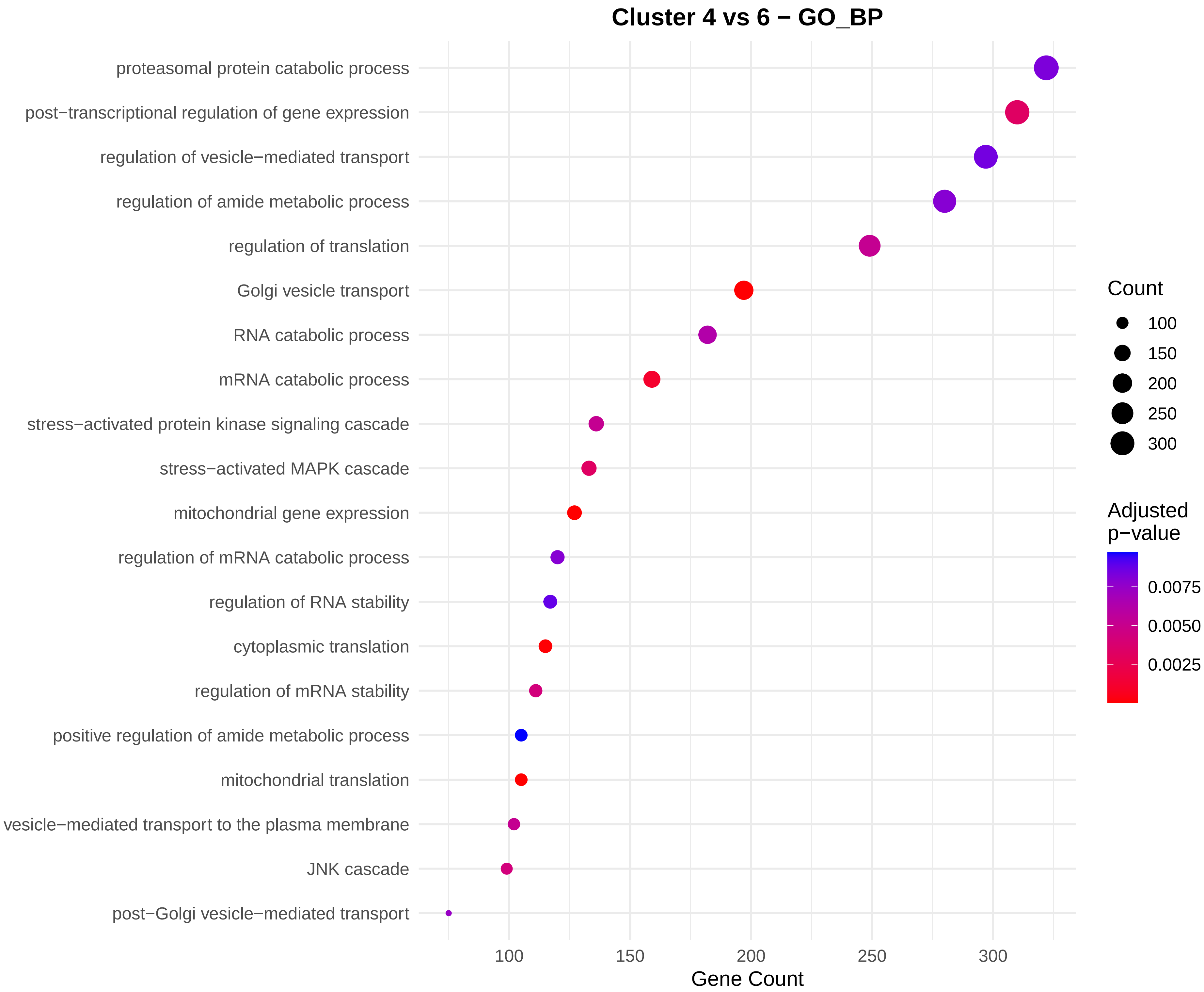

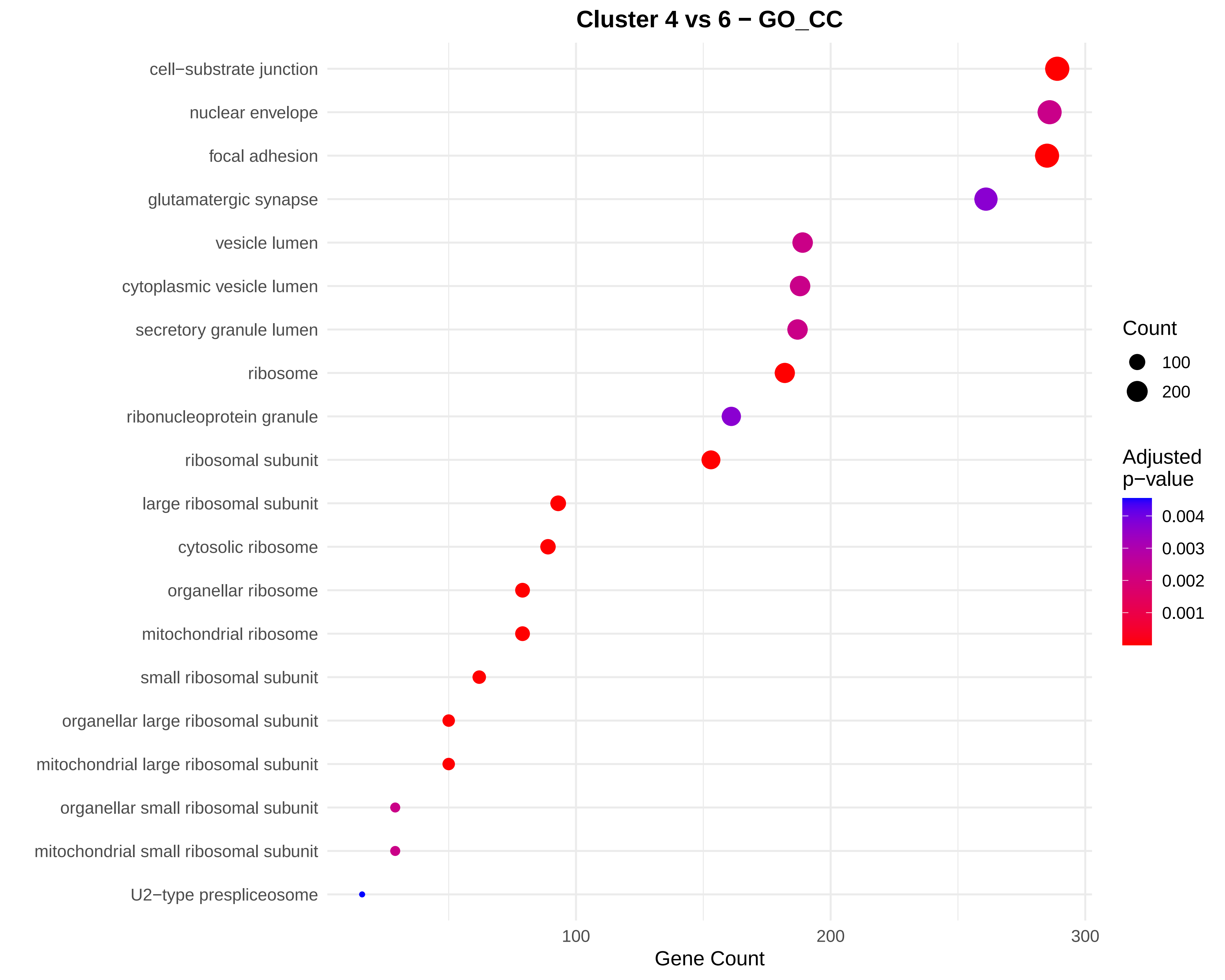

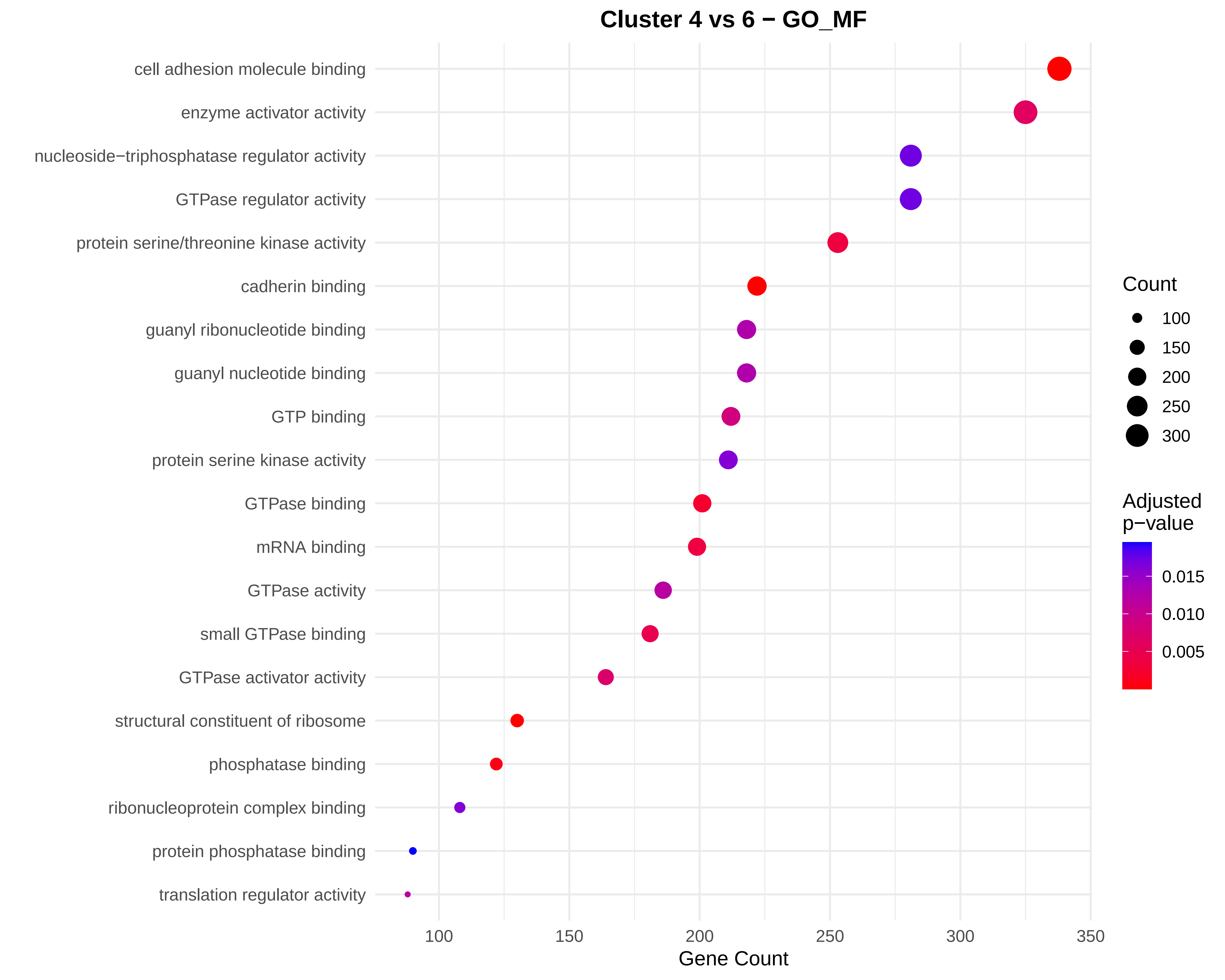

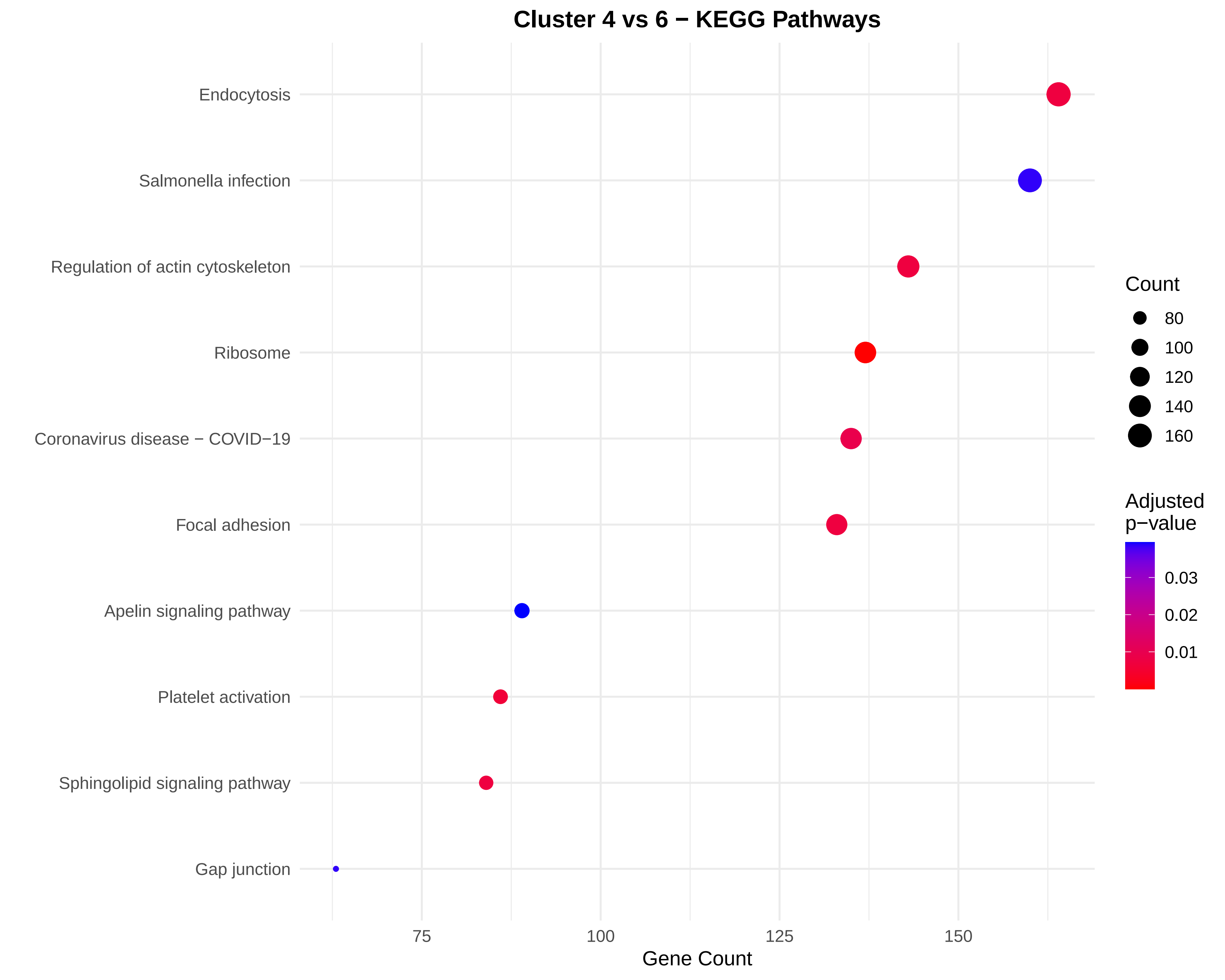

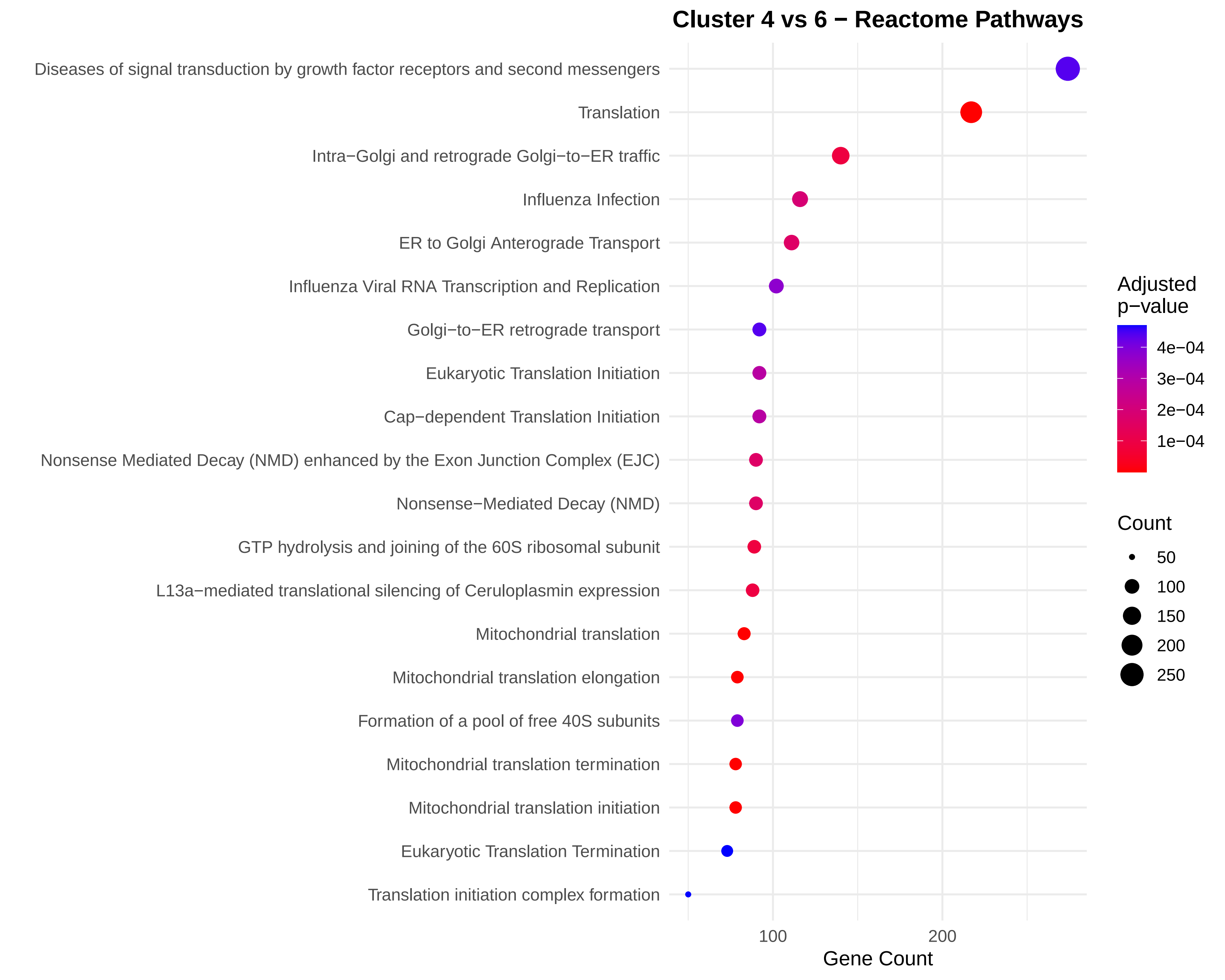
